# Functionally informed cis and trans proteome-wide association studies prioritize disease-critical genes

**DOI:** 10.64898/2026.04.24.26351667

**Authors:** Kangcheng Hou, Ali Pazokitoroudi, Benjamin Strober, Xilin Jiang, Alkes L. Price

## Abstract

Proteome-wide association studies (PWAS) typically link genetically predicted protein levels to disease using cis-pQTLs, which can be limited by low cis-heritability for disease-critical genes under negative selection and by tagging due to co-regulation among nearby genes. Trans-pQTLs provide complementary information when large sample sizes are available to detect weak polygenic effects, enabling associations between trans-predicted protein levels and disease. We developed PolyPWAS, a functionally informed, summary statistics-based framework for associating both cis- and trans-predicted protein levels to disease. PolyPWAS integrates 96 functional annotations with proteome-wide pleiotropy to improve protein prediction, while correcting for PCs of predicted protein levels to limit tagging effects. We applied PolyPWAS to 2.8K plasma proteins measured in 34K UKB-PPP participants, analyzing GWAS summary statistics for 88 diseases and complex traits (average *N*=336K). Trans-predicted protein levels explained 21% of disease heritability (vs. 9.6% for cis-predicted protein levels), leveraging a 24% relative improvement in trans-prediction accuracy from functional priors. Trans-PWAS identified more significant protein-disease associations (and more conditionally significant associations) than cis-PWAS. Cis and trans associations showed only modest excess overlap (1.18, 95% CI: 1.11-1.26). Accordingly, combining evidence from cis and trans associations improved disease gene prioritization evaluated using gene sets from rare variant association studies (+11% relative improvement) and PoPS (+7.0% relative improvement) relative to cis-only approaches. PWAS associations to disease replicated across protein level cohorts, with strong UKB-PPP/deCODE concordance after adjusting for cohort-specific prediction accuracy. We provide examples where trans-regulatory effects link multiple disease-critical genes, underscoring the importance of integrating cis- and trans-regulatory effects to map protein-mediated disease biology.

## Introduction

Integrative analyses combining molecular quantitative trait loci (QTLs) and genome-wide association studies (GWAS) have proven critical for understanding how genetic variants perturb molecular intermediates to influence disease risk^1–17^. Transcriptome- and proteome-wide association studies (TWAS/PWAS)—which construct genetic predictors of gene expression or protein abundance and test their association with disease—have shown particular promise in prioritizing disease-critical genes. The majority of these efforts have focused on cis-regulatory effects—which are more readily detectable with limited sample sizes, yet cis heritability tends to be low for evolutionarily constrained genes that are disproportionately important in disease ^12,13,18–20^. Trans-regulatory effects, which generally exceed cis effects in their contribution to molecular trait heritability^21–24^, can provide key insights about disease architectures^24–29^. Recent large-scale protein QTL (pQTL) studies have enabled systematic discovery of trans-pQTLs^23,30–33^, offering complementary information about distal regulatory effects. However, existing PWAS approaches that use cis and trans information typically restrict analyses to genome-wide significant trans-pQTLs and/or do not model functional enrichment of pQTLs^34–40^, thereby overlooking the polygenic functional architecture of protein regulation^21,23,25,27^. In addition, prior work has not systematically compared cis versus trans information or quantified their joint contribution to disease gene prioritization.

Here, we introduce PolyPWAS, a functionally informed summary statistics-based PWAS method that models genome-wide pQTL effects^41^ using functional enrichment from 96 baseline-LD model annotations^42^ and annotations representing cis and trans pleiotropy across the proteome, using only pQTL and GWAS summary statistics and reference LD^41–43^. PolyPWAS also adjusts for principal components of predicted protein levels to account for shared pQTL effects that influence many proteins. We show that trans-predicted protein levels explain substantial disease heritability beyond cis-predictions, leveraging improvements in trans-prediction accuracy from functional enrichment. Using independent validation data from rare variant association studies (RVAS)^44^ and polygenic priority score (PoPS)^45^, we show that disease genes supported by both cis and trans PWAS provide improved gene prioritization. We also highlight representative examples in which trans-regulatory effects connect multiple disease-critical genes.

## Results

### Methods overview

PolyPWAS improves genetic prediction of protein levels using either cis-pQTLs (±1Mb from transcription start site) or trans-pQTLs by integrating functional annotations and proteome-wide pleiotropy with summary statistics-based pQTL modeling and then associates cis- and trans-predicted protein levels with disease using GWAS summary statistics. To obtain protein prediction weights from pQTL summary statistics, we apply SBayesRC^41^, an empirical Bayes method that produces posterior mean estimates of genome-wide causal pQTL effect sizes, using two types of functional information: (1) 96 baseline-LD model annotations capturing coding, regulatory, conservation, and LD-related features enriched for disease and molecular trait heritability^24,25,46^, and (2) proteome-wide pleiotropy, defined by whether each SNP acts as a significant cis-pQTL, trans-pQTL, or both across all profiled proteins in a given pQTL study.

In detail, we obtain protein prediction weights 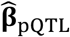 using SBayesRC (representing posterior mean causal effects, or best-fit effects in the case of missing SNPs) and partition them into cis 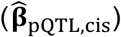 and trans 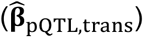 weights for each protein. We compute cis and trans PWAS association Z-scores by correlating genetically predicted protein levels with disease risk, accounting for LD^2^:

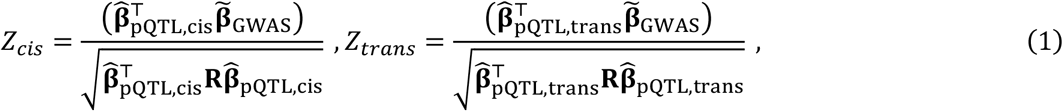

where 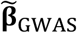 are marginal GWAS effects, and **R** is the reference LD matrix. Equation (1) separately considers cis and trans components, which may prioritize distinct gene-disease associations. To account for shared pQTL effects that influence many proteins, we adjust for 20 principal components of predicted protein levels (PPCs), separately for cis and trans (**Methods**). We determined that this adjustment reduces correlations between predicted protein levels due to shared pQTL effects and improves gene prioritization specificity (see below). We have publicly released PolyPWAS software (**Code Availability**).

We applied PolyPWAS to the UKB-PPP plasma pQTL study^23^ (34K individuals and 2.8K proteins), deCODE plasma pQTL study^31^ (35K individuals and 4.7K proteins), and a cerebrospinal fluid (CSF) pQTL study^32^ (3.5K individuals and 6.4K proteins) (**Supplementary Table 1**). We analyzed 88 heritable GWAS diseases/traits, including 32 approximately independent UK Biobank diseases/traits (with UKB-PPP samples excluded^47^) and 56 additional diseases/traits with publicly available summary statistics (average *N*=336K; **Supplementary Table 2**). We analyzed 7.7 million well-imputed variants with minor allele frequency (MAF)≥0.01 (or 1.2 million HapMap3 SNPs^48^), using LD matrices derived from UK Biobank genotypes^41^. In UKB-PPP data, the availability of individual-level genotypes and protein levels enabled direct evaluation of cis and trans prediction accuracy in a held-out sample: protein prediction models were trained on 33K unrelated white British individuals and evaluated in a held-out set of 10K white British individuals; downstream PWAS analyses for UKB-PPP used publicly available pQTL summary statistics constructed from a set of 34K individuals of European ancestry^23^. We validated the prioritized gene-trait pairs against genes implicated by RVAS^44^ providing specific causal evidence, and genes prioritized by PoPS^45^ capturing network-level convergence. We have publicly released all pQTL association statistics, protein prediction models, GWAS association statistics, PWAS association statistics, and validation metrics from this study (**Data Availability**).

### Simulations

We performed simulations to evaluate the calibration and power of PolyPWAS for detecting protein-disease associations; we focused our simulations on trans-PWAS, as cis approaches have been shown to be well-calibrated and well-powered under realistic architectures^1,2,8^. We simulated trans-pQTL (*N*=40K) and GWAS (*N*=300K) summary statistics using LD matrices from UK Biobank European-ancestry individuals restricted to HapMap3 SNPs^48^. We simulated trans-pQTL effects with 1% causal variants and SNP-heritability (*h*^*2*^) ranging from 5% to 20%, yielding prediction accuracy (mean *R*^2^=0.6-2.6%; **Supplementary Table 3**) consistent with empirical data (see below); other parameter settings were also explored. We simulated GWAS summary statistics with protein-mediated effects (mediated *h*^*2*^=5×10^-4^ per protein). We also simulated varying non-mediated genetic effects (non-mediated *h*^*2*^=0-20%) to evaluate tagging at non-causal proteins that are correlated with the causal proteins^9,10,15–17,49^. We applied SBayesRC^41^ (without functional or pleiotropy annotations) to simulated pQTL data to obtain trans-pQTL prediction weights and applied PolyPWAS for association testing (**Methods**).

We first evaluated calibration in null simulations without non-mediated GWAS effects (**Figure 1a** and **Supplementary Table 3**). At a significance threshold of p<10^-5^ (matching the threshold used in real data analyses), PolyPWAS maintained a well-calibrated false positive rate (FPR) across all values of trans-pQTL *h*^*2*^, confirming that the test statistic is properly calibrated.

**Figure 1.**
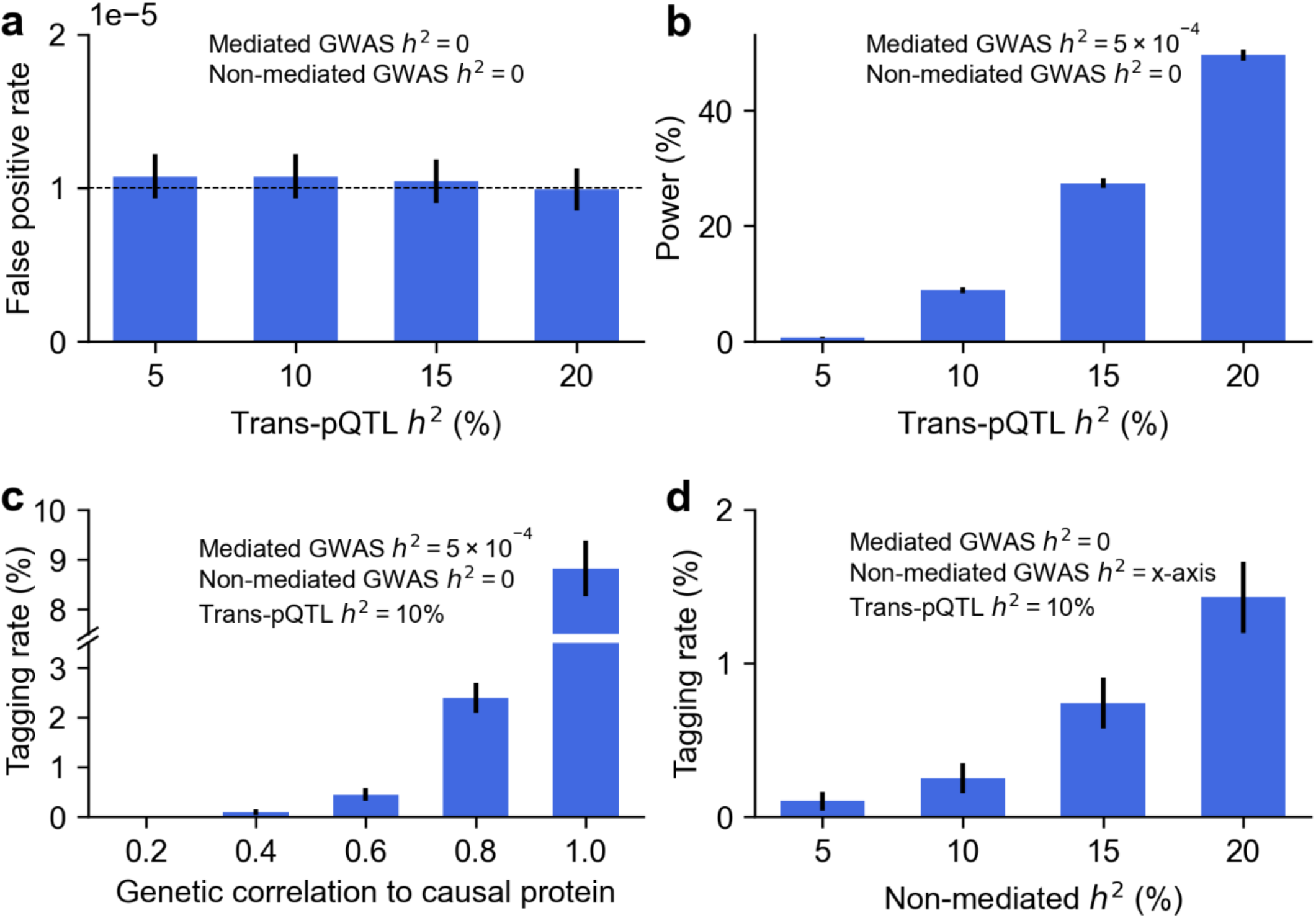
Simulations evaluating PolyPWAS calibration, power, and tagging associations. **(a)** False positive rate of protein-disease associations in null simulations without protein-mediated effects. Dashed line denotes expected FPR of 10^-5^. **(b)** Power to detect protein-disease associations in causal simulations. **(c)** Tagging association rate at non-causal proteins as a function of genetic correlation with the causal protein. **(d)** Tagging association rate as a function of non-mediated heritability. Numerical results are reported in **Supplementary Table 3**.

We next evaluated power to detect true protein-disease associations (**Figure 1b** and **Supplementary Table 3**). Under a realistic mediated *h*^*2*^ of 5×10^-4^ per protein, PolyPWAS power increased with trans-pQTL *h*^*2*^ (e.g. 50% at trans-pQTL *h*^*2*^*=*20%), reflecting improved accuracy of genetically predicted protein levels for more heritable proteins.

We also considered two scenarios (with trans-pQTL *h*^*2*^ fixed at 10%) in which non-mediated genetic effects produce tagging associations at non-causal proteins^9,10,15–17,49,50^. First, in simulations where non-causal proteins are associated because they tag a causal protein through genetic correlation (**Figure 1c** and **Supplementary Table 3**), the tagging association rate increased steeply with stronger correlation, mirroring the power-*h*^*2*^ pattern in **Figure 1b** and confirming that tagging associations arise when proteins share substantial trans-regulatory effects. Second, in simulations where non-causal proteins are associated because genetic variants influence disease risk through polygenic variant effects independent of the focal protein (**Figure 1d** and **Supplementary Table 3**), the tagging association rate increased with higher non-mediated heritability (e.g. 1.4% at non-mediated heritability of 20%), confirming that tagging associations arise from polygenic variant effects. Both findings underscore the need to account for tagging effects (**Discussion**).

We performed three secondary analyses to assess how pQTL and GWAS polygenicity affect power and tagging rates. First, we varied pQTL polygenicity in **Figure 1b** (0.1%, 1%, and 10% causal pQTL variants) while fixing trans-pQTL *h*^*2*^ at 10%. Power increased as trans-pQTL polygenicity decreased, consistent with higher protein level prediction accuracy under less polygenic architectures (**Supplementary Figure 1a**). Second, we varied pQTL polygenicity in **Figure 1d** (0.1%, 1%, and 10% causal pQTL variants) while fixing non-mediated *h*^*2*^ at 10%. Tagging rates increased slightly with higher pQTL polygenicity, implying modest sensitivity to pQTL polygenicity (**Supplementary Figure 1b**). Third, we varied GWAS polygenicity in **Figure 1d** (0.1%, 1%, and 10% causal non-mediated variants) while fixing non-mediated *h*^*2*^ at 10%. Tagging rates were similar across settings, implying low sensitivity to GWAS polygenicity (**Supplementary Figure 1c**).

### Functionally informed cis and trans genetic prediction of protein abundance

We trained cis and trans prediction models using pQTL summary statistics for 7.7 million well-imputed variants (or 1.2 million HapMap3 SNPs^48^, for comparison purposes) (adjusting for age, sex, and 10 genetic PCs) across 2.8K plasma proteins measured in 33K UKB-PPP white British participants with matched genotype and proteomic data^23^ (**Methods**). Predicted protein levels were adjusted for 20 PPCs, separately for cis and trans. Prediction *R*^2^ was evaluated in held-out individuals by correlating genetically predicted versus measured protein levels. To quantify shared genetic regulation across proteins, we computed *co-regulation scores*, defined as the sum of squared correlations between each focal cis-(resp. trans-) predicted protein and all cis-(resp. trans-) predicted proteins including the focal protein^51^.

Functional priors substantially increased trans prediction accuracy: using all 7.7 million well-imputed variants: median *R*^*2*^ for trans predictors achieved a 24% relative improvement (from 0.25% to 0.31%; these results were adjusted for 20 PPCs; **Figure 2a** and **Supplementary Table 4**), although it remained far below the estimated trans SNP-heritability (median 12%; **Supplementary Table 4**), consistent with highly polygenic distal regulation^12,23,27,40^. Cis prediction improved more modestly (+5.7%, median from 0.70% to 0.74%), approaching the estimated cis-SNP heritability (median 0.79%). The mean *R*^*2*^ was substantially higher (trans 1.7%; cis 5.6%), driven by a small number of proteins with large values (**Supplementary Figure 2**); we therefore focus on medians in subsequent analyses to avoid results driven by outliers.

**Figure 2.**
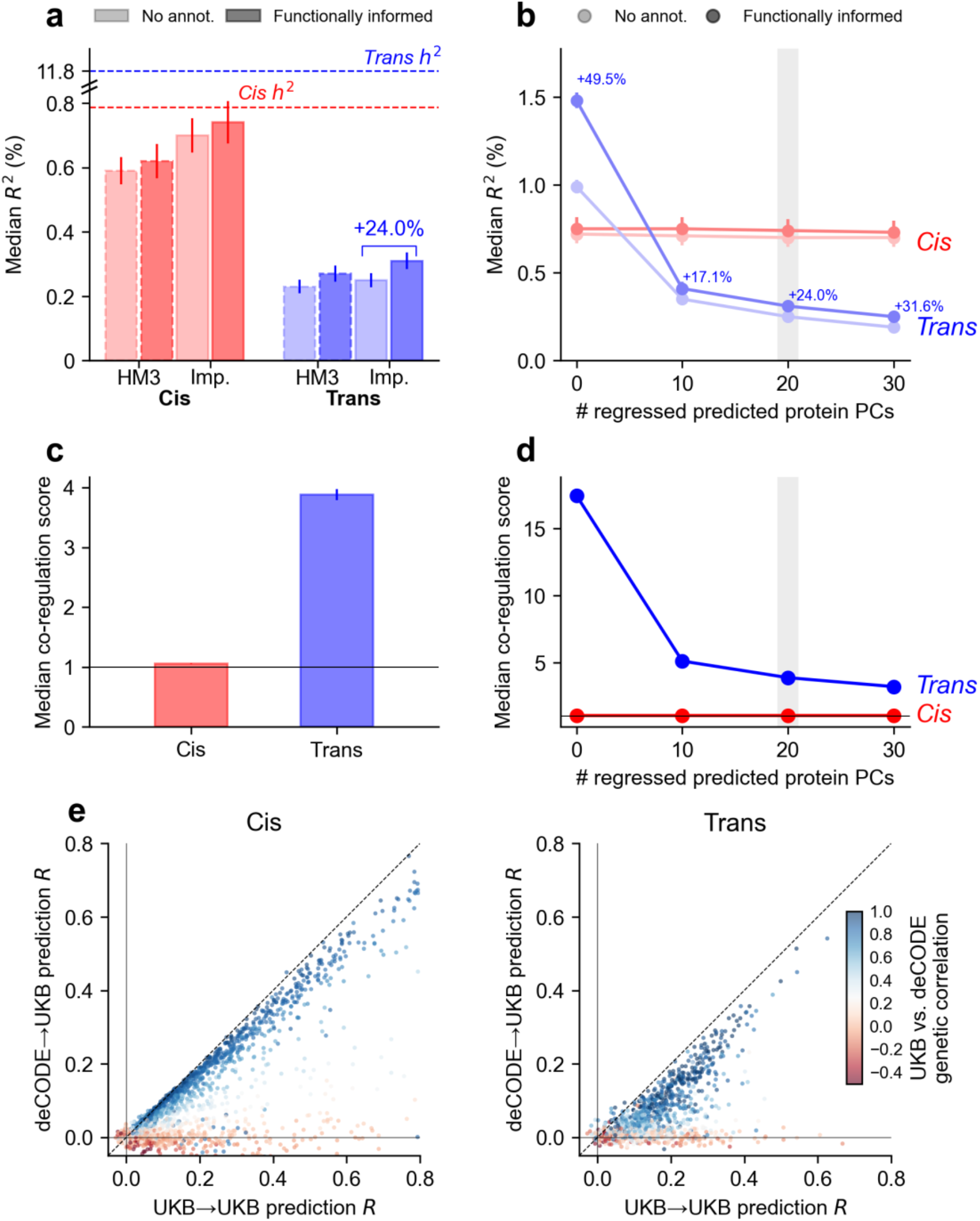
Functionally informed genetic prediction of protein levels. **(a)** Median prediction *R*^2^ across proteins for cis and trans prediction models using HapMap3 (HM3) or well-imputed (Imp.) variants, with and without functional priors (baseline-LD model and proteome-wide pQTL annotations), adjusting for 20 PPCs. Dashed lines indicate median cis and trans heritability estimates. **(b)** Median prediction *R*^2^ for cis and trans prediction models when adjusting for different numbers of PPCs. **(c)** Median cis and trans co-regulation score when adjusting for 20 PPCs. **(d)** Median cis and trans co-regulation scores when adjusting for different numbers of PPCs. **(e)** Comparison of prediction *R* in UKB-PPP vs. deCODE across matched proteins for cis (left panel) and trans (right panel); heatmaps denote corresponding UKB-PPP vs. deCODE genetic correlations. Numerical results are reported in **Supplementary Table 4-6**.

Using 7.7 million well-imputed variants instead of 1.2 million HapMap3 SNPs^48^ increased prediction accuracy by 14.8% for trans and 19.4% for cis, consistent with previous results on complex trait polygenic prediction^41^. The accuracy improvement from functional priors was larger with the expanded well-imputed variant set, likely because functional annotations are less informative when causal variants are missing. PPC adjustment reduced trans prediction accuracy (**Figure 2b, Supplementary Table 4**, and **Supplementary Figure 3**) but is preferred because it reduces trans co-regulation (see below). Across a range of PPC adjustments, functional priors consistently improved trans prediction relative to models without functional annotations. The largest improvement was observed without PPC adjustment (+49.5%), likely because trans-PPCs captured shared regulatory modules (the top 20 trans-PPCs explained 34.0% of variance in trans-predicted protein levels; **Supplementary Figure 4**) enriched in pleiotropic trans effects^30,31,52–54^ that are partially recapitulated by our functional prior.

Consistent with extensive distal regulation, trans-predicted proteins had much higher co-regulation scores (median 3.9) than cis-predicted proteins (median 1.1), even after adjusting for 20 PPCs (**Figure 2c** and **Supplementary Table 5**); we note that co-regulation scores depend on the limited number of proteins analyzed (2.8K in UKB-PPP).

Trans co-regulation decreased sharply with PPC adjustment (from 17.5 to 5.1 for 0 vs. 10 PPCs) and then plateaued (from 5.1 to 3.9 for 10 vs. 20 PPCs), whereas cis co-regulation was largely unchanged (**Figure 2d** and **Supplementary Table 5**). Although trans-eQTL studies are pervasively impacted by cell-type composition effects^27,29^, the impact of cell-type composition on trans-pQTL is subject to debate: the original UKB-PPP study reported that adjusting for cell-type proportions produced limited changes in trans-pQTL associations^23^, whereas a later study analyzing the same data reported that many trans-pQTL are likely impacted by cell-type composition effects^24^. We assessed the biological role of PPCs by correlating them to UK Biobank traits; the top trans-PPCs showed modest correlations with broad physiological measures (e.g., explaining 6.7% and 2.4% of variance in platelet and white blood cell counts respectively; **Supplementary Figure 5**), suggesting that cell-type composition has only a mild influence on trans-PPCs. These shared modules therefore likely reflect genuine biological coordination rather than cell-type composition effects—including known trans-pQTL hotspots near pleiotropic regulatory genes (e.g. *SH2B3, SERPINA1, GCKR*)^30,31,52,53^.

We evaluated cross-cohort prediction by applying deCODE-derived pQTL weights^31^ to UKB-PPP validation samples (**Figure 2e** and **Supplementary Table 6**). Despite similar sample size and tissue context, prediction accuracy decreased substantially across cohorts for both cis and trans models. Median cis *R*^2^ dropped from 0.74% to 0.21% (mean 5.6% to 3.4%) and median trans *R*^*2*^ dropped from 0.31% to 0.03% (mean 1.7% to 0.85%). Although some proteins had similar accuracy in deCODE vs. UKB-PPP, many showed markedly attenuated prediction accuracy in deCODE, consistent with platform-specific (SomaScan vs. Olink) assay designs and/or isoform-related measurement effects^55,56^. Indeed, cross-cohort portability (i.e. relative differences in prediction accuracy across cohorts) correlated strongly with cross-cohort genetic correlation estimates, and cis vs. trans cross-cohort genetic correlations were themselves concordant (*R*^2^ = 0.37; **Supplementary Figure 6**), suggesting that proteins with low cross-cohort portability are influenced by additional sources of assay- or platform-specific biological noise; accordingly, cross-cohort portability also correlated with individual-level protein measurement concordance across platforms (**Supplementary Figure 7**). In instances where two deCODE aptamers target the same protein, we detected differences between those aptamers in cross-cohort genetic correlations and cross-cohort prediction accuracies for both cis and trans models (**Supplementary Figure 8**), further confirming that these aptamers capture distinct biological variables.

We performed 7 secondary analyses. First, we extended our cross-cohort analyses by applying CSF-derived pQTL weights (instead of deCODE-derived pQTL weights) to UKB-PPP validation samples. Prediction accuracy decreased more substantially for CSF than for deCODE, consistent with the smaller CSF sample size (3.5K vs. 35K) and tissue differences (**Supplementary Figure 9**). Second, we evaluated protein prediction accuracy and co-regulation scores in deCODE and CSF datasets (instead of UKB-PPP), which showed patterns broadly similar to UKB-PPP (**Figure 2a-d**) despite reduced power in CSF (**Supplementary Figure 10** and **Supplementary Figure 11**). Third, a prediction model using only 96 baseline-LD model annotations (without genome-wide pleiotropy annotations) outperformed a prediction model using no annotations but underperformed our full model (**Supplementary Figure 12**). Fourth, we assessed prediction accuracies in non-British European individuals vs. white British individuals. Prediction accuracies were highly concordant for both cis and trans predictions (**Supplementary Figure 13**). Fifth, we compared deCODE/UKB-PPP cross-cohort prediction accuracy restricted to cis proximal vs. non-proximal variants, the former being susceptible to platform-specific coding-variant epitope effects that alter assay affinity rather than true abundance^23,55–57^. We observed comparable cross-cohort portability for both classes, suggesting that epitope effects alone do not explain the observed differences in cis prediction across platforms (**Supplementary Figure 14**). Sixth, we compared UKB-PPP cis-predicted plasma protein levels with cis-predicted expression levels across 49 GTEx tissues^12^. Concordance was highest for liver, consistent with its central role in plasma protein production^58^, and lowest for brain and testis, likely due to restricted protein exchange across biological barriers (**Supplementary Figure 15**); trans-eQTL prediction models were not available with sufficient power to be included in this analysis. Seventh, we applied stratified LD score regression^46^ to partition trans-pQTL heritability across baseline-LD functional annotations, and observed significant enrichment for most functional annotations (**Supplementary Figure 16**), broadly consistent with previously reported enrichment results for trans-eQTLs mimicking complex traits^25^.

### Cis and trans predicted protein-disease associations across 88 diseases/traits

We analyzed GWAS summary statistics for 32 UK Biobank and 56 additional diseases/traits (average *N*=336K) (**Supplementary Table 2**), performing two complementary analyses (**Methods**). First, we estimated the proportion of disease SNP-heritability explained by cis- and trans-predicted protein levels (adjusted for 20 PPCs), using an approach applied to individual-level UK Biobank data that is robust to modeling assumptions^59^ and captures both mediated^13^ and non-mediated^2,51^ effects. Second, we applied cis- and trans-PWAS to GWAS summary statistics to identify protein-disease associations. Because summary statistics for most of the 56 additional diseases/traits were available only for HapMap3 SNPs, we restricted analyses of these 56 diseases/traits to HapMap3 SNPs.

Across 32 independent UK Biobank diseases/traits, cis-predicted protein levels explained a median of 9.6% of disease SNP-heritability, whereas trans-predicted levels explained a median of 21% (**Figure 3a** and **Supplementary Table 7**). Cis and trans contributions were largely additive, explaining a median of 26% jointly. However, several traits—including lipid levels, platelet count, red blood distribution width—exhibited notable cistrans redundancy, with a combined model explaining far less variance than the sum of the two components. For LDL cholesterol, cis-predicted levels explained 26±0.3% and trans-predicted levels explained 45±0.3% of SNP-heritability, but the combined model explained only 50±0.4%. The top cis-PWAS protein, *APOE*, accounted for 38±2.5% of cis-explained heritability in LDL, yet 93% of its cis-predicted level could be explained by trans-predicted levels of other proteins, including 63% explained by *SNAP25* alone, consistent with *APOE* regulating *SNAP25* levels^60^. These results indicate that disease-associated trans-predicted protein levels can reflect downstream effects of cis regulation, consistent with previous gene expression studies^61^.

**Figure 3.**
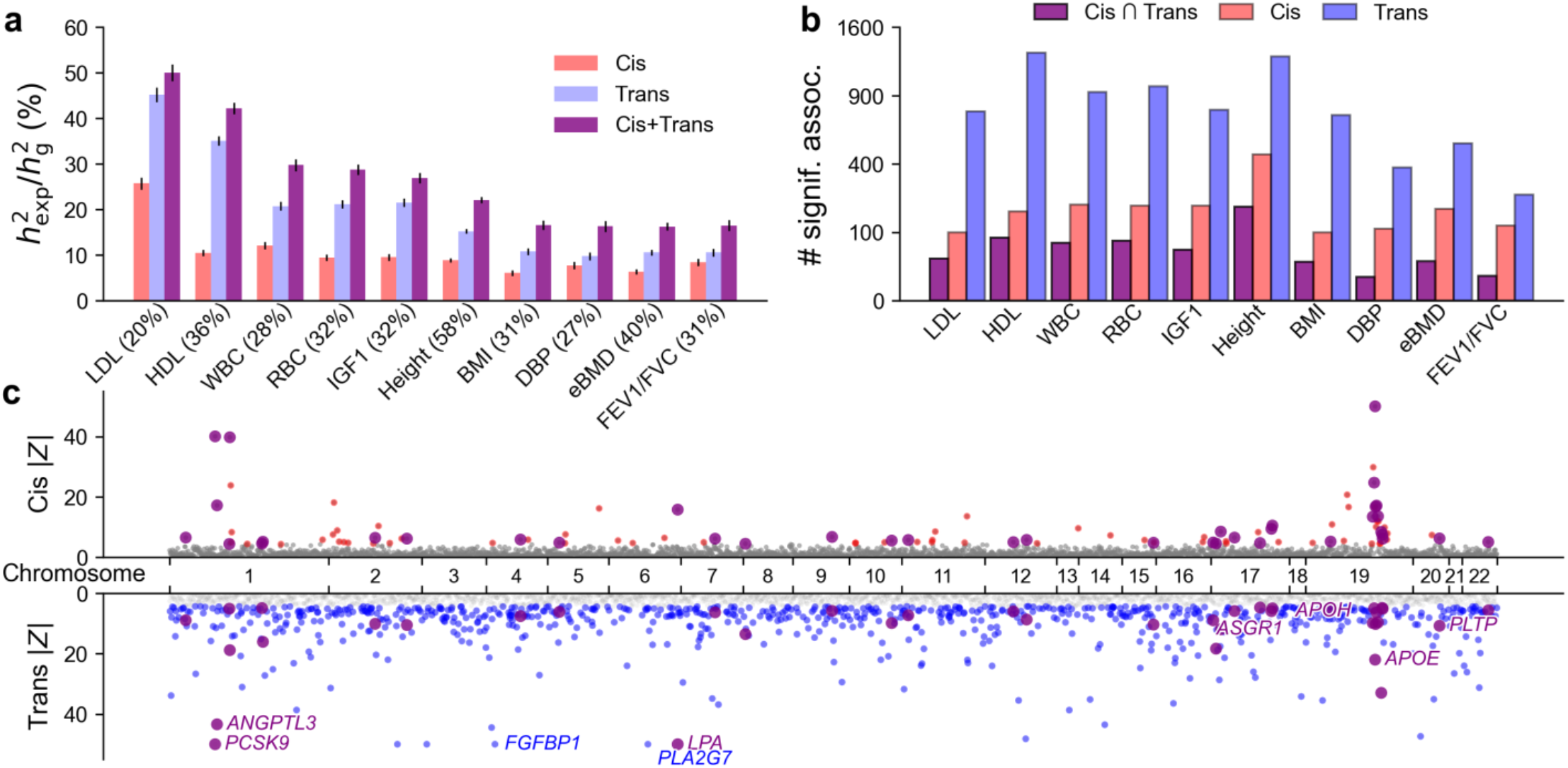
Protein-disease associations across UK Biobank diseases/traits. **(a)** Proportion of SNP-heritability explained by cis, trans, and combined cis and trans predicted protein levels (ratio between explained heritability 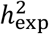 and total heritability 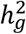) across 10 (of 32) selected UK Biobank traits. Trait labels include SNP-heritability estimates in parentheses. Error bars denote 95% confidence intervals. Traits are ordered by combined cis + trans proportion of explained heritability. **(b)** Number of significant protein-trait associations identified by cis-PWAS, trans-PWAS, and their intersection across 10 (of 32) selected UK Biobank traits. **(c)** Cis- and trans-PWAS associations for LDL cholesterol, with |Z| capped at 50 for visualization purposes. For **(a)** and **(b)**, numerical results for all 32 UK Biobank traits are reported in **Supplementary Table 8-9**. For **(c)**, full cis- and trans-PWAS association results for all traits analyzed have been publicly released (**Data Availability**).

We applied cis- and trans-PWAS to the 32 UK Biobank diseases/traits. Results are reported in **Figure 3b** and **Supplementary Table 8**. Trans-PWAS identified far more significant associations (P<10^-5^), implicating on average 717 proteins per trait (26% of all 2.8K proteins) versus 148 (5.3%) for cis-PWAS. Although cis and trans associations overlapped significantly (excess overlap = 1.18, 95% CI = [1.11, 1.26]; meta-analyzed across traits), most associations were distinct; the intersection of cis-PWAS and trans-PWAS associations implicated 51 proteins per trait (1.8%). The higher trans-PWAS discovery rate likely reflects both the greater variance explained by trans predictors (**Figure 3a**) and stronger co-regulation (**Figure 2c**), which increases tagging^9,10,15–17^. We estimated the number of conditionally independent protein-disease associations by applying stepwise regression and identified more conditionally independent trans-PWAS associations than cis-PWAS associations (average of 199 vs. 126 across traits; **Supplementary Figure 17**).

Prioritizing proteins with both cis and trans associations to LDL cholesterol identified key regulators (**Figure 3c**). Among 50 proteins prioritized in this way, the strongest trans associations included canonical lipid regulators (*PCSK9, ANGPTL3, PLTP*)^62–64^, major apolipoproteins (*APOE, APOH, LPA*)^65^, and genes involved in hepatic metabolism (*ASGR1*^66^, *FGF21*^67^). On the other hand, two proteins, *PLA2G7* and *FGFBP1*, showed trans but no cis association, despite being highly heritable in cis. Notably, *PLA2G7* is a known prognostic biomarker for CAD^68^, yet its loss-of-function variants show no protective association with CAD^69^, suggesting that the observed trans association reflects a downstream response to disease.

We next applied cis- and trans-PWAS to the 56 additional diseases/traits. Results are reported in **Figure 4a** and **Supplementary Table 8**. As in the analyses of UK Biobank diseases/traits, trans-PWAS identified far more significant associations, prioritizing 115 proteins per trait on average (4.1% of all 2.8K proteins) versus 27 (1.0%) for cis-PWAS (36 conditionally independent for trans, vs. 22 for cis); 5.3 proteins per trait (0.2%) were significant in both (excess overlap = 1.60, 95% CI = [1.39,1.84]). The reduced discovery rate vs. UK Biobank diseases/traits reflects lower statistical power due to lower GWAS SNP-heritability Z-scores (**Supplementary Table 2**).

**Figure 4.**
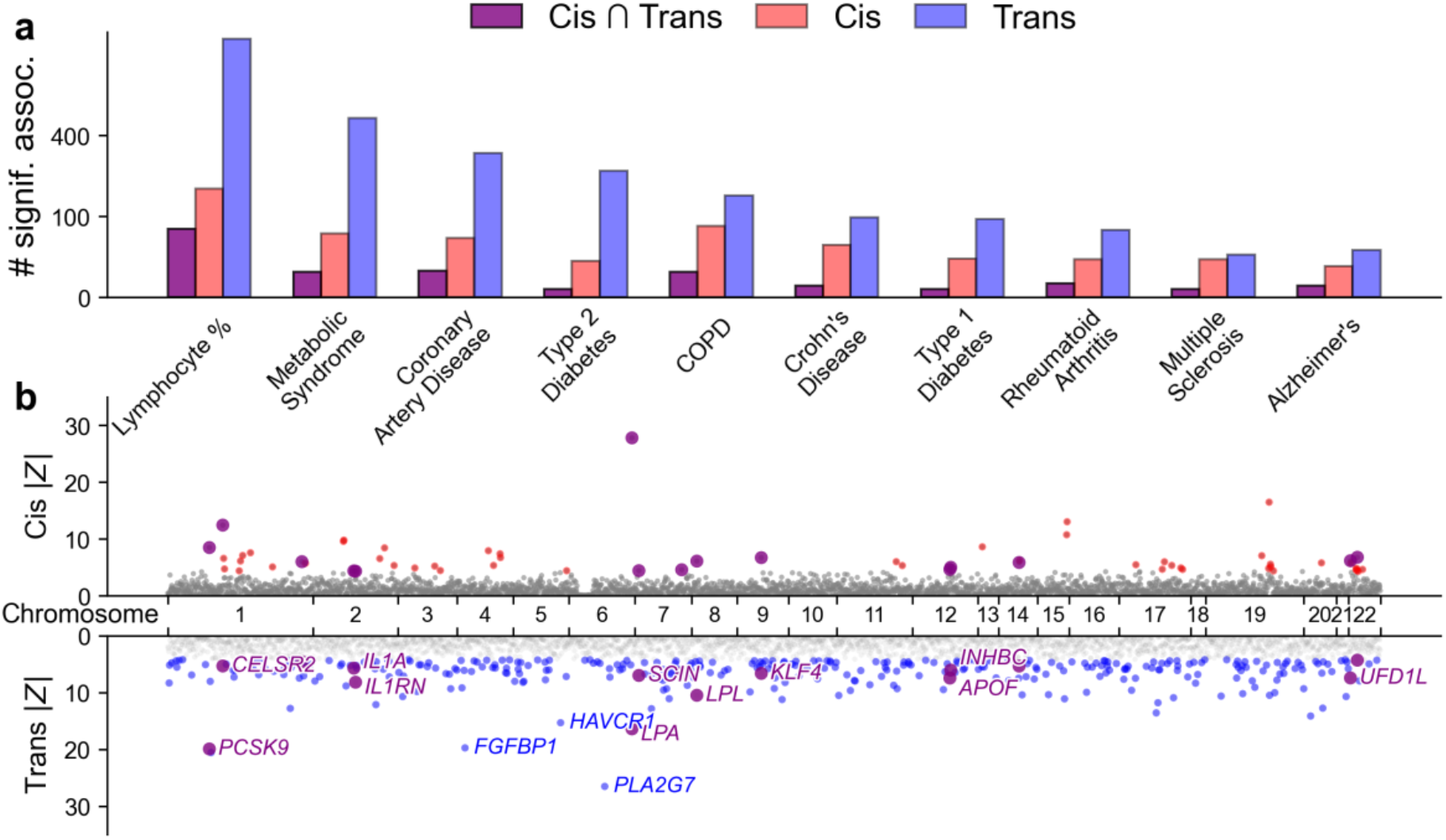
Protein-disease associations across additional diseases/traits. **(a)** Number of significant protein-trait associations identified by cis-PWAS, trans-PWAS, and their intersection across 10 (of 56) selected additional diseases/traits. Traits are ordered by number of intersections for both cis and trans associations. Numerical results for all 56 additional diseases/traits are reported in **Supplementary Table 8. (b)** Cis- and trans-PWAS associations for coronary artery disease (CAD), with |Z| capped at 50 for visualization purposes. Full cis- and trans-PWAS association results for all traits analyzed have been publicly released (**Data Availability**).

Prioritizing proteins with both cis and trans associations for coronary artery disease (CAD)^70^ identified key regulators (**Figure 4b**). The 15 proteins prioritized in this way included lipid metabolism genes (*PCSK9, LPL, LPA, APOF*)^71–73^, and a distinct inflammatory component driven by the interleukin-1 axis (*IL1A, IL1RN*), consistent with the role of innate immunity in atherogenesis^74^. Evidence for vascular remodeling and endothelial stress emerged through regulators such as *KLF4, PGF*^75,76^, while additional prioritized genes—including *AIDA*^77^, *SUSD2*^78^, and *UFD1L*^79^—implicate less well-characterized mechanisms linking metabolic dysregulation to downstream vascular responses. Together, these results show that integrating cis and trans evidence helps decompose CAD biology into metabolic, inflammatory, and vascular axes.

We assessed the cross-cohort consistency of PWAS associations across 88 diseases/traits by comparing UKB-PPP and deCODE results for 1,677 proteins shared between cohorts. After adjusting PWAS effect sizes to account for cohort-specific partial protein prediction accuracy (which differed between the two cohorts; **Figure 2e, Methods**), we observed strong concordance (**Figure 5** and **Supplementary Table 9**). Using Deming regression to account for effect size uncertainty in both cohorts, both cis-PWAS and trans-PWAS showed high concordance between UKB-PPP and deCODE (slope = 1.16 and 1.01, respectively). As expected, cross-cohort concordance was attenuated when differences in protein level prediction accuracy were not accounted for (**Supplementary Figure 18**). Results were similar across PPC adjustments (**Supplementary Figure 19**). Concordance between UKB-PPP and the smaller CSF data set was lower, consistent with its reduced sample size and with tissue-specific proteomic regulation^32^ (**Supplementary Figure 20**).

**Figure 5.**
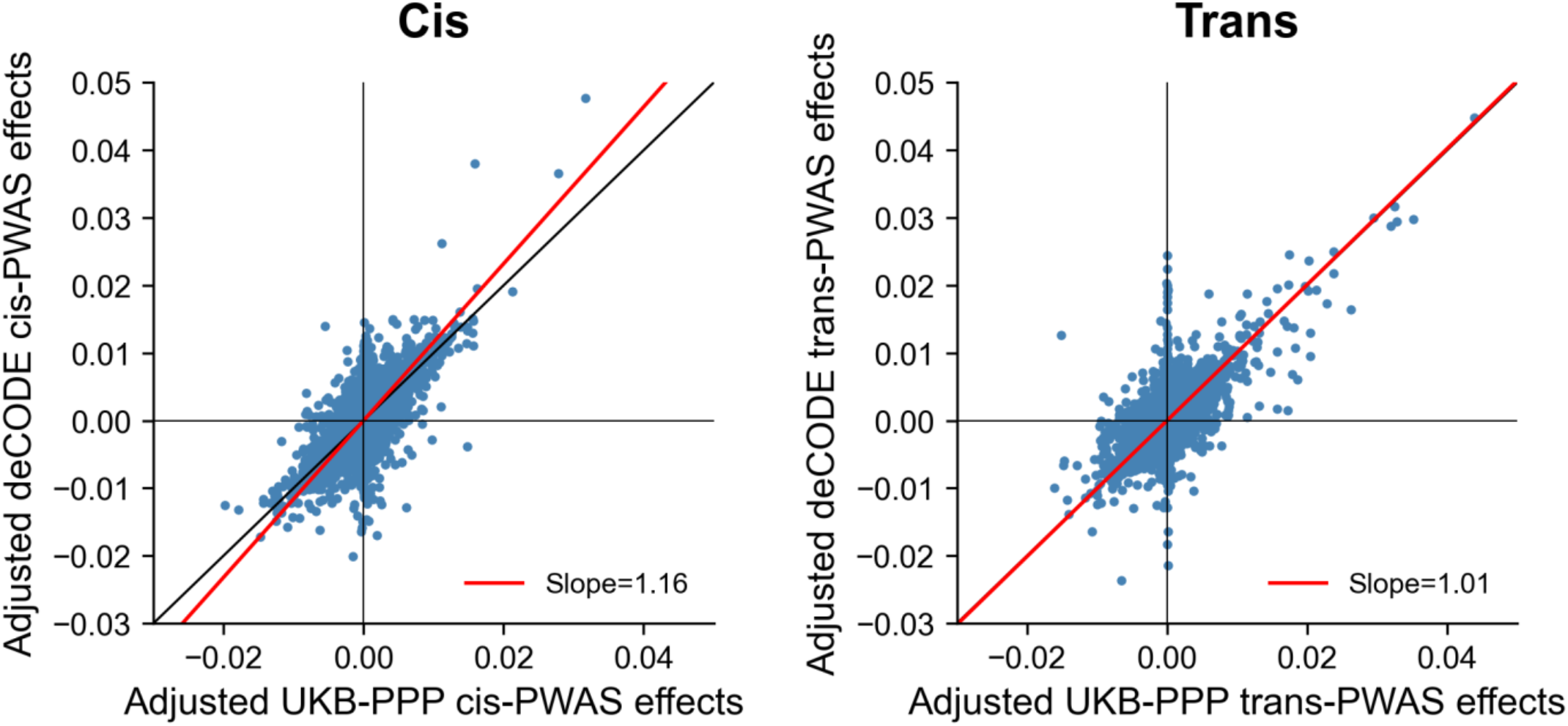
Cross-cohort concordance of PWAS associations between UKB-PPP and deCODE protein level cohorts. Adjusted PWAS effect sizes are compared between cohorts across 88 diseases/traits for 1,677 shared proteins, separately for cis-PWAS (left panel) and trans-PWAS (right panel). Each point denotes a protein-trait pair; the red line shows the Deming regression fit. Numerical results are reported in **Supplementary Table 9**.

We performed 5 secondary analyses using UKB-PPP data. First, we evaluated the heterogeneity of variance explained across trait categories: lipid and blood cell traits showed the highest values for both cis and trans, consistent with their strong representation among plasma proteins^58^ (**Supplementary Figure 21**). Second, we evaluated the impact of PPC adjustment on disease/trait variance explained by predicted protein levels. Adjusting for 20 PPCs had little effect on cis-explained variance but reduced trans-explained variance by 18% on average, with cross-trait variability consistent with how strongly trans-PPCs captured disease/trait variance (**Supplementary Figure 22**). Third, we evaluated the impact of PPC adjustment on PWAS associations. Adjusting for 20 PPCs reduced significant trans-PWAS associations by 44% on average, while cis-PWAS associations were largely unchanged (**Supplementary Figure 23**). Fourth, we partitioned cis-predicted protein levels into proximal and non-proximal components; both explained similar proportions of disease/trait variance, indicating that proximal variants—despite potential platform-specific epitope effects^55–57^—still capture disease-relevant biological variance (**Supplementary Figure 24**). In some cases, proximal and non-proximal components showed distinct effects, which may reflect differential tagging of causal cell types and contexts^80–82^ (**Supplementary Figure 25**). Fifth, we applied MESC^13^ to cis-pQTL data to estimate disease heritability mediated by cis-pQTL (**Supplementary Figure 26**). Median MESC estimates across 32 UK Biobank diseases/traits were 4.1%, lower than the 9.6% variance explained by cis-predicted protein levels, consistent with these methods capturing different quantities^13,51^ (**Methods**). Applying MESC to trans-pQTL data would likely produce strongly downward biased results at current sample sizes^83^.

### Validation of genes prioritized by cis- and trans-PWAS associations

We validated PWAS-prioritized genes using two complementary validation gene sets: a gold-standard set of genes identified by rare variant association studies^44^ (RVAS) and a silver-standard set of genes prioritized by PoPS network-based scores^45^ (we note that PoPS does not incorporate eQTL/pQTL data and thus provides an independent validation). We focused these validations on UKB-PPP protein levels and on 32 UK Biobank diseases/traits, because RVAS require large-scale rare variant data. For each trait, we selected the top PoPS-ranked genes equal in number to RVAS genes (mean 5.9 genes per trait). These two gene sets were largely non-overlapping, sharing 0.7 genes per disease/trait on average (excess overlap = 73, 95% CI: 46-115).

We assessed the overlap between PWAS-prioritized genes with RVAS/PoPS validation gene sets by jointly stratifying genes by both cis- and trans-PWAS Z-scores (**Figure 6a-b** and **Supplementary Table 10**). We reached two main conclusions. First, genes with higher cis-PWAS Z-scores showed increased excess overlap with both RVAS genes and PoPS genes; for example, excess overlap with RVAS genes increased from 6.4 for genes with |Z_cis_| = 5-10 (95% CI: [4.5, 9.0]) to 40 for genes with |Z_cis_| ≥10 (95% CI: [29, 55]). Second, within a given stratum of cis-PWAS Z-score, genes with higher trans-PWAS Z-scores showed increased excess overlap with both RVAS and PoPS genes. For example, excess overlap with RVAS genes rose to 76 for genes with both |Z_cis_| ≥10 and |Z_trans_| ≥10 (95% CI: [50, 115]; *P*=0.015 vs. 40 for genes with |Z_cis_| ≥10), highlighting the added value of trans-PWAS for gene prioritization. Notably, when considered independently of cis evidence, trans-PWAS showed lower enrichment than cis-PWAS, likely reflecting indirect tagging effects (**Supplementary Figure 27**).

**Figure 6.**
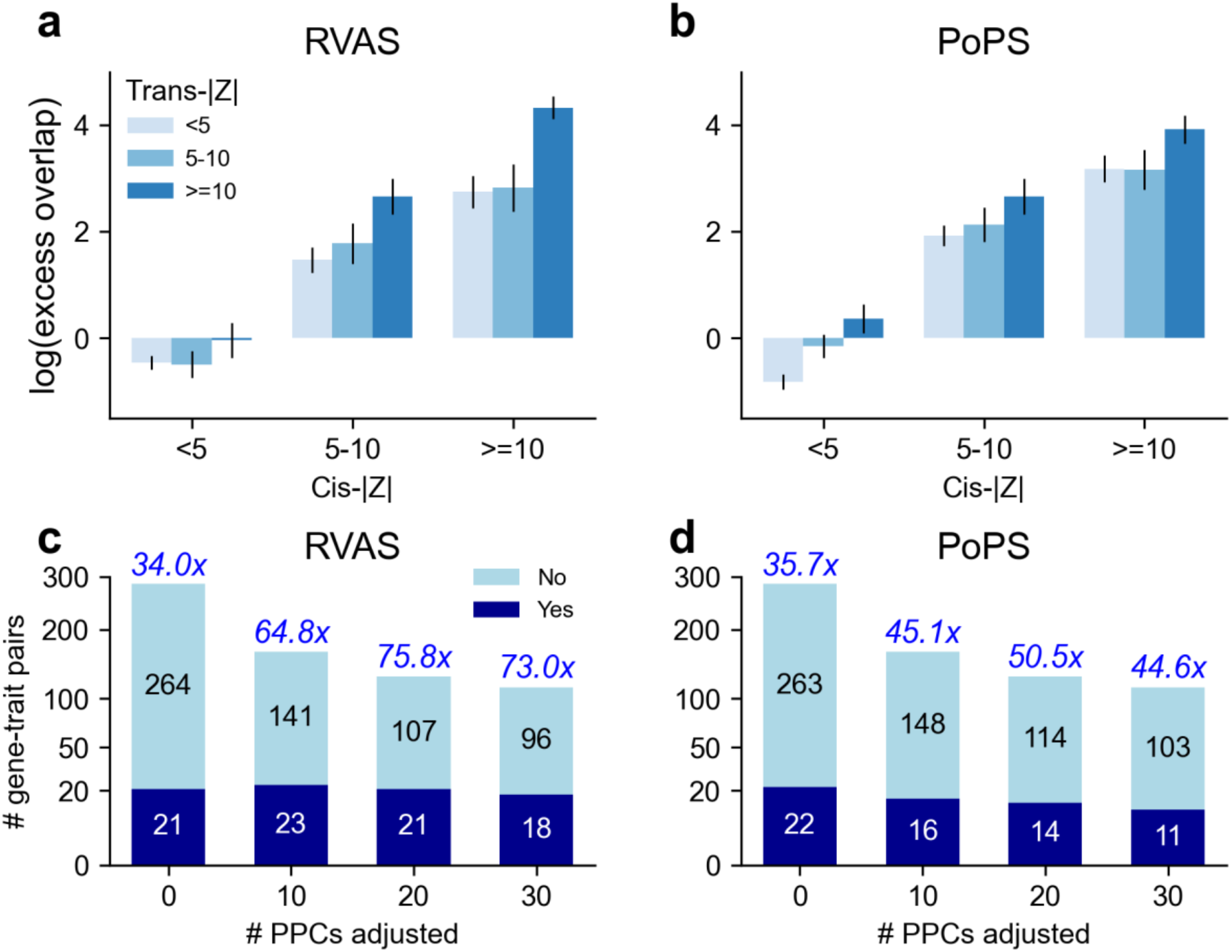
Validating protein-disease associations using RVAS and PoPS validation gene sets. **(a-b)** Excess overlap of PolyPWAS-prioritized genes for RVAS and PoPS validation gene sets, stratified jointly by absolute cis-PWAS and trans-PWAS Z-scores (|Z| < 5, 5 ≤ |Z| < 10, |Z| ≥ 10). **(c-d)** Enrichment of genes with strong cis and trans support (|Z_cis_| ≥10 and |Z_trans_| ≥10) for RVAS and PoPS validation gene sets as a function of #PPCs adjusted. Dark bars (resp. light bars) denote gene-trait associations overlapping (resp. not overlapping) validation gene sets. Numbers above each bar denote fold-enrichment relative to all gene-trait pairs. Numerical results are reported in **Supplementary Table 10**.

We next evaluated the impact of PPC adjustment on validation of genes with strong cis and trans support (|Z_cis_| ≥10 and |Z_trans_| ≥10). Adjusting for PPCs (which reduces co-regulation, **Figure 2d**) greatly reduced the number of prioritized PWAS associations but only slightly reduced the number of validated associations (**Figure 6c-d, Supplementary Figure 23** and **Supplementary Table 11**). For example, 21 of 128 gene-trait pairs prioritized after adjusting for 20 PPCs overlapped with RVAS genes, versus 21 of 285 when no PPCs were adjusted. Adjusting for 20 PPCs largely produces a subset of unadjusted results: among 108 gene-trait pairs prioritized under both conditions, 19 overlapped with RVAS genes. Thus, adjusting for PPCs improves specificity while preserving true positive associations.

To quantitatively assess the predictive value of cis- and trans-PWAS for gene prioritization, we modeled RVAS and PoPS validation gene set membership via logistic regression, using as features 4 bins of PWAS Z-scores (|Z| < 5, 5 ≤ |Z| < 10, 10 ≤ |Z| < 15, |Z| ≥ 15) for both cis and trans effects respectively (**Figure 7, Supplementary Table 11** and **Methods**). Including both cis and trans features outperformed cis-only models for both RVAS (pseudo-*R*^2^=11.6% vs. 10.5%, +11% relative improvement, *P=*1.8×10^-6^ for difference) and PoPS (pseudo-*R*^2^=14.0% vs. 13.0%, +7.0% relative improvement, *P=*1.9×10^-5^ for difference). Adding 4 bins of cis/trans coregulation scores further improved performance for both RVAS (pseudo-*R*^2^=14.3% vs. 11.6%, +23% further relative improvement, *P=*6.9×10^-8^ for difference; +36% total relative improvement, *P=*8.3×10^-18^ for difference) and PoPS (pseudo-*R*^2^=14.6% vs. 14.0%, +4.3% further relative improvement, *P=*0.14 for difference; +11% total relative improvement, *P=*1.3×10^-7^ for difference) (**Supplementary Figure 28**). In most cases, co-regulation features received negative coefficients, consistent with associations for proteins with high co-regulation scores often reflecting tagging associations rather than causal mediation^9,10,15–17^. These findings motivate future gene-level fine-mapping approaches^10,15–17^ incorporating both cis and trans associations (**Discussion**). We confirmed that validation performance improved with increasing the number of PPCs for RVAS (pseudo-*R*^*2*^ from 10.3% to 11.6%) and remained stable for PoPS (14.2% to 14.0%) when increasing from 0 to 20 PPCs (**Supplementary Figure 29**).

**Figure 7.**
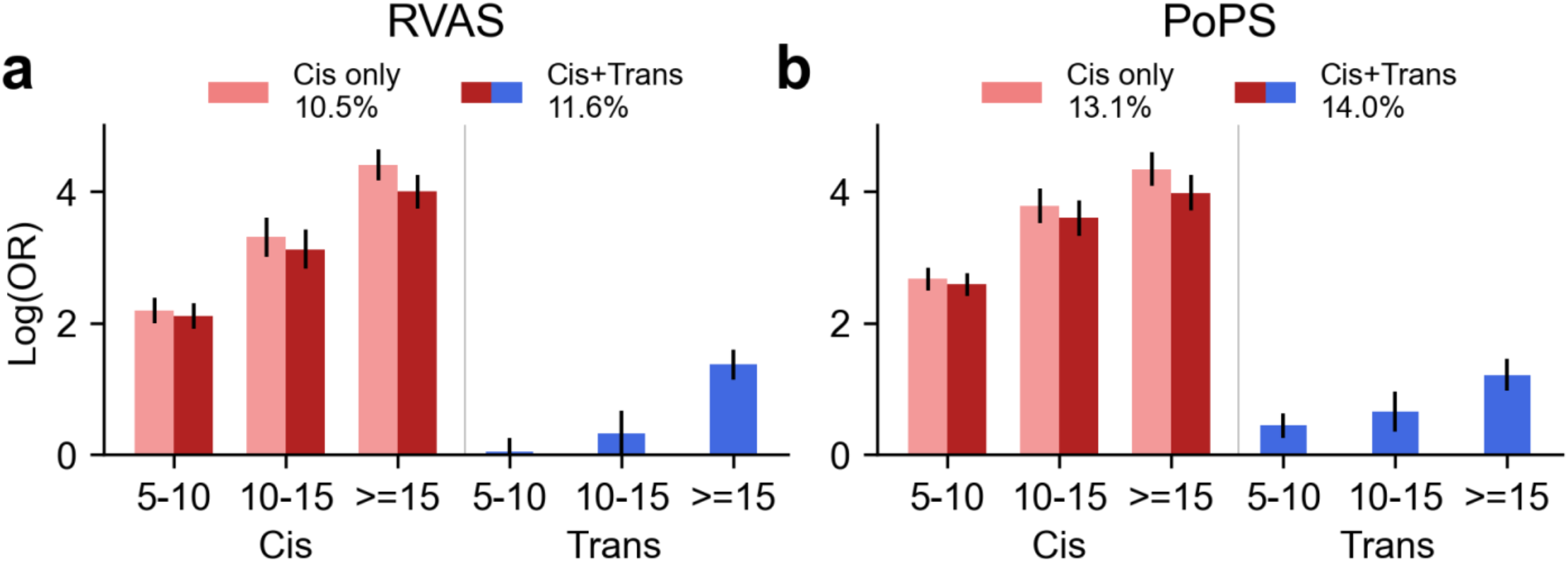
Predicting RVAS and PoPS validation gene set membership using cis- and trans-PWAS results. Logistic regression coefficients (log[OR]) for gene-trait pairs to be members of **(a)** RVAS or **(b)** PoPS gene sets, stratified by binned absolute cis-PWAS and trans-PWAS Z-scores (|Z|<5, 5≤|Z|<10, 10≤|Z|<15, |Z|≥15). Numerical results are reported in **Supplementary Table 11**.

We performed 8 secondary analyses. First, we assessed how our main findings (**Figure 7**) varied across disease/trait categories, for which we detected significant heterogeneity in how cis and trans-derived features predict validation gene sets (*P*=5.2×10^-75^ for RVAS; *P*=1.3×10^-42^ for PoPS; **Supplementary Figure 30**); per-category results were noisy for most trait categories, but blood cell traits (a relatively well-powered trait category) showed typical cis and trans validation performance, comparable to meta-analyzed results across all traits. Second, we repeated RVAS/PoPS validation with deCODE and CSF pQTL data, observing broadly similar patterns despite reduced power in CSF due to smaller sample size (**Supplementary Figure 31**). Third, we stratified cis-PWAS by proximal versus non-proximal variants in a joint model; both components had significant effects in predicting validation gene sets, and modeling both further improved validation performance (**Supplementary Figure 32**). Fourth, we included conditional (stepwise) Z-scores in a joint model with marginal Z-scores for both cis and trans; conditional features moderately improved validation performance, suggesting that fine-mapping approaches can potentially improve gene prioritization (**Supplementary Figure 33**). Fifth, we repeated RVAS/PoPS validation separately for gene-trait pairs with concordant versus discordant cis/trans sign, observing similar enrichment in validation gene sets for both (**Supplementary Figure 34**). Sixth, we evaluated effect-direction concordance at RVAS genes^84^, for which loss-of-function burden test results^44^ predict the PWAS effect direction. Among 79 RVAS gene-trait pairs that were significant in cis (P < 10^-5^) and a separate set of 79 RVAS gene-trait pairs that were significant in trans, cis-PWAS showed significant directional concordance with burden test results (79%, 62/79; binomial *P*=3.6×10^-7^), whereas trans-PWAS did not (57.0%, 45/79; *P*=0.26) (**Supplementary Figure 35**). This pattern is consistent with trans-PWAS often reflecting co-regulation/tagging effects, further emphasizing the need for gene-level fine-mapping approaches^10,15–17^ incorporating both cis and trans associations (**Discussion**). Seventh, to assess whether trans-PWAS results are driven by cell-type composition effects or highly pleiotropic pQTL hotspots, we filtered trans-pQTL weights by (i) removing blood cell trait GWAS SNPs (353K SNPs; P < 5×10^-8^ for any of 7 independent UK Biobank blood cell traits) or (ii) excluding pQTL hotspot SNPs (top 0.1% of SNPs by number of genome-wide significant pQTL associations at P < 5×10^-8^, ≥41 associated proteins, ∼7K SNPs); these two sets of SNPs are substantially overlapping: 91% of pQTL hotspot SNPs are also blood cell trait GWAS SNPs (**Supplementary Figure 36**). Removing blood cell GWAS SNPs reduced trait variance explained by trans-predicted protein levels by 43% (median across traits) and significant trans-PWAS associations by 54%, while removing hotspot SNPs had a more modest effect (10% and 22%, respectively), with both filters showing larger reductions for blood cell traits (**Supplementary Figure 37**). Among top-prioritized genes (cis-|Z| > 10 and trans-|Z| > 10, using 20 PPCs), both filters increased RVAS enrichment but reduced the number of prioritized pairs; as a result, neither improved overall validation pseudo-*R*^*2*^ (**Supplementary Figure 38**). We also stratified our main validation analysis by the 7 blood cell and 25 non-blood cell traits. The 25 non-blood cell traits remained well-powered and showed similar RVAS/PoPS enrichment and pseudo-*R*^*2*^ as results for all 32 traits (**Supplementary Figure 38**; also see **Supplementary Figure 30**), confirming that our main conclusions are not driven by cell-type composition effects. Eighth, we assessed reciprocal predictive power between PoPS and RVAS validation gene sets. Adding PoPS-prioritized genes as predictive features improved performance on RVAS validation gene sets (pseudo-*R*^*2*^ = 15.7% vs. 11.6%, *P* = 1.6×10^-25^ for difference) and adding RVAS-prioritized genes improved performance on PoPS validation gene sets (pseudo-*R*^*2*^ = 14.6% vs. 14.0%, *P* = 1.3×10^-3^ for difference) (**Supplementary Figure 39**).

### Combining cis- and trans-PWAS associations connects disease-critical genes

We highlight three examples in which PolyPWAS combines cis- and trans-PWAS associations to connect disease-critical genes that were each previously implicated in GWAS analyses (**Figure 8a**). Each example includes a trans-pQTL for a focal gene (with a direct effect on disease/trait) that is likely a cis-QTL for a gene mediating the trans-pQTL (cis-gene)^61,85^. The identity of the cis-gene is determined using locus-to-gene assignment^86^ instead of cis-pQTL (**Methods**), because most candidate cis-genes are unassayed across the proteomic platforms that we analyzed. In some cases the cis-gene has an independent direct effect on the disease/trait, which may lead to discordant (marginal) cis-PWAS and trans-PWAS effects for the focal gene due to tagging (**Supplementary Figure 35**). Because cis co-regulation reflects shared local regulation while trans co-regulation reflects shared distal regulation, concordant cis and trans associations can provide a stronger basis for gene prioritization.

**Figure 8.**
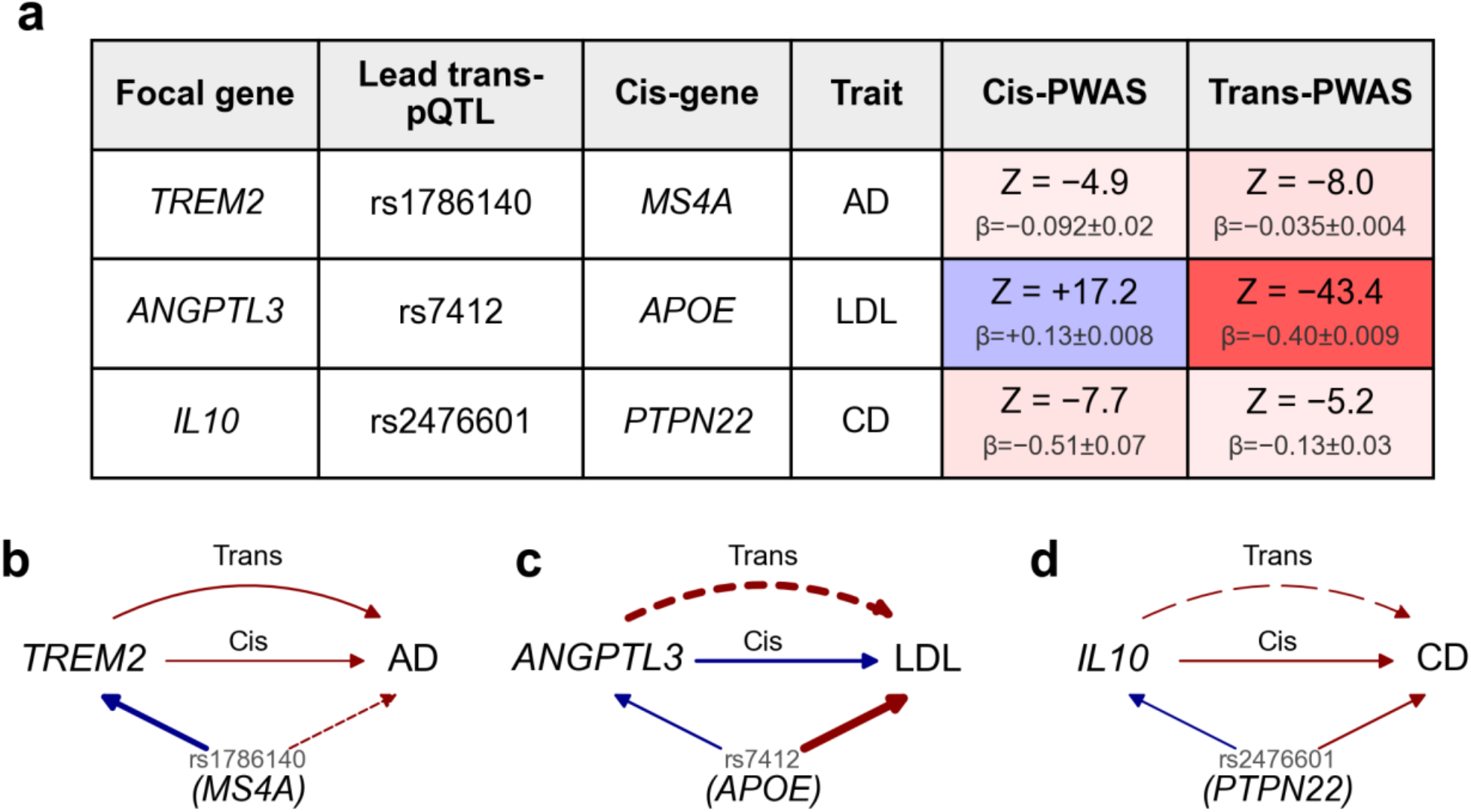
Combining cis- and trans-PWAS associations connects disease-critical genes. **(a)** Summary statistics for three selected examples, including Z-scores, effect sizes, and standard errors for cis and trans effects, and focal gene, lead trans-pQTL, cis-gene. Cis and trans effects are color-shaded by effect direction (blue: positive, red: negative) and Z-score magnitudes; the ratio between cis-Z and trans-Z differs from the ratio between cis-β, trans-β due to differences in standard errors arising from differences between cis protein prediction vs. trans protein prediction accuracy. **(b-d)** Diagrams of three focal gene-trait pairs with both cis- and trans-PWAS associations (*TREM2*-AD, *ANGPTL3*-LDL, *IL10*-CD). The focal gene and trait are shown as nodes on the left and right, respectively; edges between them denote cis-PWAS (straight) and trans-PWAS (curved) Z-scores. The cis-gene node at the bottom shows the lead trans-pQTL SNP (above) and the inferred cis-gene (below); arrows from the cis-gene to the focal gene and trait denote the pQTL and GWAS Z-scores, respectively. Edge color denotes effect direction (blue: positive, red: negative) and edge width denotes Z-score magnitude. Edge style denotes the degree of causal evidence: solid edges indicate direct effects; dashed edges indicate likely tagging or indirect effects.

First, we highlight an example involving concordant cis-PWAS and trans-PWAS effects, where the trans-PWAS effect reflects a likely causal relationship (**Figure 8b**). PolyPWAS associated *TREM2* with Alzheimer’s disease (AD) (cis-Z=−4.9; trans-Z=−8.0). *TREM2* encodes a microglial surface receptor^87^; rare coding variants substantially increase AD risk^88,89^. The negative cis-PWAS association is directionally consistent with this prior evidence. The negative trans-PWAS association was primarily driven by a non-coding variant in the *MS4A* gene cluster locus (rs1786140-C; trans-Z induced by rs1786140-C = −7.0; C allele lowers *MS4A4A* and *MS4A6A* expression in GTEx whole blood; **Methods**), an established AD GWAS locus^90^ embedded among other trans-pQTLs (**Supplementary Figure 40**). The concordant negative cis and trans PWAS effects (−0.092 and −0.040; differences arise from different prediction accuracies; **Methods**) support a likely causal trans-PWAS association in which *TREM2* mediates genetic effects at the *MS4A* locus on AD risk.

Second, we highlight an example with discordant cis-PWAS and trans-PWAS effects, with the trans effect reflecting a combination of likely causal and tagging effects (**Figure 8c**). PolyPWAS associated *ANGPTL3* with LDL cholesterol (cis-Z=+17.2; trans-Z=−43.4). *ANGPTL3* is a hepatocyte-secreted inhibitor of lipoprotein lipase^91^; reduced *ANGPTL3* levels lower LDL cholesterol and cardiovascular risk^92,93^. The positive cis-PWAS association is consistent with this prior evidence. The strong negative trans association was primarily driven by an *APOE* coding variant (rs7412-T; trans-Z induced by rs7412-T = −103), an established LDL GWAS locus embedded among other trans-pQTLs (**Supplementary Figure 41**). Because *APOE* is assayed in UKB-PPP, we were able to verify the locus-to-gene assignment: rs7412-T is a cis-pQTL for APOE (pQTL Z = 54), and the *APOE* cis-PWAS Z-score for LDL (−77) is concordant with the GWAS Z for rs7412-T (−103). This variant increases *ANGPTL3* levels but also has a direct LDL-lowering effect through altered *APOE*-dependent lipoprotein clearance^94^. Because the direct *APOE*-driven LDL effect opposes the indirect LDL-increasing effect mediated through higher *ANGPTL3* levels, the resulting trans association for *ANGPTL3* is discordant in sign and largely reflects tagging of *APOE*. Despite the discordant signs, the strong cis and trans associations indicate that *ANGPTL3* lies centrally within LDL regulatory networks; according to our validation analyses (**Figures 6-7**), such genes are more likely to be critical for disease.

Third, we highlight an example with partially concordant cis-PWAS and trans-PWAS effects, with the trans effect reflecting a combination of likely causal and tagging effects (**Figure 8d**). PolyPWAS associated *IL10* with Crohn’s disease (CD) (cis-Z=−7.7; trans-Z=−5.2). *IL10* is an anti-inflammatory cytokine^95^; rare loss-of-function variants in *IL10* cause severe early-onset inflammatory bowel disease^96^. The negative cis association is consistent with this prior evidence. The negative trans-PWAS association was partly driven by a *PTPN22* missense variant (rs2476601-T; trans-Z induced by rs2476601-T = −8.1; rs2476601-T is not a significant eQTL for *PTPN22* in GTEx immune-relevant tissues, suggesting its effects on *IL10* levels and CD risk may operate through altered protein function rather than transcription regulation); rs2476601-T is at a pleiotropic autoimmune susceptibility locus^97^ embedded among other trans-pQTLs (**Supplementary Figure 42**). This variant increases *IL10* levels (which is expected to decrease CD risk), but also independently decreases CD risk through additional pathways^98^, so the *IL10* trans-PWAS effect partly reflects tagging of *PTPN22*. Intriguingly, a missense variant in *IL10RA* (rs3135932) is a strong pQTL for *IL10* but shows no significant association with CD, despite the severe phenotypes caused by *IL10RA* loss-of-function mutations^99^—possibly reflecting compensatory feedback within *IL10* signaling, whereby modest reductions in *IL10RA* signaling trigger increased IL-10 production to preserve downstream anti-inflammatory flux and buffer Crohn’s disease risk^95^. Thus, the *IL10* trans-PWAS association aggregates multiple trans-pQTLs with distinct relationships to CD. Despite this complexity, the convergent cis-PWAS and trans-PWAS associations indicate that *IL10* is a central immune regulator in CD.

Additional examples are reported in **Supplementary Figure 43-45**. Overall, these examples show that trans-PWAS associations can reflect causal and/or tagging effects, and that combining cis-PWAS and trans-PWAS associations can help distinguish between causal vs. tagging effects in gene regulatory networks.

## Discussion

We have developed PolyPWAS, a functionally informed PWAS approach that combines cis- and trans-regulatory information to prioritize disease genes. Incorporating functional enrichment (96 baseline-LD model annotations^42^ and proteome-wide pleiotropy) substantially improves trans-prediction accuracy. Unlike previous methods leveraging cis- and trans-molecular QTL^37–39^, PolyPWAS uses polygenic prediction^41^ to model polygenic regulatory effects instead of restricting to genome-wide significant loci; analyzes cis and trans components separately; and operates entirely on pQTL and GWAS summary statistics. Trans-predicted protein levels explain roughly twice as much disease/trait heritability as cis-predicted levels. Integrating cis and trans evidence improved gene prioritization, attaining higher concordance with RVAS and PoPS gene sets than cis-only analyses; adjusting for PPCs reflecting broadly shared regulatory modules improved disease gene specificity. Overall, trans-predicted protein levels provide substantial complementary information. Importantly, in the examples that we have highlighted, trans-PWAS associations can reflect either direct causal effects (concordant cis/trans associations) or tagging effects (discordant cis/trans) in gene regulatory networks.

Our results have several implications for downstream analyses. First, integrating cis-PWAS and trans-PWAS should be broadly prioritized for gene prioritization, because joint models outperform cis-only models in validation analyses using RVAS and PoPS benchmarks (**Figures 6-7**); cis-PWAS and trans-PWAS could also be combined with other gene prioritization resources and modalities^86,100–102^. Second, we have publicly released genome-wide protein prediction weights for immediate integration with GWAS summary statistics without further pQTL analysis. Third, trans-PWAS will become more powerful and impart a greater contribution to gene prioritization as much larger pQTL datasets become available (e.g. *N*=300K individuals and 5.4K proteins available in UK Biobank in 2027^103^). Fourth, although convergent cis-PWAS and trans-PWAS evidence provides the most confident gene prioritization, PolyPWAS can detect protein-disease associations that cis-only analyses may miss, particularly for evolutionarily constrained genes with low cis-heritability^18–20^; incorporating information about evolutionary constraint may help prioritize causal genes implicated by trans-PWAS but not cis-PWAS.

We note several limitations of this study. First, we focused primarily on plasma proteins measured in UKB-PPP, analyzing only 2.8K proteins. Although key findings replicated in both a deCODE plasma pQTL dataset and a smaller CSF pQTL dataset, analysis of larger multi-tissue proteomic datasets would improve resolution^104^. In addition, we did not analyze trans-eQTL data; however, PolyPWAS could be applied to perform trans-TWAS as genome-wide trans-eQTL summary statistics in large samples become available^85^. Second, trans-PWAS power was limited by low trans prediction accuracy (**Figure 2a**), which is far below trans-SNP heritability at current pQTL sample sizes; much larger pQTL datasets will help address this. Third, PWAS associations remain susceptible to tagging effects from protein co-regulation^9,10,15–17^ (especially in trans) or non-mediated causal variant effects^15,17,49,50^; in addition, PWAS (or TWAS) results may not reflect variant-level correlation between pQTL (or eQTL) and disease effects (**Methods**). We corrected for PCs of predicted protein levels (**Figure 2d**) (and also performed conditional analyses), but rigorously distinguishing causal from tagging associations will require formal fine-mapping methods^10,15–17^ that jointly model mediated and non-mediated components^15,17,50^; developing such methods that incorporate both cis and trans associations is a key priority. Indeed, incorporating features derived from cis/trans co-regulation scores substantially improved prediction of validation gene sets (**Supplementary Figure 28**), underscoring the promise of a more rigorous approach. Existing cis fine-mapping methods^10,15–17^ could also be applied to cis-PWAS, but we did not pursue this here given limited proteomic coverage; this will become increasingly valuable as proteomic coverage expands. Fourth, co-regulation effects impacting trans-PWAS may partly reflect variation in cell-type composition^23,24,27^. However, filtering variants associated with blood cell traits did not substantially improve enrichment in validation gene sets (**Supplementary Figure 37** and **Supplementary Figure 38**). Fifth, correcting for PCs of predicted protein levels reduces tagging associations driven by protein co-regulation (e.g., due to shared upstream regulators), but may also reduce power for causal proteins with shared upstream regulators. However, PPC adjustment improved gene prioritization (**Figure 6**), suggesting that the gain in specificity outweighs the loss in power. Sixth, to avoid bias from sample overlap, we excluded UKB-PPP participants from GWAS analyses^47^; explicitly modeling sample overlap could increase power. Seventh, PWAS is complementary to colocalization-based approaches, analogous to TWAS; integrating advantages of both frameworks remains an important direction^105,106^. Eighth, although the association step (Equation 1) is fast, SBayesRC^41^ is computationally intensive (**Supplementary Table 12**); restricting to HapMap3 SNPs reduces runtime (from 6 hours to 1 hour per protein; **Supplementary Table 12**) with a modest prediction accuracy loss, and is a plausible option when computational resources are limited. Ninth, our functional priors do not exhaustively incorporate variant- and gene-level annotations; expanding to network- or pathway-informed features^24,51^ may further improve trans prediction accuracy. Tenth, we did not consider other validation gene sets such as drug target^107,108^ and Mendelian^109^ disease-associated genes, even though they have been proposed as benchmarks for disease relevance^24,108^ and could inform integrated scores for drug target prioritization^100^. Eleventh, we treat each cis- or trans-predicted protein level as a single aggregate quantity, even though components of prediction from different variant classes can show distinct patterns of association to disease (**Supplementary Figure 25**). Indeed, performing cis-PWAS separately using proximal and non-proximal components improved prediction of validation gene sets (**Supplementary Figure 32**), suggesting that partitioning both cis and trans predictions by different variant classes could be a valuable future direction. Twelfth, we have not formally quantified the extent to which trans-pQTL effects are mediated by cis-pQTL^61,85^, which remains a future direction. Despite these limitations, our results show that trans-predicted protein levels explain substantial disease/trait heritability such that trans-PWAS complements cis-PWAS, and that jointly modeling cis and trans architectures is crucial for effectively prioritizing protein-mediated disease biology.

## Acknowledgements

We thank Hilary Finucane for sharing PoPS results; we thank Gaspard Kerner, Luke O’Connor, Alexander Gusev, Benjamin Neale, Jordan Rossen, Joshua Popp for helpful discussions and feedback at various stages of this project. This research was conducted using the UK Biobank Resource under Application #16549. This research was funded by NIH grants U01 HG012009, R01 MH101244, R37 MH107649, R01 HG006399, R01 MH115676.

## Methods

### PolyPWAS methods

#### Overview

PolyPWAS associates genetically predicted protein levels with diseases using both cis-(±1 Mb from transcription start site) and trans-(all remaining genome-wide) variants. First, we infer genome-wide SNP effects on protein levels from pQTL summary statistics using SBayesRC^41^. Second, we test associations between cis- and trans-genetically predicted protein levels and disease. For each protein, we partition SBayesRC-derived prediction weights 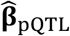 into cis 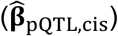 and trans 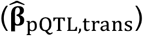, and compute cis and trans PWAS association Z-scores by correlating genetically predicted protein levels with disease risk, accounting for linkage disequilibrium (LD)^2^:

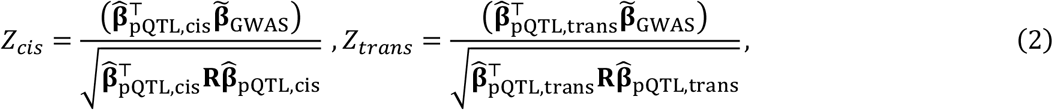

where 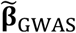 denotes marginal GWAS effects, and **R** is the LD correlation matrix.

We caution that due to LD, PWAS (or TWAS) results may not reflect variant-level correlation between pQTL (or eQTL) and disease effects^15,17,49,50^. In detail, PWAS tests for correlations between predicted protein levels and disease across individuals, which may be different from the correlation between disease and pQTL effect sizes across variants due to LD. For example, if a pQTL variant and a disease-causal variant are in perfect LD, predicted protein levels will correlate perfectly with disease risk across individuals—yet variant-level correlation between pQTL and disease effect sizes is zero, since neither variant carry both effects. Despite these caveats, our disease gene prioritization results confirm the utility of PWAS based on individual-level correlations (which can be efficiently computed from GWAS and pQTL summary statistics without requiring individual-level data).

#### Quantifying co-regulation across proteins

We computed co-regulation matrix 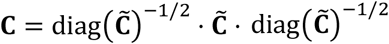, where 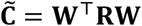 is the covariance of predicted protein levels, **W** is standardized SNP weights across proteins, computed separately for cis and trans. For each protein *i*, we defined its co-regulation score as 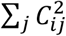 (summing over all protein *j*, including *i*). This score measures how strongly a protein’s genetic prediction correlates with predicted levels of other proteins. Higher co-regulation scores indicate that proteins are more susceptible to tagging associations^9,51^. We note that ref. ^51^ computed co-regulation scores with bias-corrected self-correlation *C*_*ii*_ ; we omit this correction because the bias is negligible at large sample sizes for cis co-regulation scores, and the self-correlation term is negligible for trans co-regulation scores. Co-regulation scores that we report are conservative estimates because current pQTL datasets profile only a fraction of all proteins.

#### Adjusting PWAS associations via predicted protein principal components

We performed conditional PWAS analyses using summary statistics^110^ to remove shared regulation from marginal PWAS associations. Let **W** ∈ ℝ^*N*×*P*^ denote standardized genetically predicted protein levels, **y** ∈ ℝ^*N*^ the phenotype, 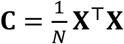 the protein co-regulation matrix, and 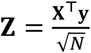 the marginal PWAS Z-scores. For a linear combination **s** = **Ww**, the marginal association is 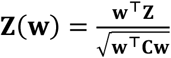. To adjust protein *i* for **s** , we regress **W**_*i*_ on **s** and test the residual, producing the conditional Z-score 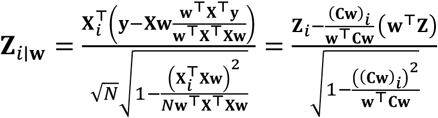 , which depends only on summary-level quantities **Z** and **C**. More generally, for multiple linear combinations **S** = **WW** where **W** ∈ ℝ^*P*×*K*^ with *K* columns **w**_1_ , … , **w**_*K*_ , adjusting protein *i* for all columns of **W** produces **Z** 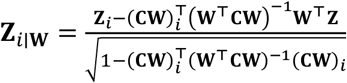. This formulation allows for adjusting for predicted protein principal components (PPCs) or for stepwise conditional analysis without requiring individual-level data. Specifically, we account for PPCs separately for cis and trans. Let **C**= **UΛU**^⊤^ denote the eigendecomposition of the protein co-regulation matrix. To adjust protein *i* for the top *K* principal components (**WU**_*K*_, with eigenvectors **W** = **U**_*K*_ and eigenvalues λ_1_, … , λ_*K*_), the PPC-adjusted Z-score for protein *i* is: 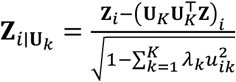. In our primary analysis, we adjusted for 20 PPCs.

### pQTL data processing and protein prediction

#### Proteogenomic resources and pQTL mapping

We analyzed three pQTL datasets: (i) UK Biobank Pharma Proteomics Project (UKB-PPP)^23^, comprising 54,219 individuals across 2,923 Olink assays in plasma; (ii) deCODE^31^, comprising 35,559 individuals with 4,907 SomaScan aptamers in plasma; and (iii) a cerebrospinal fluid (CSF) pQTL study^32^, comprising 3,506 individuals with 6,361 SomaScan aptamers in CSF. For UKB-PPP, we performed pQTL mapping using PLINK2^111^ in unrelated individuals of white British ancestry (N = 33,350), regressing inverse-rank normal transformed protein levels on genotypes with covariates for age, sex, and the top 10 genetic principal components. For downstream analyses that do not involve individual-level data, we used publicly pQTL summary statistics^23^. For deCODE and CSF, we used publicly released pQTL summary statistics. We used two LD reference panels previously computed from 20,000 randomly sampled unrelated UK Biobank individuals of European ancestry^41^: a HapMap3-based panel (1.2 million variants) and a dense well-imputed panel (7.7 million variants). Our primary analyses used the dense panel; we restricted to HapMap3 SNPs for compatibility with publicly available GWAS.

#### Functionally informed protein prediction

We trained per-protein genome-wide predictors using SBayesRC, which takes genome-wide pQTL summary statistics, reference LD, and variant-level annotations. The annotation set comprised: (i) 96 baseline-LD v2.2 annotations^42^; (ii) a binary cis-region indicator marking variants within ±1 Mb of the transcription start site; and (iii) proteome-wide pQTL pleiotropy annotations, where each variant was assigned to one of three annotations (or no annotation) based on whether it was a significant cis-pQTL (±1 Mb of gene), trans-pQTL (outside ±1 Mb), or both, across proteins in each dataset. We used SBayesRC posterior mean effects as prediction weights. For each protein, we partitioned weights into cis and trans components.

#### Evaluating protein prediction accuracy

We evaluated prediction accuracy as *R*^*2*^ between genetically predicted and covariate-adjusted measured protein levels in 9,746 held-out UKB-PPP individuals not used for pQTL mapping and prediction model training (i.e., not in the sample of pQTL mapping individuals; see above). We regressed out the top 20 PPCs (computed via PCA on standardized predicted proteins, separately for cis and trans) and evaluated *R*^*2*^ on the residuals.

#### Evaluating cross-cohort portability

We evaluated cross-cohort portability by applying deCODE and CSF prediction weights to held-out UKB-PPP individuals and computed *R*^*2*^ for proteins assayed in both UKB-PPP and deCODE, or in both UKB-PPP and CSF, matched by UniProt ID. We estimated cross-cohort genetic correlations for UKB-PPP vs. deCODE, and UKB-PPP vs. CSF. We estimated cis genetic correlations using Pearson correlations between cis-predicted protein levels in held-out individuals. We estimated trans genetic correlations using cross-trait LDSC^112^ with 1,000 Genomes^113^ as the LD reference. We also compared cross-cohort prediction *R*^*2*^ differences with individual-level measured protein concordance between Olink and SomaScan platforms^55^.

### Simulations

We performed simulations to evaluate PolyPWAS type I error rate and power using LD matrices computed from European-ancestry UK Biobank individuals restricted to HapMap3 SNPs. For each pQTL replicate, we generated protein genetic effects by randomly selecting 1% of SNPs as causal and drawing standardized effect sizes from a standard normal distribution, rescaled to achieve target SNP-heritability 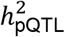. GWAS effects were simulated as a sum of protein-mediated and non-mediated components: β_GWAS_ = α β_pQTL_ + β_non-mediated_ , where 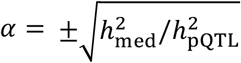 (with random sign per replicate) and β_non-mediated_ had 1% causal SNPs scaled to 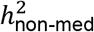. We varied mediated heritability 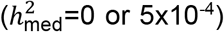 and non-mediated heritability 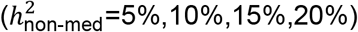. To evaluate tagging associations from shared genetic architecture, we simulated pairs of genetically correlated proteins (*r*_*g*_ = 0.2, 0.4, 0.6, 0.8) by drawing their causal effects from a bivariate normal distribution. Prediction weights were fitted on one protein and PWAS Z-scores were computed using GWAS effects mediated through the correlated protein. We generated pQTL summary statistics (*N*_*pQTL*_=40K) and GWAS Z-scores (*N*_*GWAS*_=300K) by adding multivariate normal noise under: 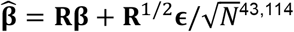, where **R**^1/2^ denotes the matrix square root of **R** obtained via eigen decomposition. For each simulated protein, we applied SBayesRC to the pQTL summary statistics using default parameters without functional annotation, obtained posterior mean prediction weights and computed PWAS Z-scores, assessing significance using a threshold of P < 10^-5^. We used a nested simulation for computational speedup: for each pQTL replicate, SBayesRC is fitted once to obtain prediction weights, and many independent GWAS replicates are then generated conditioned on those pQTL weights. To evaluate false positive rates, we simulated 1,000 pQTL replicates each with 20,000 GWAS replicates (20 million total tests per setting). To evaluate power and tagging rate, we simulated 100 pQTL replicates each with 100 GWAS replicates (10,000 total tests per setting).

### Associating proteins with GWAS diseases/traits

#### GWAS datasets

We analyzed 32 heritable, approximately independent UK Biobank disease and traits (**Supplementary Table 2**). We selected by filtering trait pairs at a squared genetic correlation threshold of 0.25 estimated via LD Score regression. We computed GWAS summary statistics using BOLT-LMM^115^ (v2.3.4), excluding UKB-PPP participants to avoid overlap between pQTL and GWAS samples. We also analyzed 56 additional diseases/traits with publicly available summary statistics (**Supplementary Table 2**), selected under the same independence criteria.

#### Variance explained by predicted protein levels

We estimated the proportion of trait SNP-heritability explained by predicted protein levels using a previously validated approach that estimates SNP-heritability explained by a set of SNPs^59^. For a set of predicted proteins (cis, trans, or combined), the proportion of trait variance explained can be estimated via 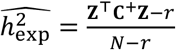, where **Z** are PWAS Z-scores, **C**^+^ is the pseudo-inverse of the protein co-regulation matrix, *r*is the rank of the protein prediction matrix, and *N* is the GWAS sample size. These quantities are computed from individual-level data. This is equivalent to the adjusted *R*^*2*^ from a multivariate regression of the trait on all predicted protein levels and is unbiased across sample size given that sample size is larger than the rank of protein prediction matrix (*N > r*). This estimator quantifies both mediated effects (disease variants acting through protein levels) and non-mediated effects (disease variants correlated with predicted protein levels but acting through other pathways)^51^. This contrasts with the Mediated Expression Score Regression (MESC)^13^ estimator, which explicitly partitions mediated from non-mediated components. We computed *h*^2^ for UK Biobank traits using individual-level phenotypes and predicted proteins in a held-out subset of 50K individuals not in UKB-PPP. We adjusted traits for covariates (sex, age, age^2^). Predicted protein levels were adjusted for top PPCs, computed separately for cis and trans. These estimates are conservative lower bounds because they reflect only currently profiled proteins and because measurement noise in trans-predicted levels introduces downward bias.

#### Conditionally independent associations

We identified conditionally independent protein-trait associations using stepwise forward regression of trait values against predicted protein levels. At each forward regression step, we computed the partial correlation of each remaining protein with the trait, conditioning on all previously selected proteins, using the co-regulation matrix and PWAS Z-scores. The protein with the largest absolute conditional Z-score was added to the model. Conditional Z-scores were computed from the summary-level partial correlation approach (see above). We estimated the number of conditionally independent associations by counting number of associations with P < 10^-5^.

#### Cross-cohort concordance of PWAS associations

We compared PWAS effects between UKB-PPP and deCODE or CSF across matched protein-trait pairs. Let *R*_*1*_ and *R*_*2*_ denote cohort-specific prediction accuracy after PPC adjustment, estimated in the same held-out UKB-PPP individuals for protein predictors from the two cohorts. For GWAS sample size *N*, and cohort-specific PWAS Z-scores *Z*_*1*_, *Z*_*2*_, we define scaled effect estimates 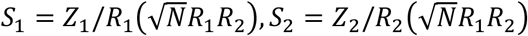 with standard errors 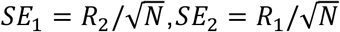, computed for cis and trans. This scaling allows them to be at the same effect scale up to the common multiplicative factor *R*_1_*R*_2_. We then fit Deming regression to *S*_1_and *S*_2_, accounting for uncertainty using *SE*_1_ and *SE*_2_.

#### Validation against RVAS and PoPS gene sets

We evaluated gene prioritization performance against two benchmark gene sets. First, we used rare variant association study (RVAS) genes prioritized by loss-of-function burden tests^44^, defining RVAS genes as those with burden P-values passing per-trait Bonferroni correction (P < 0.05/# genes with both PWAS and burden test results for that trait). Second, we used Polygenic Priority Score (PoPS)^45^, which integrates GWAS with gene co-expression and protein-protein interaction networks independent of pQTL data. For each trait, we labeled the top K genes by PoPS score as positives, where K equaled the number of RVAS genes for that trait. We stratified gene-trait pairs by bins of |Z_cis_| and |Z_trans_| (using bins: |Z| < 5, 5 ≤ |Z| < 10, 10 ≤ |Z| < 15, |Z| ≥ 15) and computed excess overlap within each stratum, defined as the ratio of observed to expected overlap with benchmark gene sets. We reported 95% confidence intervals for excess overlap based on binomial sampling. We additionally computed enrichment specifically among gene-trait pairs with |Z_cis_| > 10 and |Z_trans_| > 10 across different PPC adjustment. We fit logistic regression models to predict RVAS or PoPS membership using four nested feature sets: (1) binned |Z_cis_| alone; (2) binned |Z_cis_| and |Z_trans_|; (3) the preceding features plus cis co-regulation score quartiles; and (4) the preceding features plus trans co-regulation score quartiles. Z-scores were binned into 4 levels (|Z| < 5, 5 ≤ |Z| < 10, 10 ≤ |Z| < 15, |Z| ≥ 15); co-regulation scores were binned into quartiles. Model performance was quantified using pseudo-*R*^*2*^, computed as 1−(log-likelihood of fitted model / log-likelihood of null model). We assessed improvement from adding trans features to cis-only models, and from adding co-regulation features, via likelihood ratio tests comparing consecutive nested models. We additionally stratified validation models by trait categories and assessed cross-category heterogeneity by comparing the sum of per-trait log-likelihoods against the pooled model log-likelihood via a likelihood ratio test. To assess the complementarity of RVAS and PoPS, we added PoPS score quartiles as features to models predicting RVAS membership and conversely added burden Z-score quartiles to models predicting PoPS membership, with improvement assessed via pseudo-*R*^*2*^ and likelihood ratio tests.

#### Examples of PWAS associations

For examples of PWAS associations (**Figure 8**), we transformed PWAS Z-scores to standardized effect estimates by dividing by 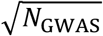 and the corresponding prediction correlation. Differences between cis and trans effect sizes are expected because of differences in prediction accuracy and because trans associations can include both causal and tagging effects. For each focal gene, we identified lead trans-pQTL loci by selecting the strongest pQTL per previously defined LD block (P < 10^-10^, excluding the cis region)^41^. The trans-PWAS Z-score driven by an individual lead SNP *i* is quantified as sign(*Z*_pQTL,*i*_) ⋅ *Z*_GWAS,*i*_. We annotated each lead SNP with its most likely linked gene (trans-pGene) using the Open Targets locus-to-gene (L2G) score^86^ (platform release 26.03), which integrates colocalization, chromatin interaction, and variant consequence data. We used L2G rather than direct pQTL evidence because trans-pGenes (e.g., *MS4A, PTPN22*) are often unassayed across the proteomics platforms we examined.

## Data availability

pQTL association statistics, protein prediction models, GWAS association statistics, PWAS association statistics, and validation metrics used in this study will be made publicly available upon publication.

## Code availability

PolyPWAS: https://github.com/kangchenghou/polypwas.

SBayesRC (v0.2.6): https://github.com/zhilizheng/SBayesRC.

PLINK2: https://www.cog-genomics.org/plink/2.0/.

BOLT-LMM: https://storage.googleapis.com/broad-alkesgroup-public/BOLT-LMM/BOLT-LMM_manual.html.

LDSC: https://github.com/bulik/LDSC.

## References

1. Gamazon, E. R. et al. A gene-based association method for mapping traits using reference transcriptome data. Nat. Genet. 47, 1091–1098 (2015).

2. Gusev, A. et al. Integrative approaches for large-scale transcriptome-wide association studies. Nat. Genet.48, 245–252 (2016).

3. Li, Y. I. et al. RNA splicing is a primary link between genetic variation and disease. Science 352, 600–604 (2016).

4. Chun, S. et al. Limited statistical evidence for shared genetic effects of eQTLs and autoimmune-disease-associated loci in three major immune-cell types. Nat. Genet. 49, 600–605 (2017).

5. Gusev, A. et al. Transcriptome-wide association study of schizophrenia and chromatin activity yields mechanistic disease insights. Nat. Genet. 50, 538–548 (2018).

6. Gandal, M. J. et al. Transcriptome-wide isoform-level dysregulation in ASD, schizophrenia, and bipolar disorder. Science 362, (2018).

7. Gamazon, E. R. et al. Using an atlas of gene regulation across 44 human tissues to inform complex disease-and trait-associated variation. Nat. Genet. 50, 956–967 (2018).

8. Barbeira, A. N. et al. Exploring the phenotypic consequences of tissue specific gene expression variation inferred from GWAS summary statistics. Nat. Commun. 9, 1825 (2018).

9. Wainberg, M. et al. Opportunities and challenges for transcriptome-wide association studies. Nat. Genet. 51, 592–599 (2019).

10. Mancuso, N. et al. Probabilistic fine-mapping of transcriptome-wide association studies. Nat. Genet. 51, 675–682 (2019).

11. Gamazon, E. R., Zwinderman, A. H., Cox, N. J., Denys, D. & Derks, E. M. Multi-tissue transcriptome analyses identify genetic mechanisms underlying neuropsychiatric traits. Nat. Genet. 51, 933–940 (2019).

12. GTEx Consortium. The GTEx Consortium atlas of genetic regulatory effects across human tissues. Science369, 1318–1330 (2020).

13. Yao, D. W., O’Connor, L. J., Price, A. L. & Gusev, A. Quantifying genetic effects on disease mediated by assayed gene expression levels. Nat. Genet. 52, 626–633 (2020).

14. Lappalainen, T., Li, Y. I., Ramachandran, S. & Gusev, A. Genetic and molecular architecture of complex traits. Cell 187, 1059–1075 (2024).

15. Zhao, S. et al. Adjusting for genetic confounders in transcriptome-wide association studies improves discovery of risk genes of complex traits. Nat. Genet. 56, 336–347 (2024).

16. Liu, L. et al. Conditional transcriptome-wide association study for fine-mapping candidate causal genes. Nat. Genet. 56, 348–356 (2024).

17. Strober, B. J., Zhang, M. J., Amariuta, T., Rossen, J. & Price, A. L. Fine-mapping causal tissues and genes at disease-associated loci. Nat. Genet. 57, 42–52 (2025).

18. Boyle, E. A., Li, Y. I. & Pritchard, J. K. An expanded view of complex traits: From polygenic to omnigenic. Cell 169, 1177–1186 (2017).

19. Liu, X., Li, Y. I. & Pritchard, J. K. Trans effects on gene expression can drive omnigenic inheritance. Cell 177, 1022–1034.e6 (2019).

20. Mostafavi, H., Spence, J. P., Naqvi, S. & Pritchard, J. K. Systematic differences in discovery of genetic effects on gene expression and complex traits. Nat. Genet. 55, 1866–1875 (2023).

21. Price, A. L. et al. Single-tissue and cross-tissue heritability of gene expression via identity-by-descent in related or unrelated individuals. PLoS Genet. 7, e1001317 (2011).

22. Grundberg, E. et al. Mapping cis-and trans-regulatory effects across multiple tissues in twins. Nat. Genet. 44, 1084–1089 (2012).

23. Sun, B. B. et al. Plasma proteomic associations with genetics and health in the UK Biobank. Nature 622, 329–338 (2023).

24. Li, J., Li, Y. I. & Liu, X. Protein-protein interactions shape trans-regulatory impact of genetic variation on protein expression and complex traits. Nat. Genet. 58, 77–87 (2026).

25. Liu, X. et al. Functional architectures of local and distal regulation of gene expression in multiple human tissues. Am. J. Hum. Genet. 100, 605–616 (2017).

26. Dutta, D. et al. Aggregative trans-eQTL analysis detects trait-specific target gene sets in whole blood. Nat. Commun. 13, 4323 (2022).

27. Võsa, U. et al. Large-scale cis-and trans-eQTL analyses identify thousands of genetic loci and polygenic scores that regulate blood gene expression. Nat. Genet. 1–11 (2021).

28. Zhao, J. H. et al. Genetics of circulating inflammatory proteins identifies drivers of immune-mediated disease risk and therapeutic targets. Nat. Immunol. 24, 1540–1551 (2023).

29. Wang, L., Babushkin, N., Liu, Z. & Liu, X. Trans-eQTL mapping in gene sets identifies network effects of genetic variants. Cell Genom. 4, 100538 (2024).

30. Sun, B. B. et al. Genomic atlas of the human plasma proteome. Nature 558, 73–79 (2018).

31. Ferkingstad, E. et al. Large-scale integration of the plasma proteome with genetics and disease. Nat. Genet. 53, 1712–1721 (2021).

32. Western, D. et al. Proteogenomic analysis of human cerebrospinal fluid identifies neurologically relevant regulation and implicates causal proteins for Alzheimer’s disease. Nat. Genet. 56, 2672–2684 (2024).

33. Puerta, R. et al. Human CSF proteogenomics links genetic variation to neurodegenerative disease proteins. medRxiv (2026) doi:10.64898/2026.02.12.26345733.

34. Iakovliev, A. et al. Genome-wide aggregated trans-effects on risk of type 1 diabetes: A test of the “omnigenic” sparse effector hypothesis of complex trait genetics. Am. J. Hum. Genet. 110, 913–926 (2023).

35. Wittich, H. et al. Transcriptome-wide association study of the plasma proteome reveals cis and trans regulatory mechanisms underlying complex traits. Am. J. Hum. Genet. 111, 445–455 (2024).

36. Fang, L., Xue, H., Lin, Z. & Pan, W. Multivariate proteome-wide association study to identify causal proteins for Alzheimer disease. Am. J. Hum. Genet. 112, 291–300 (2025).

37. Donoghue, L. J. et al. Integration of biobank-scale genetics and plasma proteomics reveals evidence for causal processes in asthma risk and heterogeneity. Cell Genom. 5, 100840 (2025).

38. Luningham, J. M. et al. Bayesian Genome-wide TWAS Method to Leverage both cis-and trans-eQTL Information through Summary Statistics. Am. J. Hum. Genet. 107, 714–726 (2020).

39. Head, S. T. et al. Cis-and trans-eQTL TWASs of breast and ovarian cancer identify more than 100 susceptibility genes in the BCAC and OCAC consortia. The American Journal of Human Genetics vol. 111 1084–1099 (2024).

40. Xu, Y. et al. An atlas of genetic scores to predict multi-omic traits. Nature 616, 123–131 (2023).

41. Zheng, Z. et al. Leveraging functional genomic annotations and genome coverage to improve polygenic prediction of complex traits within and between ancestries. Nat. Genet. 56, 767–777 (2024).

42. Gazal, S. et al. Linkage disequilibrium-dependent architecture of human complex traits shows action of negative selection. Nat. Genet. 49, 1421–1427 (2017).

43. Pasaniuc, B. & Price, A. L. Dissecting the genetics of complex traits using summary association statistics. Nat. Rev. Genet. 18, 117–127 (2017).

44. Backman, J. D. et al. Exome sequencing and analysis of 454,787 UK Biobank participants. Nature 599, 628–634 (2021).

45. Weeks, E. M. et al. Leveraging polygenic enrichments of gene features to predict genes underlying complex traits and diseases. Nat. Genet. 55, 1267–1276 (2023).

46. Finucane, H. K. et al. Partitioning heritability by functional annotation using genome-wide association summary statistics. Nat. Genet. 47, 1228–1235 (2015).

47. Loesch, D. P. et al. Identification of plasma proteomic markers underlying polygenic risk of type 2 diabetes and related comorbidities. Nat. Commun. 16, 2124 (2025).

48. The International HapMap 3 Consortium. Integrating common and rare genetic variation in diverse human populations. Nature 467, 52–58 (2010).

49. de Leeuw, C., Werme, J., Savage, J. E., Peyrot, W. J. & Posthuma, D. On the interpretation of transcriptome-wide association studies. PLoS Genet. 19, e1010921 (2023).

50. Liang, Y., Nyasimi, F. & Im, H. K. A gene-specific variance-control approach corrects polygenicity-driven inflation observed in transcriptome-wide association studies. Am. J. Hum. Genet. (2026) doi:10.1016/j.ajhg.2025.12.014.

51. Siewert-Rocks, K. M., Kim, S. S., Yao, D. W., Shi, H. & Price, A. L. Leveraging gene co-regulation to identify gene sets enriched for disease heritability. Am. J. Hum. Genet. 109, 393–404 (2022).

52. Emilsson, V. et al. Co-regulatory networks of human serum proteins link genetics to disease. Science 361, 769–773 (2018).

53. Pietzner, M. et al. Mapping the proteo-genomic convergence of human diseases. Science 374, eabj1541 (2021).

54. Dey, K. K. et al. SNP-to-gene linking strategies reveal contributions of enhancer-related and candidate master-regulator genes to autoimmune disease. Cell Genom. 2, 100145 (2022).

55. Eldjarn, G. H. et al. Large-scale plasma proteomics comparisons through genetics and disease associations.Nature 622, 348–358 (2023).

56. Nicholas, J. C. et al. Cross-ancestry comparison of aptamer and antibody protein measures. Nat. Commun. 17, 1054 (2026).

57. Kuliesius, J. et al. Epitope Effect Prevalence in Affinity-based pQTL studies. bioRxiv (2025) doi:10.1101/2025.06.20.660695.

58. Malmström, E. et al. Human proteome distribution atlas for tissue-specific plasma proteome dynamics. Cell (2025) doi:10.1016/j.cell.2025.03.013.

59. Hou, K. et al. Accurate estimation of SNP-heritability from biobank-scale data irrespective of genetic architecture. Nat. Genet. 51, 1244–1251 (2019).

60. Wang, S., Zhang, J., Pan, T. & for Alzheimer’s Disease Neuroimaging Initiative. APOE ε4 is associated with higher levels of CSF SNAP-25 in prodromal Alzheimer’s disease. Neurosci. Lett. 685, 109–113 (2018).

61. Yang, F., Wang, J., GTEx Consortium, Pierce, B. L. & Chen, L. S. Identifying cis-mediators for trans-eQTLs across many human tissues using genomic mediation analysis. Genome Res. 27, 1859–1871 (2017).

62. Luo, J., Yang, H. & Song, B.-L. Mechanisms and regulation of cholesterol homeostasis. Nat. Rev. Mol. Cell Biol. 21, 225–245 (2020).

63. Tikka, A. & Jauhiainen, M. The role of ANGPTL3 in controlling lipoprotein metabolism. Endocrine 52, 187–193 (2016).

64. Tzotzas, T., Desrumaux, C. & Lagrost, L. Plasma phospholipid transfer protein (PLTP): review of an emerging cardiometabolic risk factor. Obes. Rev. 10, 403–411 (2009).

65. Mehta, A. & Shapiro, M. D. Apolipoproteins in vascular biology and atherosclerotic disease. Nat. Rev. Cardiol. 19, 168–179 (2022).

66. Wang, J.-Q. et al. Inhibition of ASGR1 decreases lipid levels by promoting cholesterol excretion. Nature 608, 413–420 (2022).

67. Szczepańska, E. & Gietka-Czernel, M. FGF21: A novel regulator of glucose and lipid metabolism and whole-body energy balance. Horm. Metab. Res. 54, 203–211 (2022).

68. Lp-PLA(2) Studies Collaboration et al. Lipoprotein-associated phospholipase A(2) and risk of coronary disease, stroke, and mortality: collaborative analysis of 32 prospective studies. Lancet 375, 1536–1544 (2010).

69. Gregson, J. M. et al. Genetic invalidation of Lp-PLA2 as a therapeutic target: Large-scale study of five functional Lp-PLA2-lowering alleles. Eur. J. Prev. Cardiol. 24, 492–504 (2017).

70. Aragam, K. G. et al. Discovery and systematic characterization of risk variants and genes for coronary artery disease in over a million participants. Nat. Genet. 54, 1803–1815 (2022).

71. Lambert, G., Sjouke, B., Choque, B., Kastelein, J. J. P. & Hovingh, G. K. The PCSK9 decade. J. Lipid Res. 53, 2515–2524 (2012).

72. Wu, S. A., Kersten, S. & Qi, L. Lipoprotein lipase and its regulators: An unfolding story. Trends Endocrinol. Metab. 32, 48–61 (2021).

73. Morton, R. E. & Mihna, D. Apolipoprotein F concentration, activity, and the properties of LDL controlling ApoF activation in hyperlipidemic plasma. J. Lipid Res. 63, 100166 (2022).

74. Abbate, A. et al. Interleukin-1 and the inflammasome as therapeutic targets in cardiovascular disease. Circ. Res. 126, 1260–1280 (2020).

75. Yoshida, T. & Hayashi, M. Role of Krüppel-like factor 4 and its binding proteins in vascular disease. J. Atheroscler. Thromb. 21, 402–413 (2014).

76. Accornero, F. & Molkentin, J. D. Placental growth factor as a protective paracrine effector in the heart. Trends Cardiovasc. Med. 21, 220–224 (2011).

77. Lalonde, S. et al. Integrative analysis of vascular endothelial cell genomic features identifies AIDA as a coronary artery disease candidate gene. Genome Biol. 20, 133 (2019).

78. Bruikman, C. S. et al. Genetic variants in SUSD2 are associated with the risk of ischemic heart disease. J. Clin. Lipidol. 14, 470–481 (2020).

79. Mohamed, S. A., Hanke, T., Schlueter, C., Bullerdiek, J. & Sievers, H.-H. Ubiquitin fusion degradation 1-like gene dysregulation in bicuspid aortic valve. J. Thorac. Cardiovasc. Surg. 130, 1531–1536 (2005).

80. Kartha, V. K. et al. Functional inference of gene regulation using single-cell multi-omics. Cell Genom 2, 100166 (2022).

81. Hou, L. et al. Multitissue H3K27ac profiling of GTEx samples links epigenomic variation to disease. Nat. Genet. 55, 1665–1676 (2023).

82. Siraj, L. et al. Functional dissection of complex trait variants at single-nucleotide resolution. Nature (2026) doi:10.1038/s41586-026-10121-6.

83. Benjamin J. Strober, Jordan Rossen, Xilin Jiang, Kangcheng Hou, Alkes L. Price. Estimating the proportion of disease heritability mediated by molecular traits. Abstract presented at the 77th annual meeting of the American Society of Human Genetics. 2025, Boston, MA.

84. Gerlach, P. A., Milind, N., Spence, J. P. & Pritchard, J. K. High false sign rates in transcriptome-wide association studies. bioRxiv (2025) doi:10.64898/2025.12.19.695550.

85. Warmerdam, C. A. R. et al. Trans-eQTLs reveal the architecture of human gene regulatory networks. medRxiv (2026) doi:10.64898/2026.02.04.26343575.

86. Mountjoy, E. et al. An open approach to systematically prioritize causal variants and genes at all published human GWAS trait-associated loci. Nat. Genet. 53, 1527–1533 (2021).

87. Ulland, T. K. & Colonna, M. TREM2-a key player in microglial biology and Alzheimer disease. Nat. Rev. Neurol. 14, 667–675 (2018).

88. Jonsson, T. et al. Variant of TREM2 associated with the risk of Alzheimer’s disease. N. Engl. J. Med. 368, 107–116 (2013).

89. Guerreiro, R. et al. TREM2 variants in Alzheimer’s disease. N. Engl. J. Med. 368, 117–127 (2013).

90. Deming, Y. et al. The MS4A gene cluster is a key modulator of soluble TREM2 and Alzheimer’s disease risk. Sci. Transl. Med. 11, eaau2291 (2019).

91. Kersten, S. ANGPTL3 as therapeutic target. Curr. Opin. Lipidol. 32, 335–341 (2021).

92. Musunuru, K. et al. Exome sequencing, ANGPTL3 mutations, and familial combined hypolipidemia. N. Engl. J. Med. 363, 2220–2227 (2010).

93. Dewey, F. E. et al. Genetic and pharmacologic inactivation of ANGPTL3 and cardiovascular disease. N. Engl. J. Med. 377, 211–221 (2017).

94. Yang, L. G., March, Z. M., Stephenson, R. A. & Narayan, P. S. Apolipoprotein E in lipid metabolism and neurodegenerative disease. Trends Endocrinol. Metab. 34, 430–445 (2023).

95. Saraiva, M. & O’Garra, A. The regulation of IL-10 production by immune cells. Nat. Rev. Immunol. 10, 170–181 (2010).

96. Sharifinejad, N. et al. The clinical, molecular, and therapeutic features of patients with IL10/IL10R deficiency: a systematic review. Clin. Exp. Immunol. 208, 281–291 (2022).

97. Tizaoui, K. et al. The role of PTPN22 in the pathogenesis of autoimmune diseases: A comprehensive review. Semin. Arthritis Rheum. 51, 513–522 (2021).

98. Mustelin, T., Bottini, N. & Stanford, S. M. The contribution of PTPN22 to rheumatic disease. Arthritis Rheumatol. 71, 486–495 (2019).

99. Glocker, E.-O. et al. Inflammatory bowel disease and mutations affecting the interleukin-10 receptor. N. Engl. J. Med. 361, 2033–2045 (2009).

100. Duffy, Á. et al. Development of a human genetics-guided priority score for 19,365 genes and 399 drug indications. Nat. Genet. 56, 51–59 (2024).

101. Schipper, M. et al. Prioritizing effector genes at trait-associated loci using multimodal evidence. Nat. Genet. 57, 323–333 (2025).

102. Mu, Z. et al. Impact of disease-associated chromatin accessibility QTLs across immune cell types and contexts. Cell Genom. 6, 101061 (2026).

103. Launch of world’s most significant protein study set to usher in new understanding for medicine. https://www.ukbiobank.ac.uk/news/launch-of-worlds-most-significant-protein-study-set-to-usher-in-new-understanding-for-medicine/ (2025).

104. Amariuta, T., Siewert-Rocks, K. & Price, A. L. Modeling tissue co-regulation estimates tissue-specific contributions to disease. Nat. Genet. 55, 1503–1511 (2023).

105. Hukku, A., Sampson, M. G., Luca, F., Pique-Regi, R. & Wen, X. Analyzing and reconciling colocalization and transcriptome-wide association studies from the perspective of inferential reproducibility. Am. J. Hum. Genet. 109, 825–837 (2022).

106. Liu, F. et al. Mitigating inconsistencies in GWAS follow-up analyses with LocusCompare2. Nat. Genet. 57, 2606–2613 (2025).

107. Koscielny, G. et al. Open Targets: a platform for therapeutic target identification and validation. Nucleic Acids Res. 45, D985–D994 (2017).

108. Minikel, E. V., Painter, J. L., Dong, C. C. & Nelson, M. R. Refining the impact of genetic evidence on clinical success. Nature 1–6 (2024).

109. Freund, M. K. et al. Phenotype-specific enrichment of Mendelian disorder genes near GWAS regions across 62 complex traits. Am. J. Hum. Genet. 103, 535–552 (2018).

110. Yang, J. et al. Conditional and joint multiple-SNP analysis of GWAS summary statistics identifies additional variants influencing complex traits. Nat. Genet. 44, 369–75, S1-3 (2012).

111. Chang, C. C. et al. Second-generation PLINK: rising to the challenge of larger and richer datasets. Gigascience 4, 7 (2015).

112. Bulik-Sullivan, B. et al. An atlas of genetic correlations across human diseases and traits. Nat. Genet. 47, 1236–1241 (2015).

113. The 1000 Genomes Project Consortium et al. A global reference for human genetic variation. Nature 526, 68–74 (2015).

114. Conneely, K. N. & Boehnke, M. So many correlated tests, so little time! Rapid adjustment of P values for multiple correlated tests. Am. J. Hum. Genet. 81, 1158–1168 (2007).

115. Loh, P.-R., Kichaev, G., Gazal, S., Schoech, A. P. & Price, A. L. Mixed-model association for biobank-scale datasets. Nat. Genet. 50, 906–908 (2018).

116. Tashman, K. C., Cui, R., O’Connor, L. J., Neale, B. M. & Finucane, H. K. Significance testing for small annotations in stratified LD-Score regression. medRxiv (2021) doi:10.1101/2021.03.13.21249938.

117. Baron, R. & Kneissel, M. WNT signaling in bone homeostasis and disease: from human mutations to treatments. Nat. Med. 19, 179–192 (2013).

118. Brunkow, M. E. et al. Bone dysplasia sclerosteosis results from loss of the SOST gene product, a novel cystine knot-containing protein. Am. J. Hum. Genet. 68, 577–589 (2001).

119. Nakashima, T. et al. Evidence for osteocyte regulation of bone homeostasis through RANKL expression. Nat. Med. 17, 1231–1234 (2011).

120. Movérare-Skrtic, S. et al. B4GALNT3 regulates glycosylation of sclerostin and bone mass. EBioMedicine 91, 104546 (2023).

121. Teslovich, T. M. et al. Biological, clinical and population relevance of 95 loci for blood lipids. Nature 466, 707–713 (2010).

122. Obeidat, M. et al. Surfactant protein D is a causal risk factor for COPD: results of Mendelian randomisation. Eur. Respir. J. 50, 1700657 (2017).

123. Jiang, Z. et al. A chronic obstructive pulmonary disease susceptibility gene, FAM13A, regulates protein stability of β-catenin. Am. J. Respir. Crit. Care Med. 194, 185–197 (2016).

124. Lin, X. et al. Tempo-spatial regulation of the Wnt pathway by FAM13A modulates the stemness of alveolar epithelial progenitors. EBioMedicine 69, 103463 (2021).

