## Supplementary figures for "Functionally informed cis and trans proteome-wide association studies prioritize disease-critical genes"

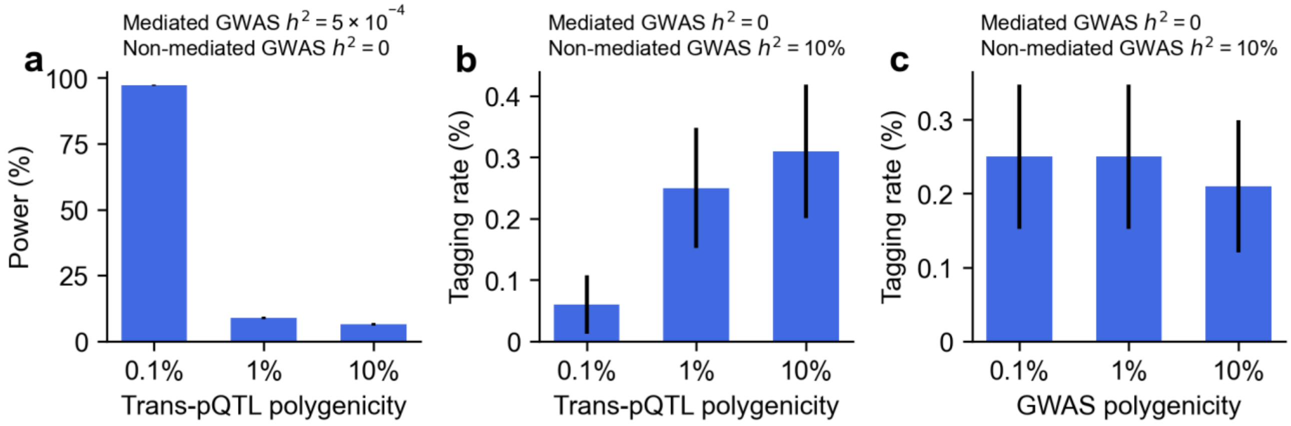


**Supplementary Figure 1. Secondary simulation analyses on sensitivity to pQTL and GWAS polygenicity. (a)** Power to detect protein-disease associations as a function of trans-pQTL polygenicity (fraction of causal SNPs: 0.1%, 1%, 10%), at fixed pQTL heritability = 10% and mediated heritability = $5 \times 10^{-4}$. **(b)** Tagging association rate at non-causal proteins as a function of trans-pQTL polygenicity, at fixed pQTL heritability = 10%, non-mediated heritability = 10%, and mediated heritability = 0. **(c)** Tagging association rate as a function of GWAS polygenicity (0.1%, 1%, 10% causal SNPs), at fixed pQTL heritability = 10%, and non-mediated heritability = 10%. Error bars denote 95% confidence intervals.

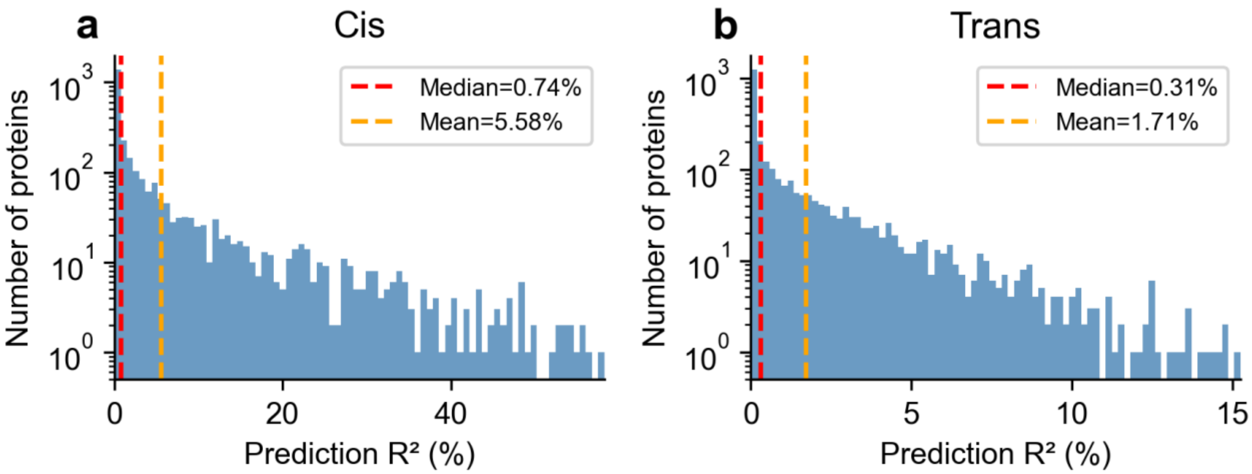


**Supplementary Figure 2. Distribution of cis and trans prediction accuracy in UKB-PPP**. Histograms of prediction $R^2$ (%) across proteins for **(a)** cis and **(b)** trans prediction models using well-imputed variants with functional priors (baseline-LD + pQTL annotations) at 20 PPCs. Y-axis uses log scale. Red and orange dashed lines indicate median and mean $R^2$, respectively.

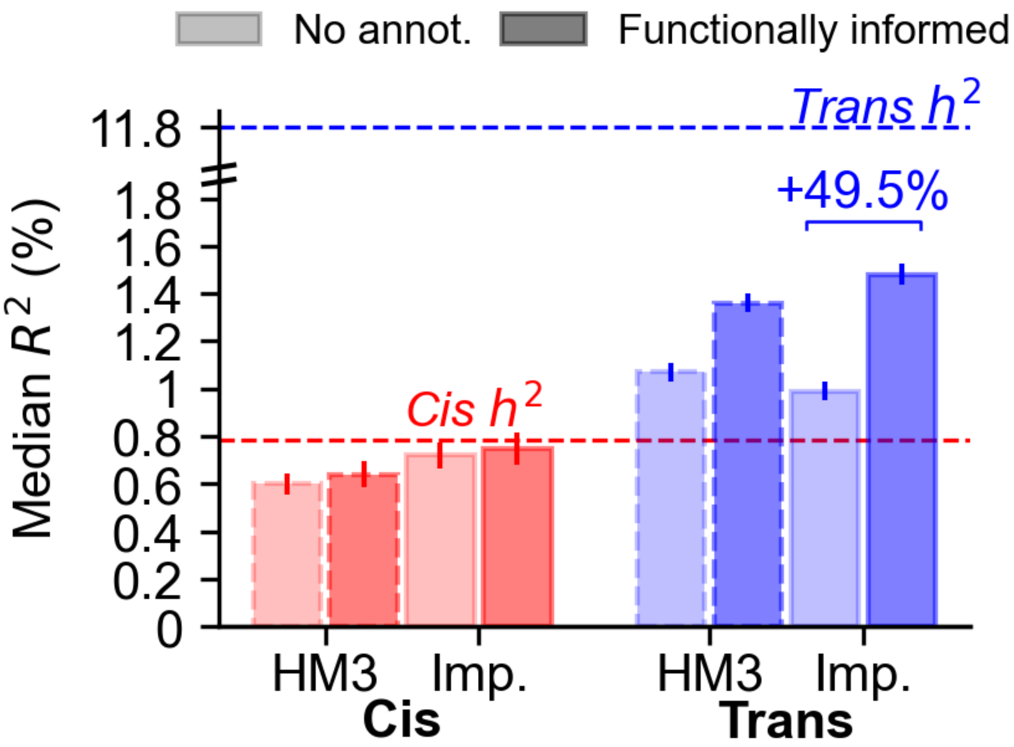


**Supplementary Figure 3.** Median prediction $R^2$ across proteins for cis and trans prediction models using HapMap3 (HM3) or well-imputed (Imp.) variants, with and without functional priors (baseline-LD model and proteome-wide pQTL annotations), without adjusting for PPCs. Dashed lines indicate median cis and trans heritability estimates. For trans models without functional annotations, imputed variants produce a lower median R2 but higher mean R2 compared to HM3 (median: 0.99% vs. 1.07%; mean: 2.62% vs. 2.52%), suggesting that the larger imputed SNP set improves prediction for well-powered proteins but introduces additional noise for the majority of proteins with low prediction accuracy.

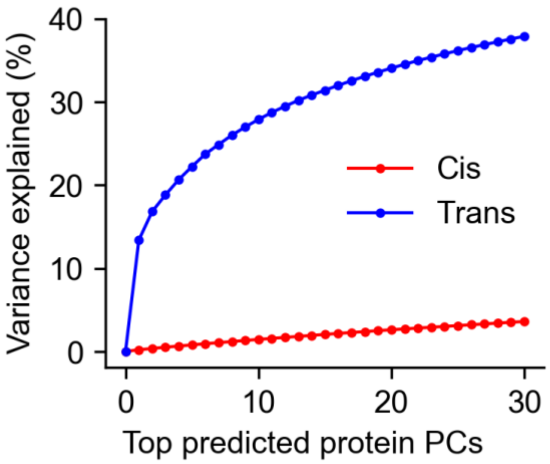


**Supplementary Figure 4. Variance explained by predicted protein principal components in UKB-PPP.** Proportion of variance in predicted protein levels explained by the top PPCs as a function of the number of PPCs, shown separately for cis (red) and trans-predicted (blue) protein levels. PPCs are computed separately for cis and trans predictions.

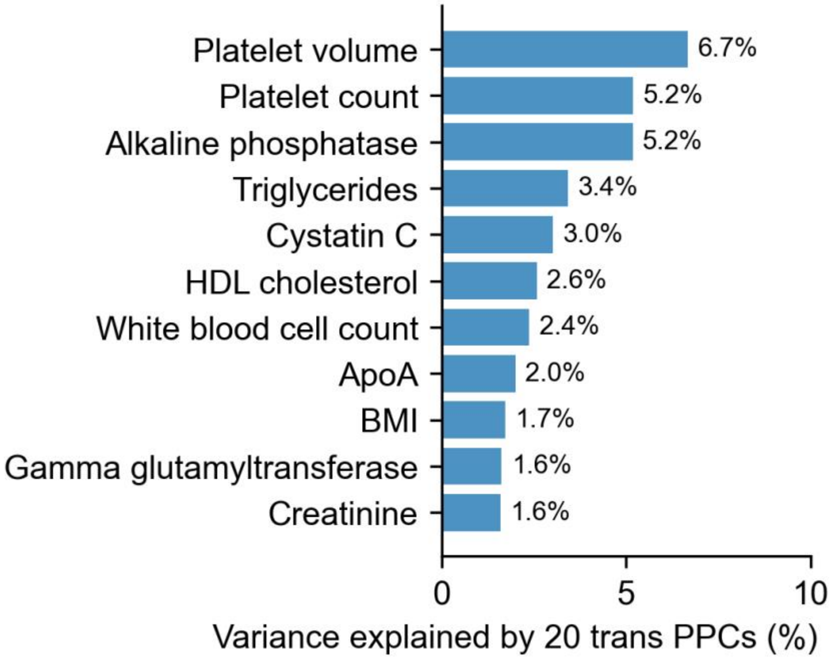


**Supplementary Figure 5. Biological relevance of trans-predicted protein principal components for UK Biobank traits.** Percentage of trait variance explained by the top 20 trans-PPCs across 32 UK Biobank diseases and traits; traits with the highest variance explained are shown.

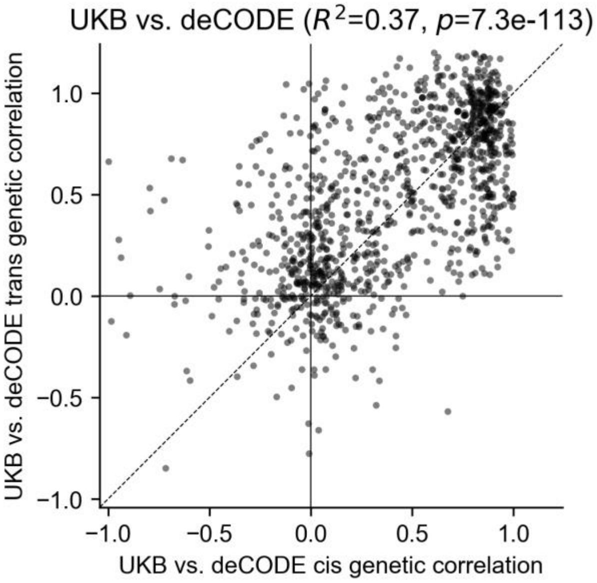


**Supplementary Figure 6. Concordance between UKB-PPP vs. deCODE cis and trans cross-cohort genetic correlations.** Scatter plot of cis (x-axis) versus trans (y-axis) cross-cohort genetic correlations for proteins present in both UKB-PPP and deCODE. Each point denotes one protein. Pearson correlation and P-value are shown.

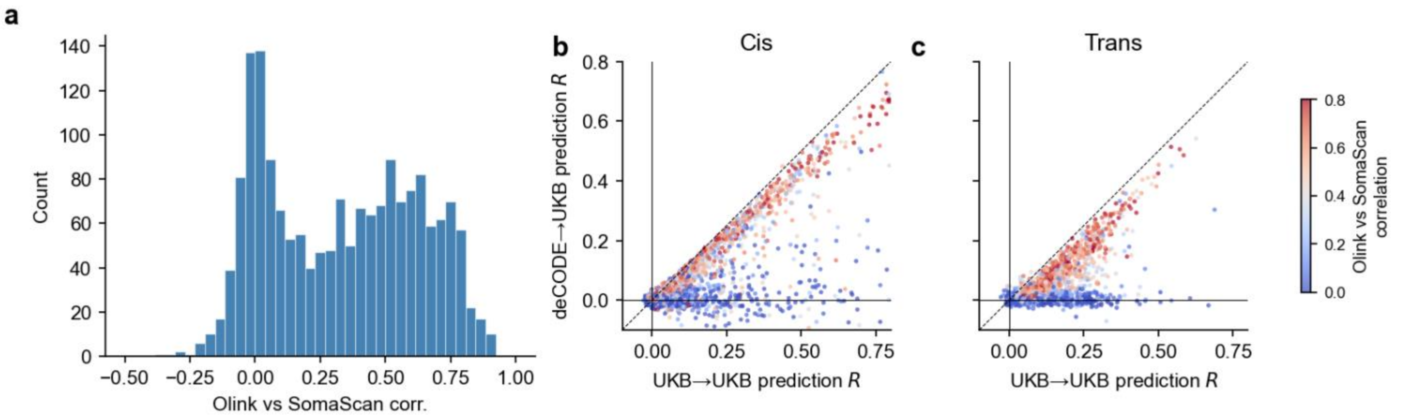


**Supplementary Figure 7. Prediction accuracy is consistent with individual-level protein level correlation.** **(a)** Distribution of individual-level protein level correlations between Olink vs. SomaScan platforms, as reported in Eldjarn et al.[55]. **(b-c)** Comparisons of cross-cohort prediction R in UKB-PPP and deCODE for **(b)** cis and **(c)** trans predictions across proteins assayed in both platforms. Each point denotes one protein, colored by individual-level Olink-SomaScan protein level correlation.

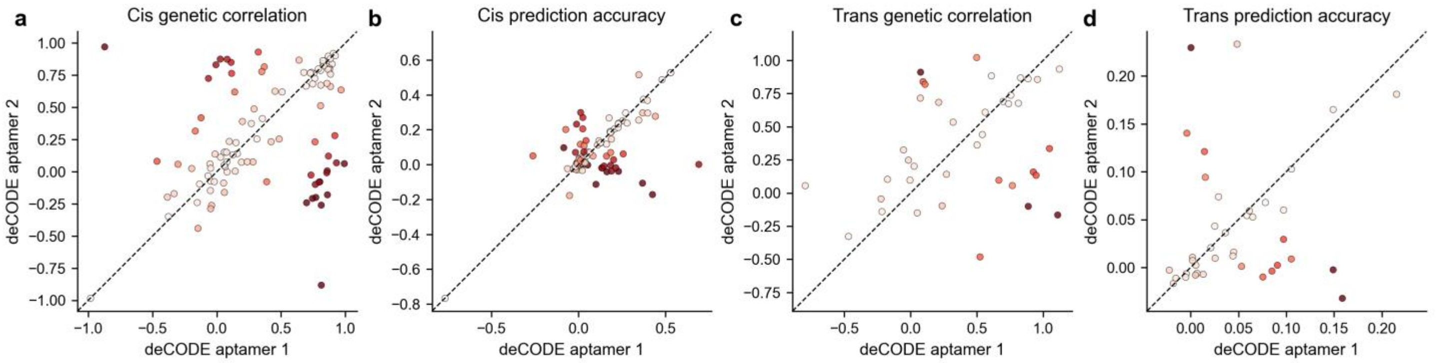


**Supplementary Figure 8. Cross-cohort portability for deCODE aptamer pairs targeting the same proteins.** **(a)** Cis genetic correlation between UKB-PPP and deCODE for aptamer 1 and 2 for deCODE aptamers targeting the same protein. **(b)** Cis prediction accuracy for the same aptamer pairs. **(c)** Trans genetic correlation for the same aptamer pairs. **(d)** Trans prediction accuracy for the same aptamer pairs. Color intensity reflects statistical significance of the difference between aptamers.

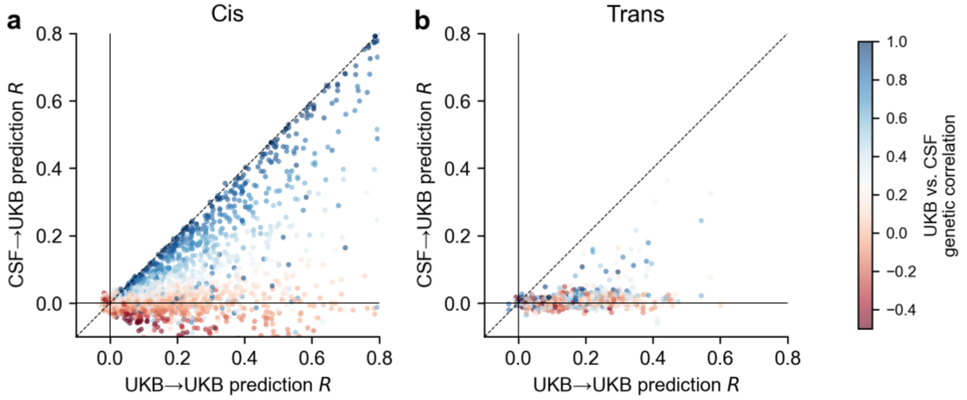


**Supplementary Figure 9. UKB-PPP versus CSF cross-cohort comparison for prediction accuracy.** Scatter plots of cis **(a)** and trans **(b)** prediction R in UKB-PPP versus CSF for matched proteins assayed in both UKB-PPP and CSF. Each point denotes one protein; color denotes cross-cohort genetic correlation. Dashed line indicates y=x.

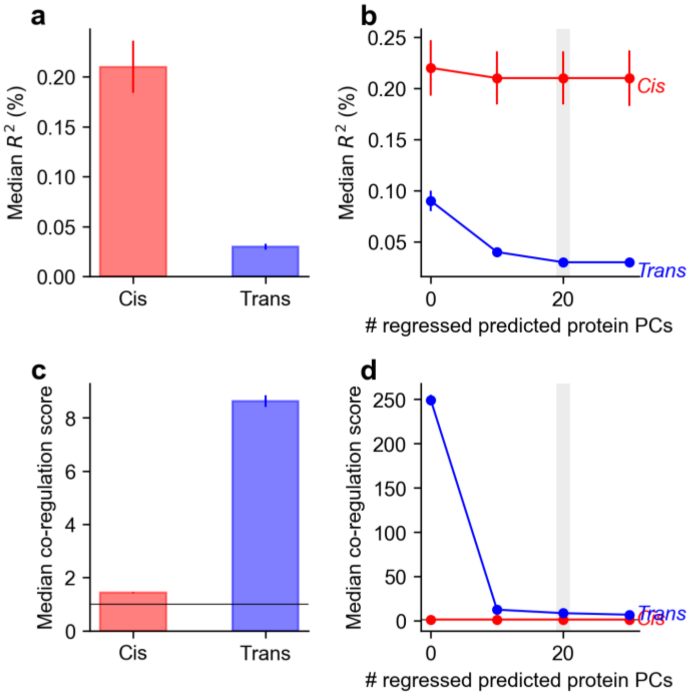


**Supplementary Figure 10. deCODE protein prediction model performance. (a)** Median prediction $R^2$ for cis and trans models. **(b)** Median prediction $R^2$ as a function of the number of adjusted PPCs. **(c)** Median co-regulation score for cis and trans predictions with 20 PPCs adjusted. **(d)** Median co-regulation score as a function of the number of adjusted PPCs.

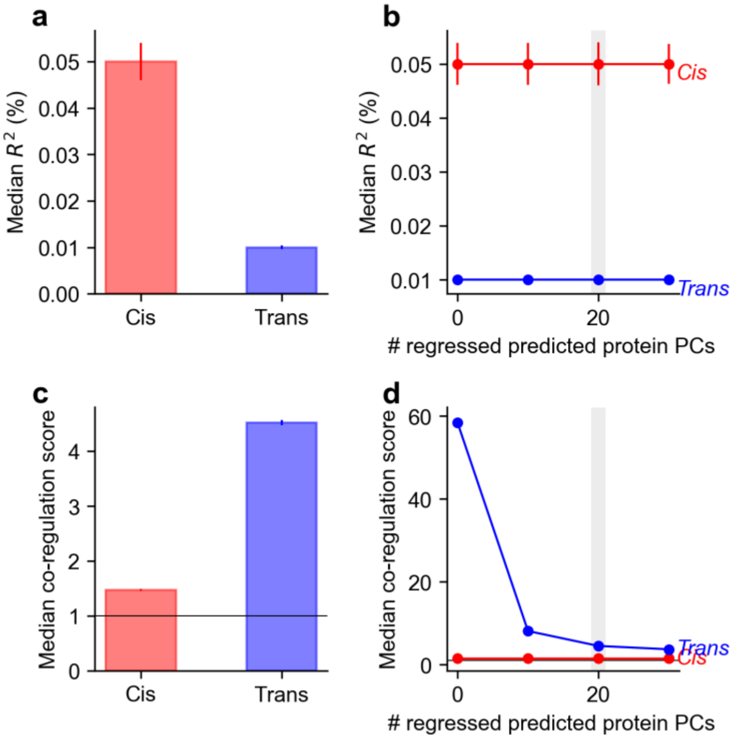


**Supplementary Figure 11. CSF protein prediction model performance. (a)** Median prediction $R^2$ for cis and trans models. **(b)** Median prediction $R^2$ as a function of the number of adjusted PPCs. **(c)** Median co-regulation score for cis and trans predictions with 20 PPCs adjusted. **(d)** Median co-regulation score as a function of the number of adjusted PPCs.

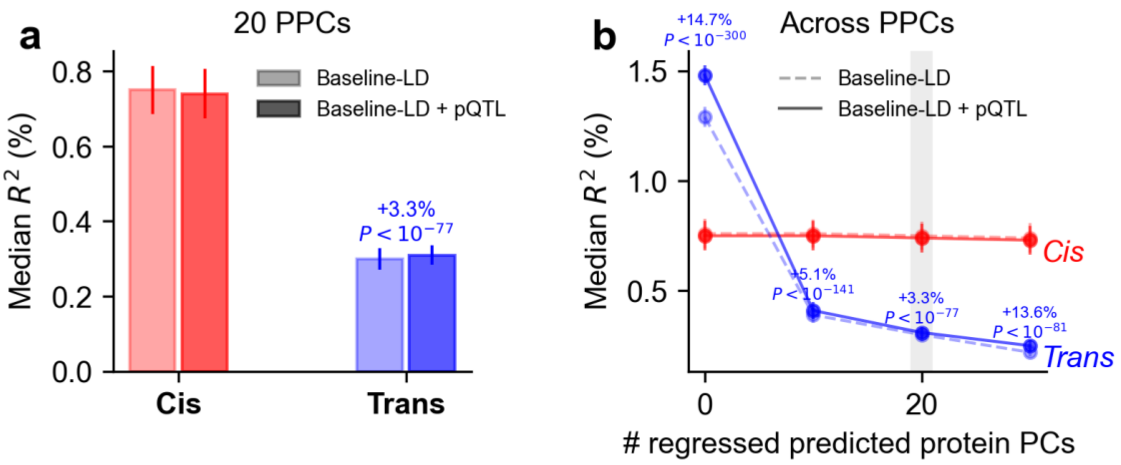


**Supplementary Figure 12. Impact of pQTL priors on protein prediction accuracy in UKB-PPP.** Comparison of baseline-LD versus baseline-LD plus proteome-wide pleiotropy (baseline-LD+pQTL) models for cis and trans predictors using imputed variants. **(a)** Median prediction $R^2$ at 20 adjusted PPCs; percentage improvement and one-sided paired Wilcoxon P-value shown for trans prediction. **(b)** Median prediction $R^2$ as a function of the number of adjusted PPCs; dashed lines denote baseline-LD and solid lines denote baseline-LD+pQTL.

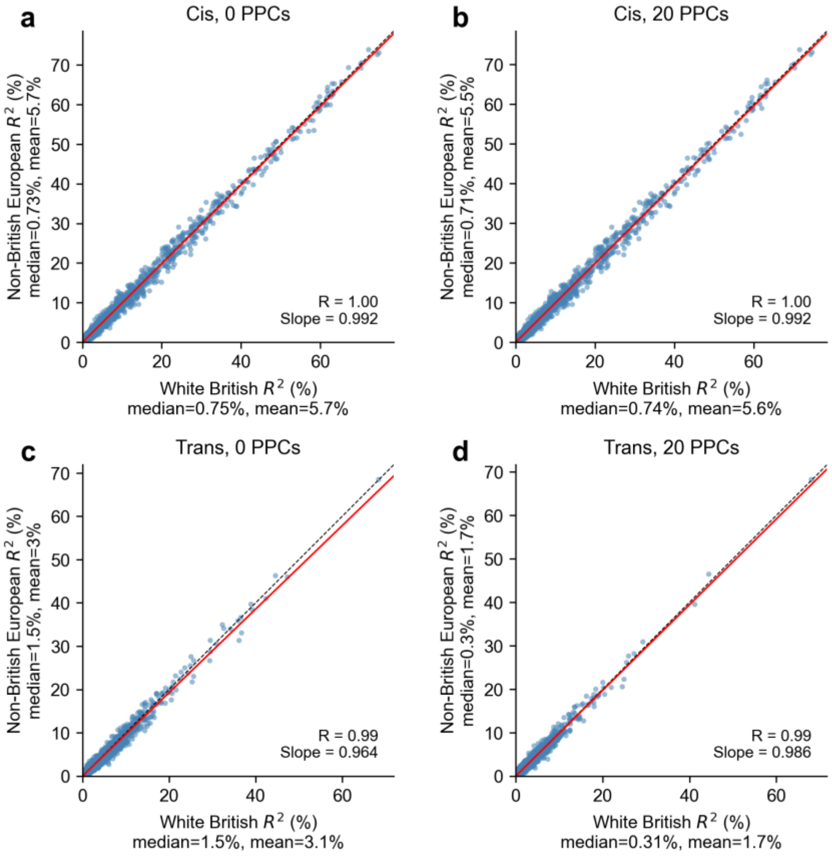


**Supplementary Figure 13. Prediction accuracy in non-British European UKB-PPP participants.** Per-protein prediction $R^2$ in 9,746 White British (x-axis) versus 3,044 non-British European (y-axis) held-out UKB-PPP individuals for cis **(a,b)** and trans **(c,d)** prediction models using imputed variants with functional priors (baseline-LD + pQTL annotations), at 0 PPCs **(a,c)** and 20 PPCs **(b,d)**. Each point denotes one protein. Dashed line indicates y=x; red line shows OLS regression slope.

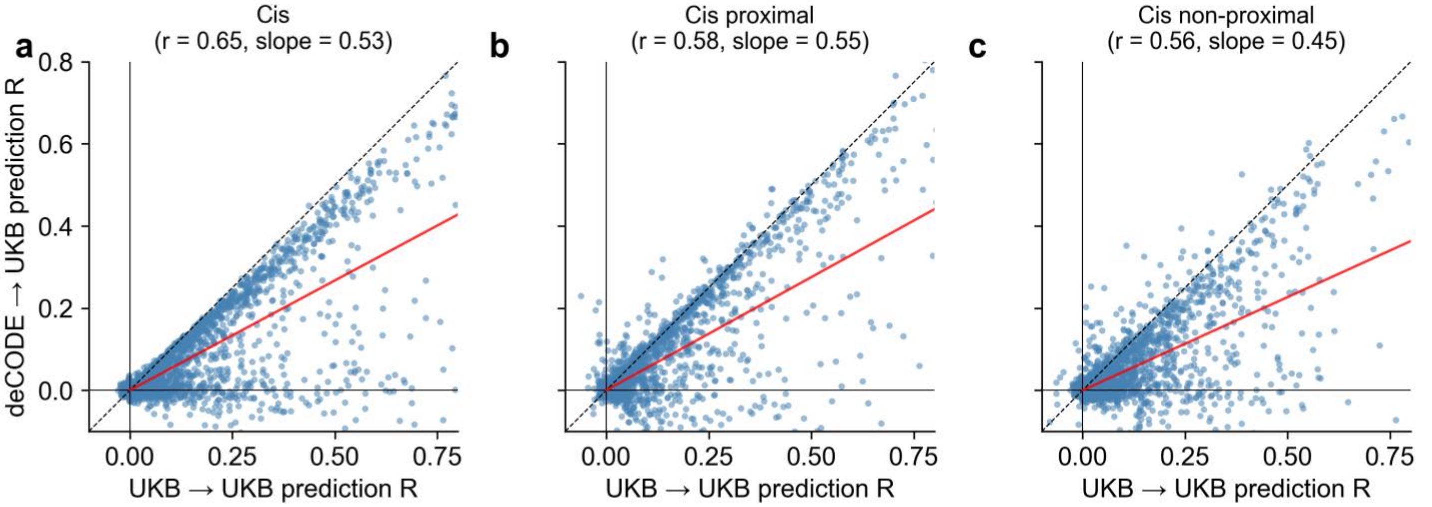


**Supplementary Figure 14. Cross-cohort cis prediction portability across variant subsets.** Cross-cohort prediction performance (UKB-PPP vs. deCODE) for cis-predicted protein levels using **(a)** all cis variants, **(b)** cis proximal variants (within 2 kb of transcribed region), and **(c)** cis non-proximal variants (remaining ±1 Mb). Each point denotes a protein. Dashed line indicates y=x; red line shows OLS regression slope.

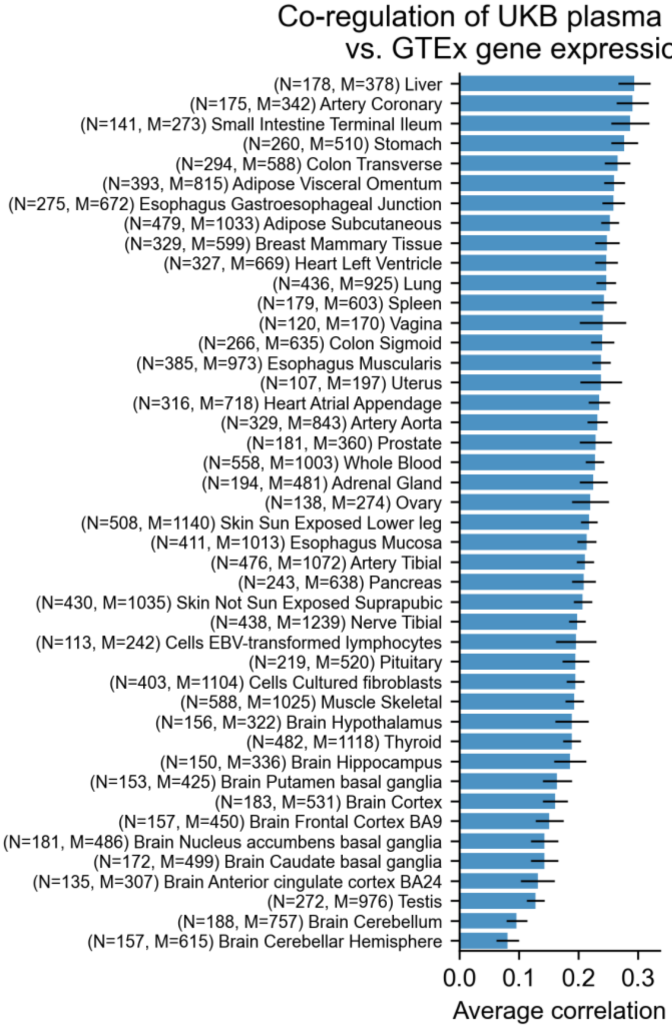


**Supplementary Figure 15. Concordance between cis-pQTL and cis-eQTL predictions across GTEx tissues.** We compared UKB-PPP cis-predicted protein levels with cis-predicted gene expression across 49 GTEx tissues[12], computing the average correlation between cis-predicted protein levels and cis-predicted expression levels across matched protein-gene pairs for each tissue. Average correlation between UKB-PPP cis-predicted protein levels and GTEx cis-predicted gene expression across 49 tissues, ordered by concordance. N denotes GTEx sample size; M denotes matched protein-gene pairs; error bars denote standard errors.

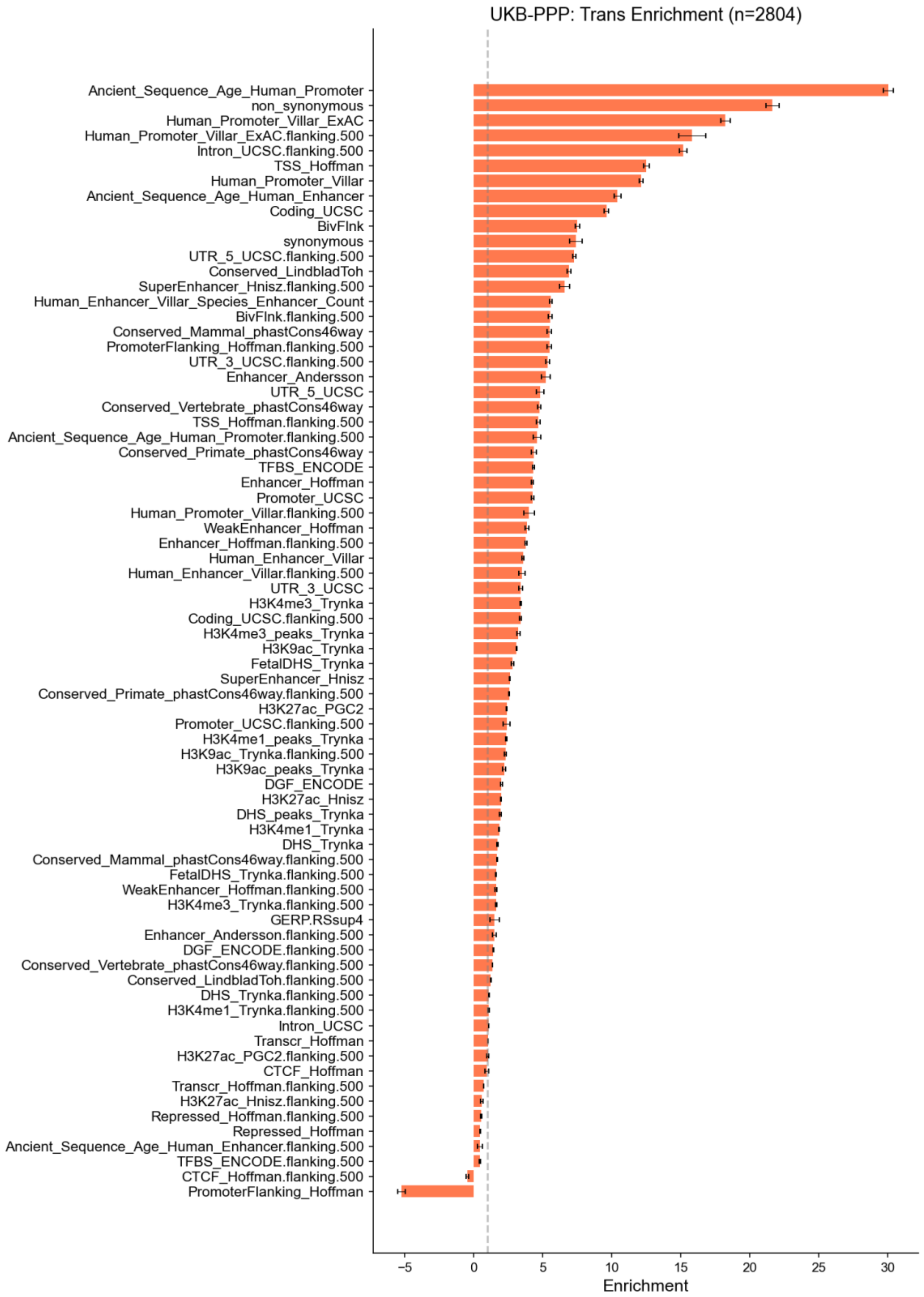


**Supplementary Figure 16. Functional enrichment of trans-pQTL heritability.** We applied S-LDSC[46] to partition trans-pQTL SNP-heritability across 96 baseline-LD v2.2 functional annotation categories, excluding continuous annotations and MAF bin. Horizontal bars show enrichment; error bars denote jackknife standard errors across proteins grouped by genomic position. Dashed vertical line indicates enrichment = 1 (no enrichment). Enrichment was estimated by applying S-LDSC to each protein separately, then meta-analyzing per-annotation coefficients across proteins and converting to enrichment via the annotation overlap matrix[25]. The biologically implausible significantly negative estimate for “PromoterFlanking_Hoffman” likely reflects mis-calibrated standard errors[116] for small annotations with substantial collinearity with other annotations.

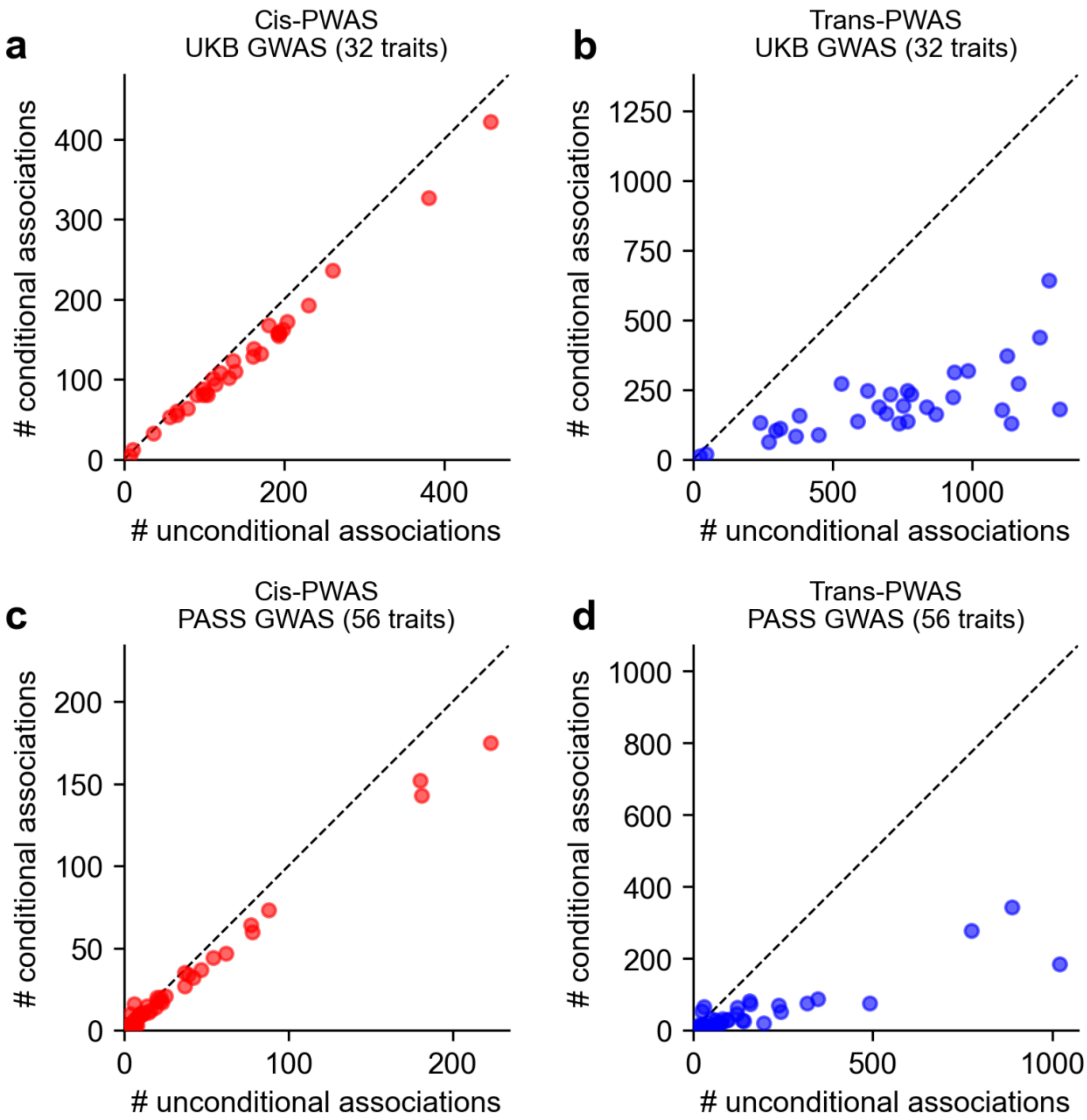


**Supplementary Figure 17. Stepwise conditional analysis of PWAS associations.** **(a)** Cis-PWAS and **(b)** trans-PWAS for 32 UK Biobank diseases/traits, and **(c)** cis-PWAS and **(d)** trans-PWAS for 56 additional disease/traits with publicly available summary statistics. Each panel plots unconditional versus conditional significant associations per trait ($P < 10^{-5}$); points denote traits, dashed lines indicate y=x, and text reports mean reduction after conditioning.

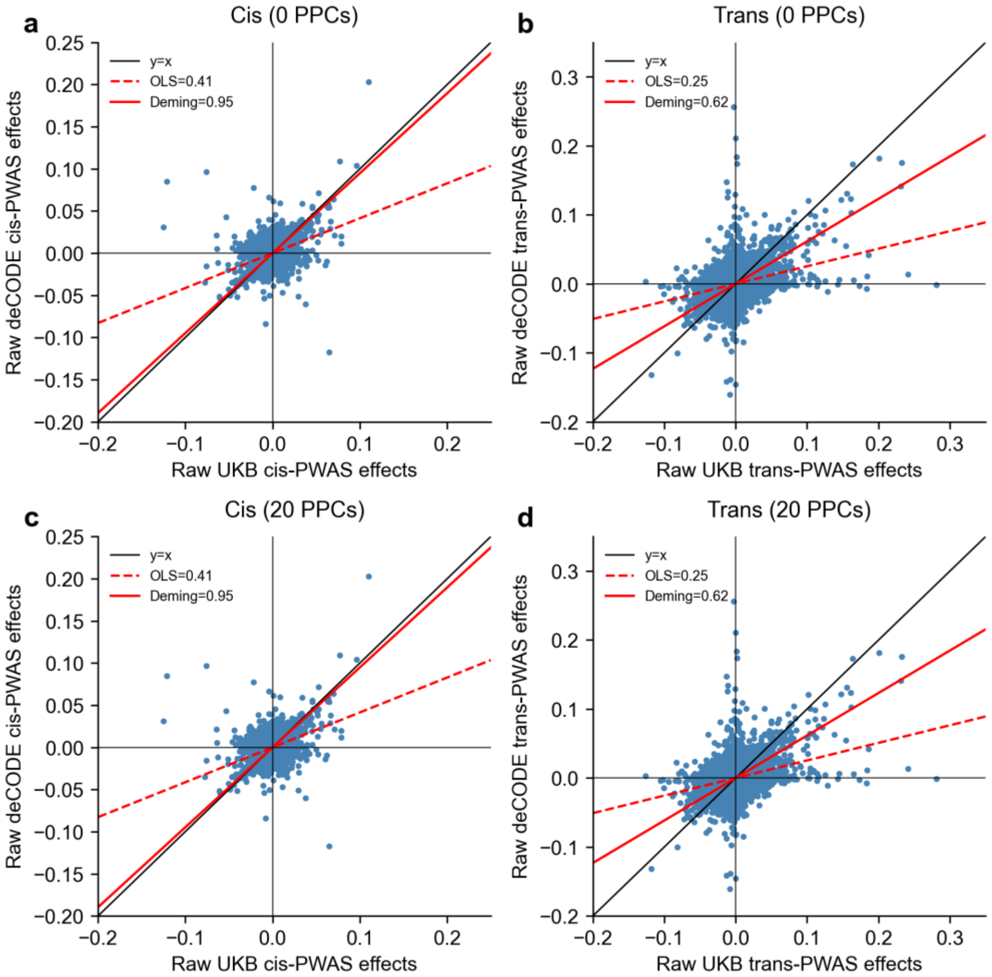


**Supplementary Figure 18. Cross-cohort PWAS concordance between UKB-PPP and deCODE using unadjusted PWAS Z-scores.** Cis-PWAS **(a,c)** and trans-PWAS **(b,d)** are shown at 0 PPCs **(a,b)** and 20 PPCs **(c,d)**. Each point denotes a protein-trait pair. The solid black line denotes y=x, the solid red line denotes the Deming regression slope, and the dashed red line denotes the OLS slope.

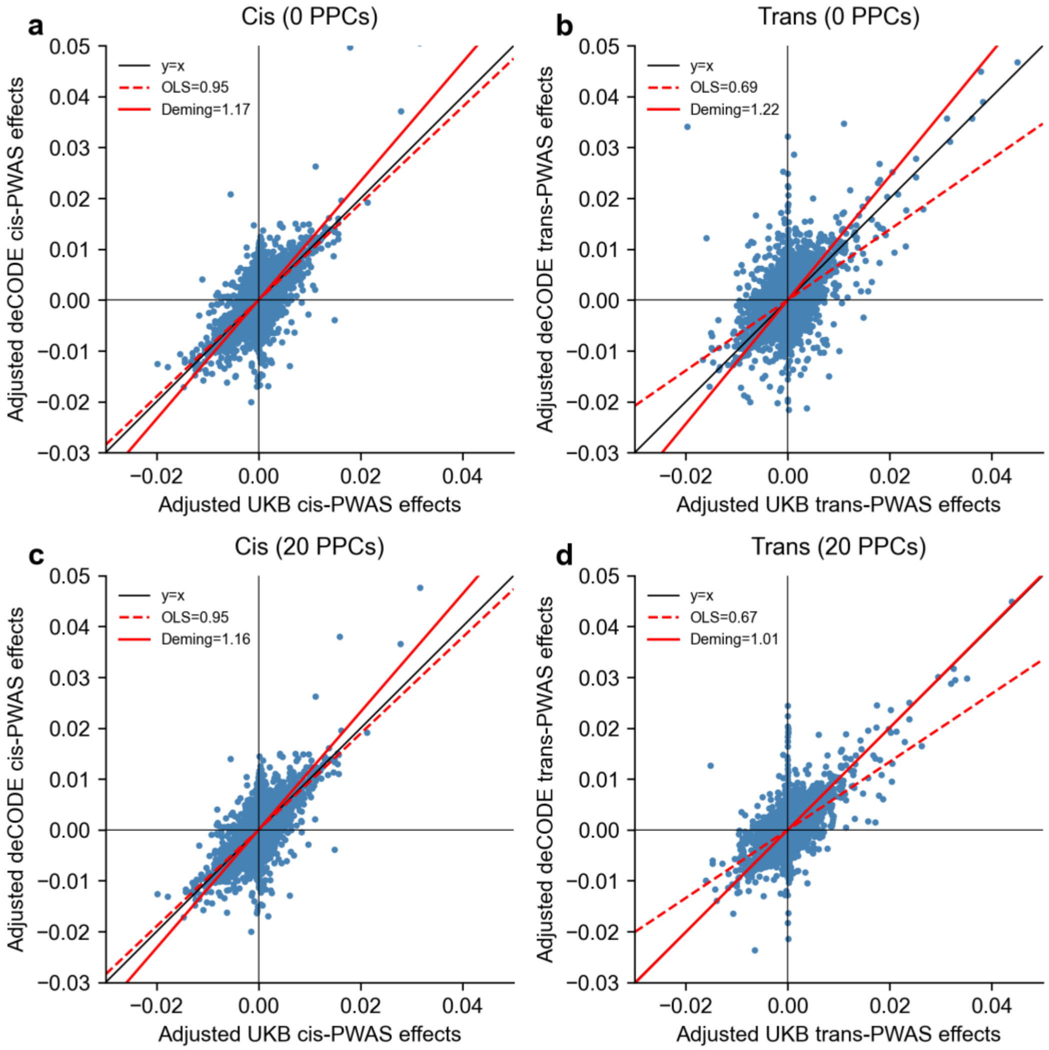


**Supplementary Figure 19. Cross-cohort PWAS concordance between UKB-PPP and deCODE using different numbers of PPCs.** Cis-PWAS **(a,c)** and trans-PWAS **(b,d)** are shown at 0 PPCs (a,b) and 20 PPCs (c,d). Each point denotes a protein-trait pair. The solid black line denotes y=x, the solid red line denotes the Deming regression slope, and the dashed red line denotes the OLS slope.

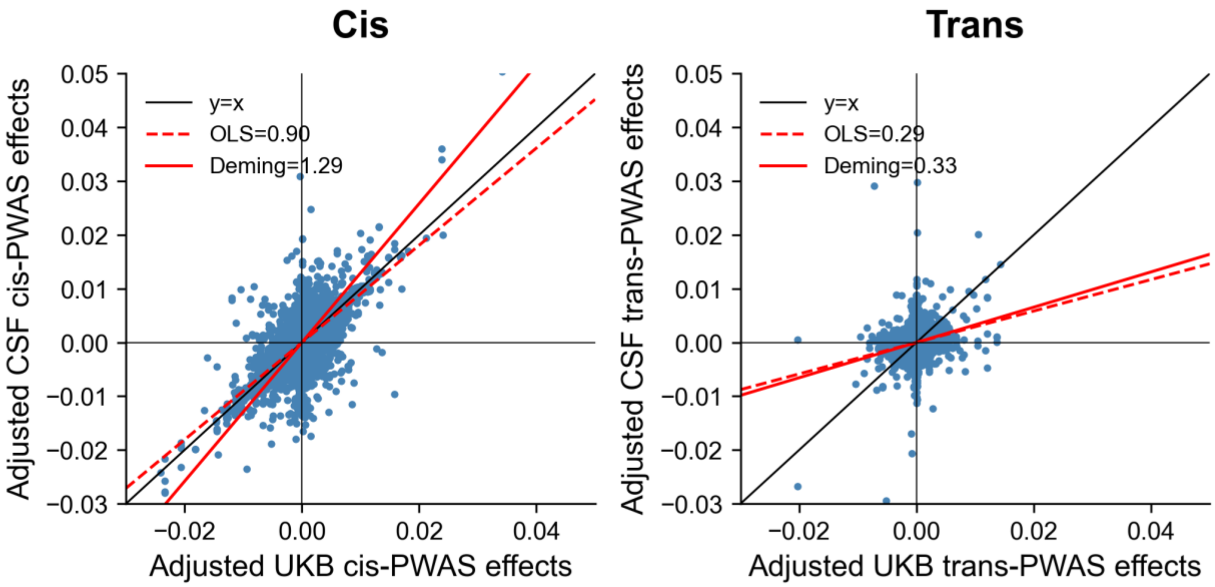

**Supplementary Figure 20. Cross-cohort PWAS concordance between UKB-PPP and CSF.** Cis-PWAS **(a)** and trans-PWAS **(b)** are shown for proteins shared across cohorts. Each point denotes a protein-trait pair. The solid black line denotes y=x, the solid red line denotes the Deming regression slope, and the dashed red line denotes the OLS slope. Lower concordance is consistent with smaller CSF sample size and tissue-specific proteomic regulation. These results use adjusted PWAS effects with 20 PPCs.

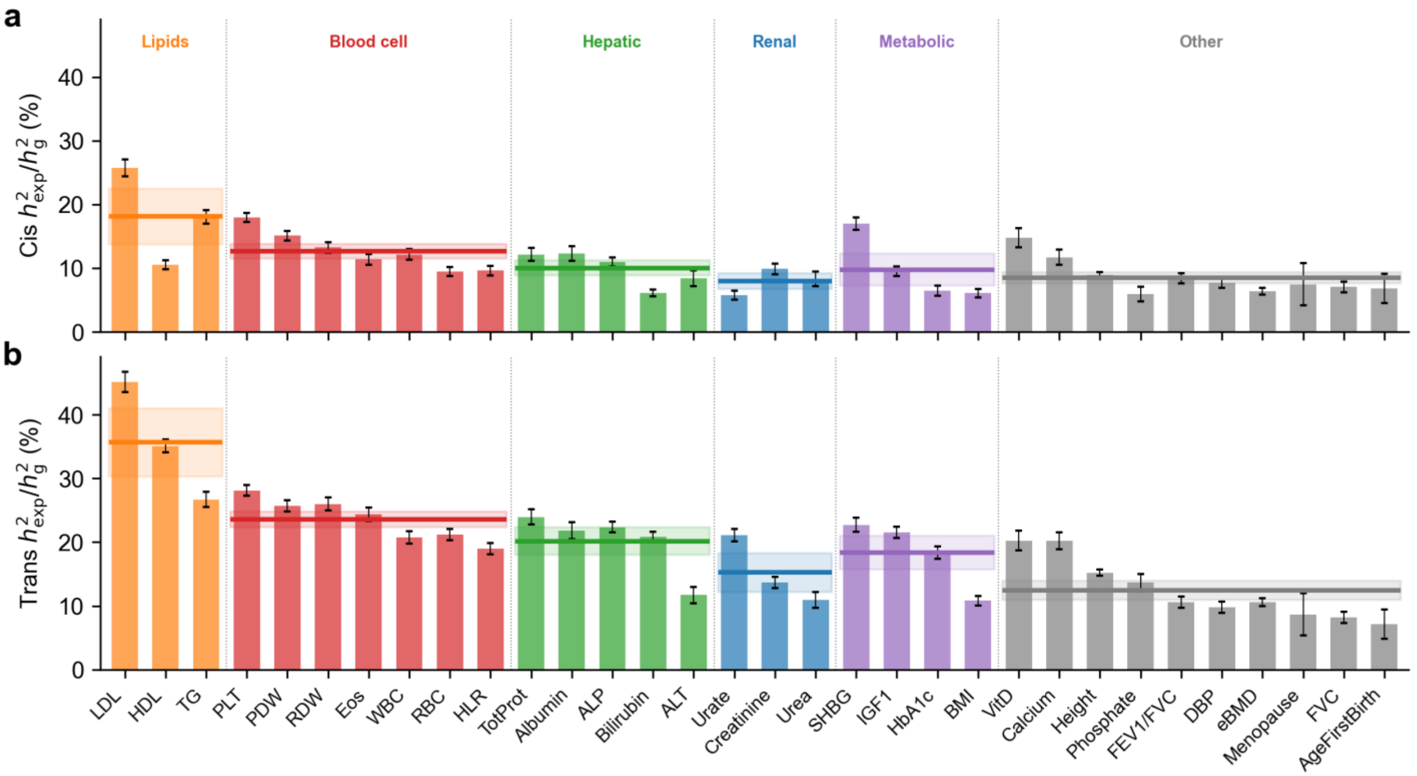


**Supplementary Figure 21. Variance explained by predicted protein levels across trait categories.** Cis **(a)** and trans **(b)** proportion of variance explained across 32 UK Biobank traits, grouped by trait category (Lipids, Blood cell, Hepatic, Renal, Metabolic, Other). Traits ordered within each category by average variance explained. Horizontal lines and shaded bands denote category mean and standard errors of the mean.

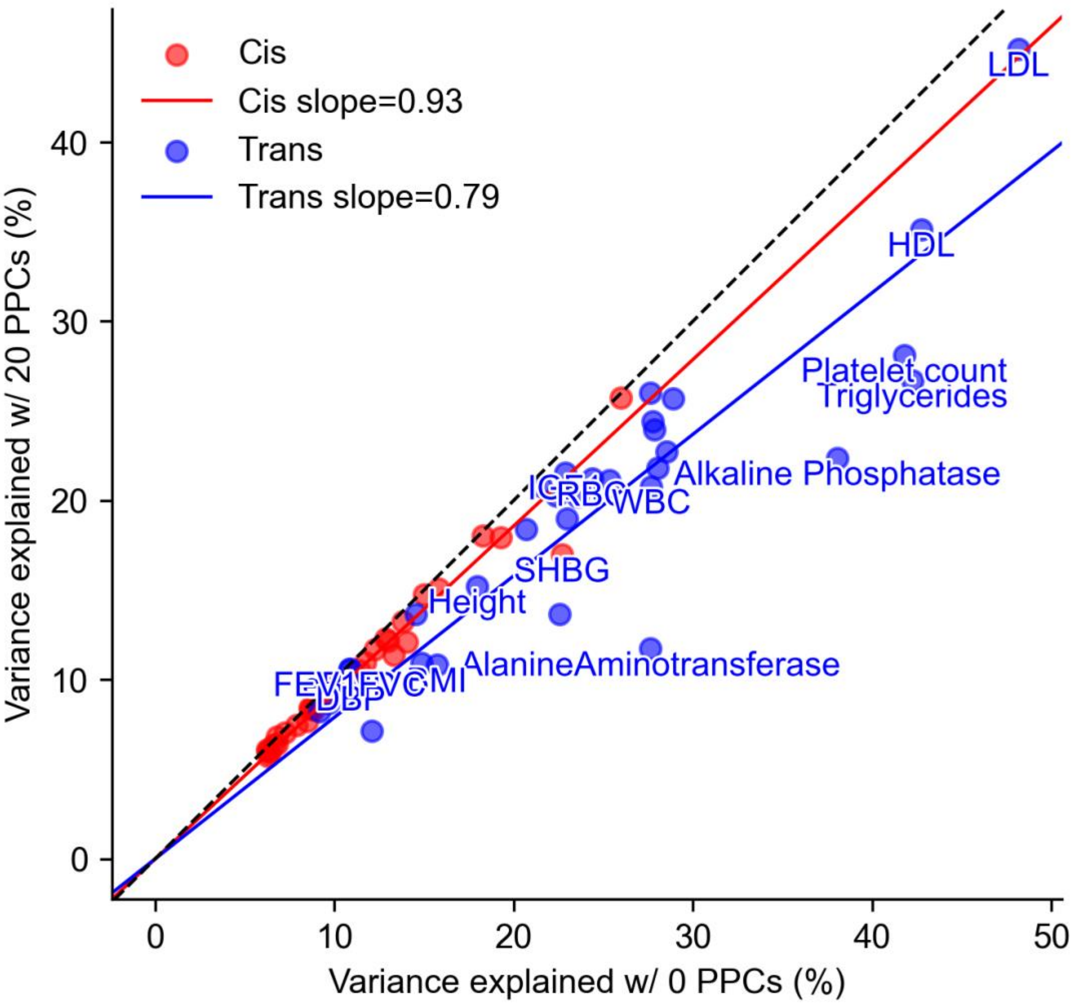


**Supplementary Figure 22. Impact of PPC adjustment on trait variance explained**. Comparison of the proportion of SNP-heritability explained at 0 PPCs (x-axis) versus 20 PPCs (y-axis) across traits, shown separately for cis-predicted (red) and trans-predicted (blue) protein levels. Each point denotes one trait. Solid red and blue lines indicate subset-specific slope fits, and the black dashed line denotes y=x.

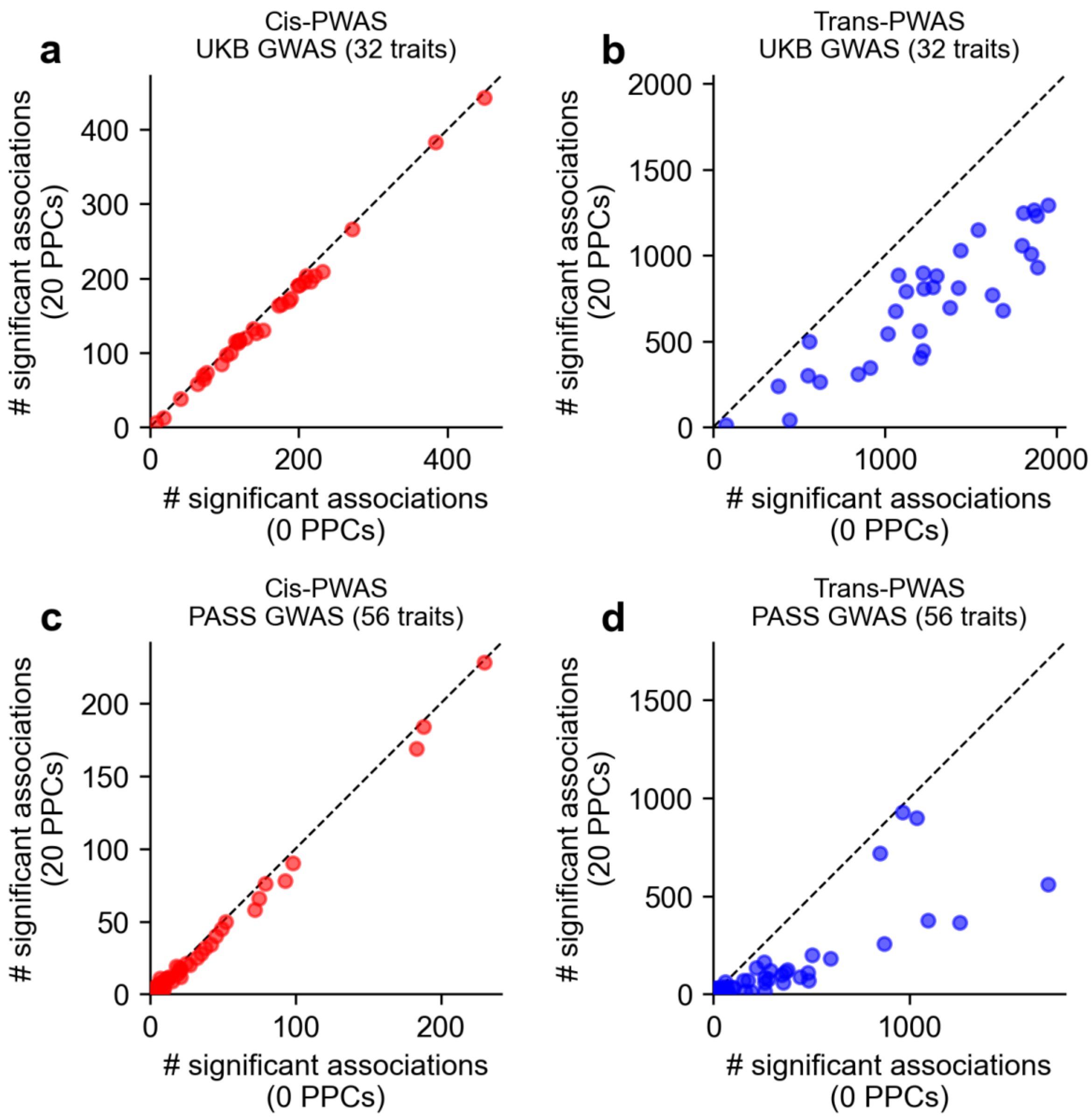


**Supplementary Figure 23. Impact of PPC adjustment on number of significant PWAS associations.** Cis-PWAS **(a,c)** and trans-PWAS **(b,d)** are shown for UKB GWAS (32 traits; **a,b**) and publicly available GWAS (56 traits; **c,d**). Each point denotes one trait, comparing significant associations at 0 PPCs (x-axis) versus 20 PPCs (y-axis), using a two-sided significance threshold of $P < 10^{-5}$. Dashed lines denote y=x.

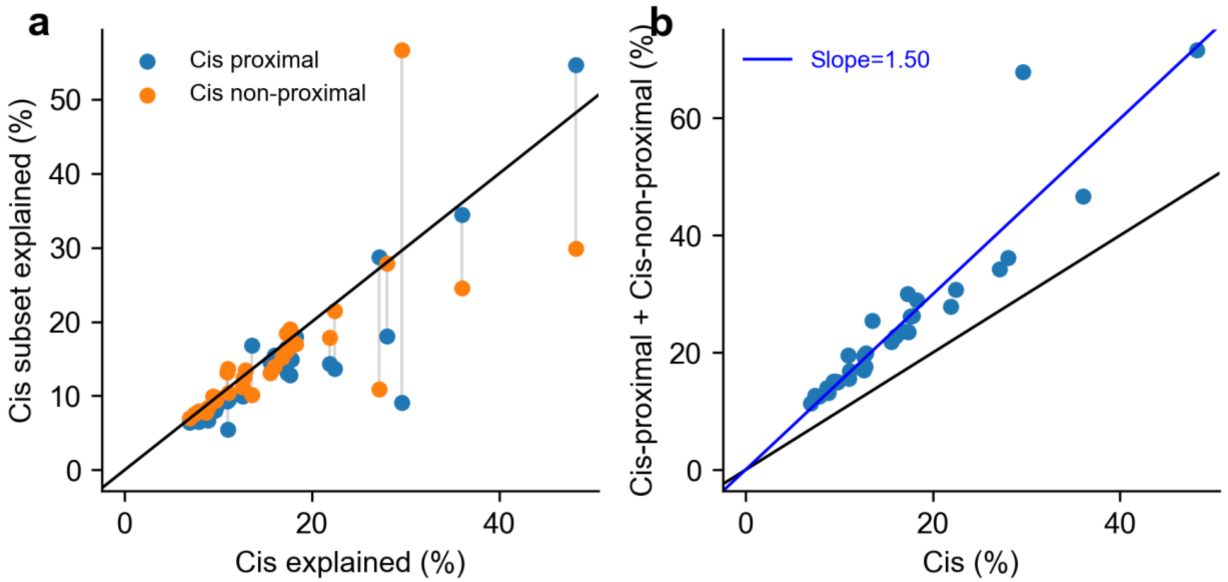


**Supplementary Figure 24. Trait variance explained by proximal and non-proximal cis variant components.** **(a)** Cis proximal (within 2 kb of transcribed region) vs cis non-proximal (remaining ±1 Mb) variance explained across 32 UK Biobank traits. **(b)** Sum of cis proximal and cis non-proximal variance explained versus total cis variance explained. Each point denotes one trait.

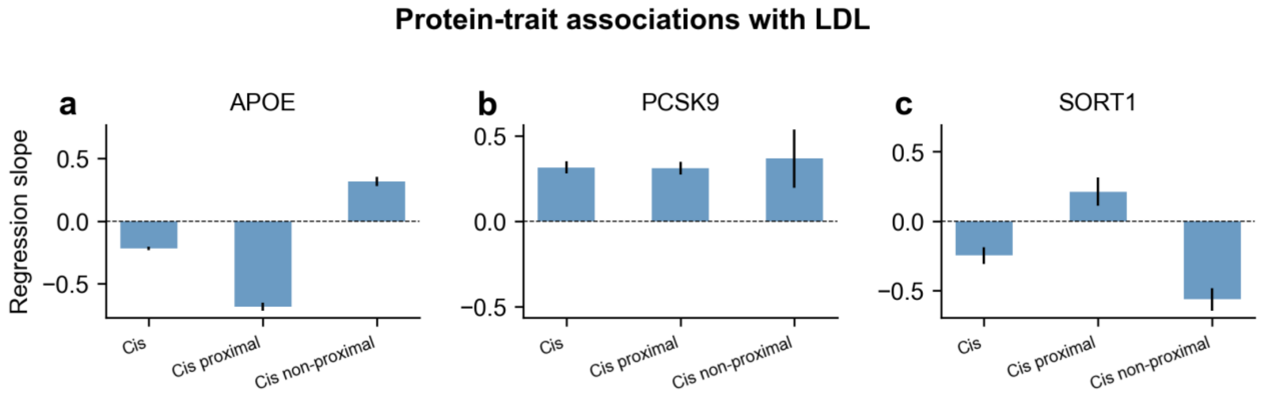


**Supplementary Figure 25. Examples of distinct or similar disease effects across cis variant subsets.** Regression coefficients (95% CI) for LDL cholesterol association with predicted protein levels using all cis variants, cis proximal variants, and cis non-proximal variants are shown for **(a)** *APOE*, **(b)** *PCSK9*, and **(c)** *SORT1*.

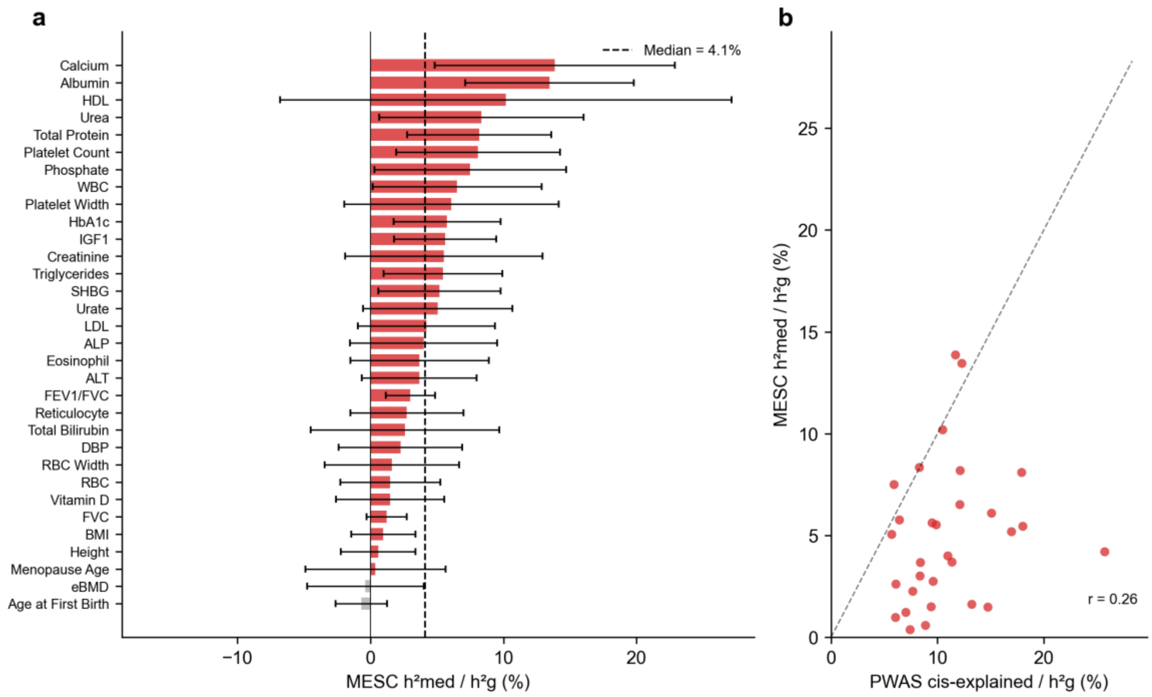


**Supplementary Figure 26. Mediated heritability estimates from MESC using cis-pQTL data.** We applied MESC[13] to estimate the proportion of trait heritability mediated by cis-pQTL effects to the 32 approximately independent UK Biobank diseases/traits using cis-pQTL weights from UKB-PPP and LD scores computed from UK Biobank European-ancestry individuals. We avoided sample overlap between pQTL and GWAS by excluding UKB-PPP participants from GWAS computation. **(a)** Proportion of SNP-heritability mediated through cis-pQTL effects across 32 UK Biobank traits, with 95% confidence intervals. **(b)** Comparison of MESC-mediated heritability versus cis-predicted protein-level variance explained from PolyPWAS across traits.

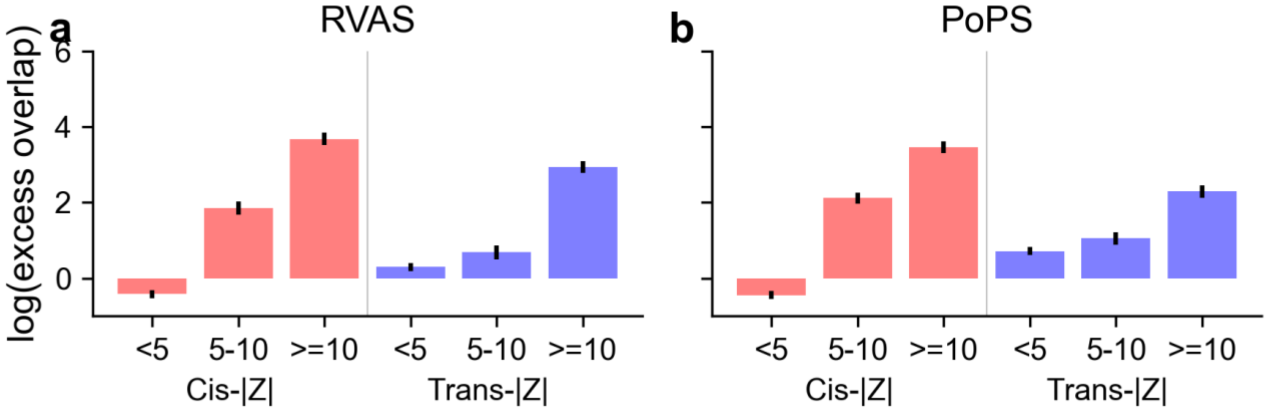


**Supplementary Figure 27. Marginal enrichments of cis PWAS and trans PWAS-prioritized genes in RVAS and PoPS validation sets.** Log(excess overlap) with **(a)** RVAS and **(b)** PoPS validation gene sets at 20 PPCs using UKB-PPP pQTL models. Within each panel, red and blue bars show marginal cis-/trans-|Z| strata (<5, 5-10, >=10). Error bars denote standard errors.

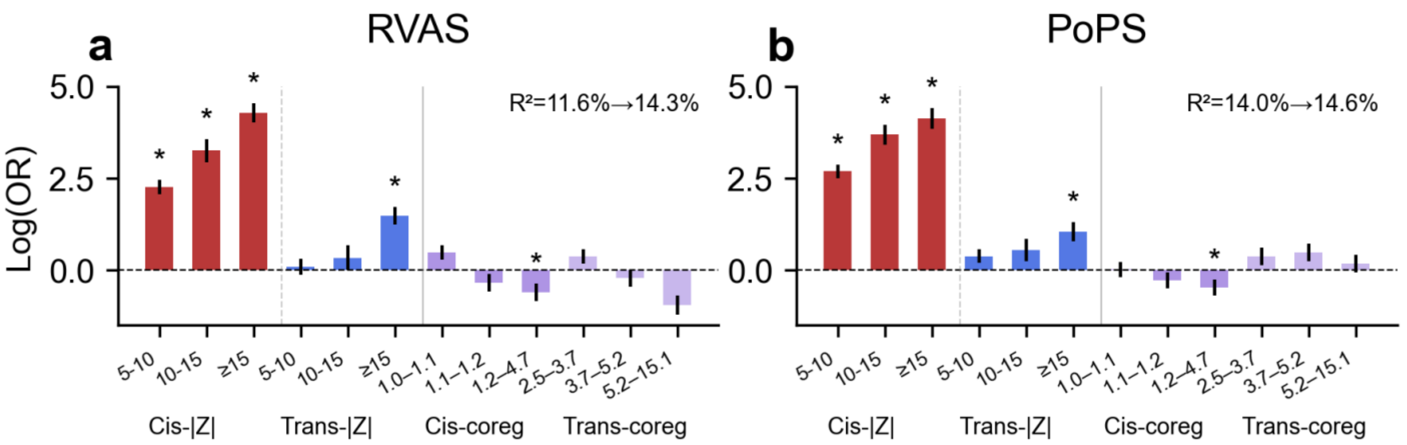


**Supplementary Figure 28. Logistic regression coefficients with co-regulation features.** Coefficients (log odds ratio) for UKB-PPP pQTL models, predicting **(a)** RVAS and **(b)** PoPS at 20 PPCs. Bars show binned cis-|Z|, trans-|Z|, cis co-regulation quartile, and trans co-regulation quartile features. We show pseudo-$R^2$ with or without co-regulation features. Asterisks denote terms significant in both RVAS and PoPS within the same cohort (|z| > 1.96 in both panels).

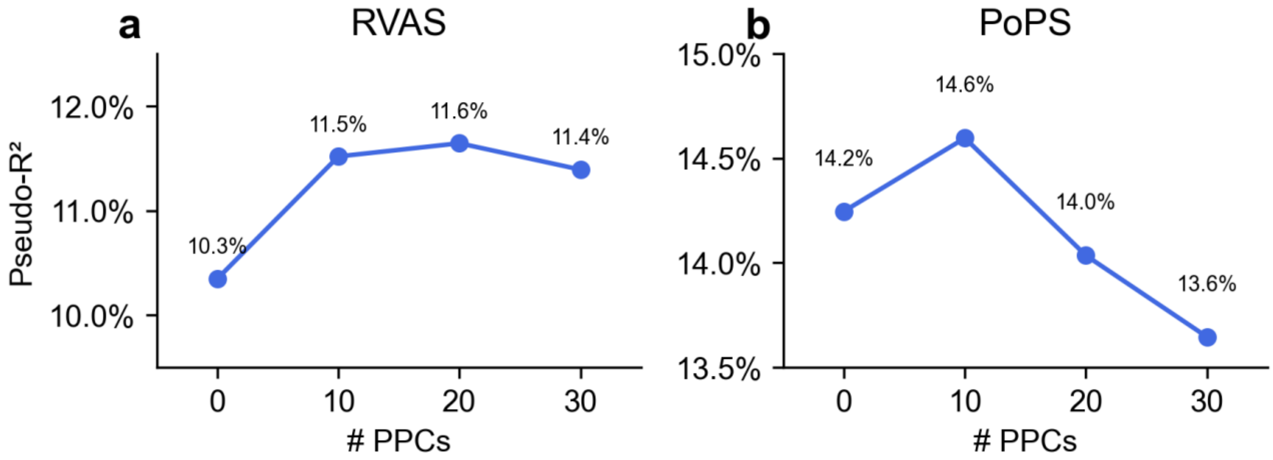


**Supplementary Figure 29. Validation performance as a function of PPCs.** Logistic regression pseudo-$R^2$ for predicting **(a)** RVAS and **(b)** PoPS validation gene set membership across PPCs (0, 10, 20, 30), using cis+trans features (4 bins of |Z|-scores) with UKB-PPP pQTL models.

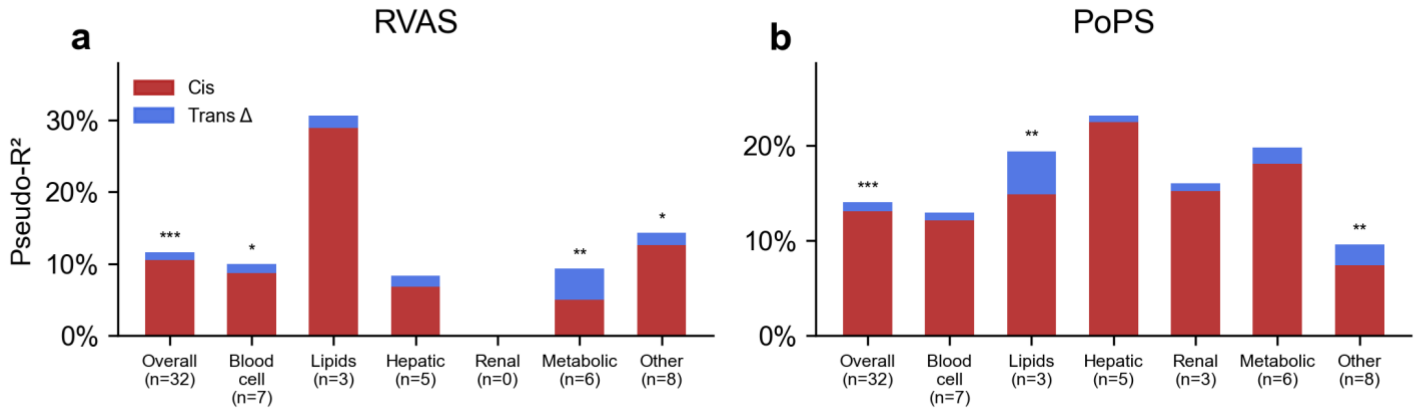


**Supplementary Figure 30. Validation performance across disease/trait categories.** Stacked bars show pseudo-$R^2$ for **(a)** RVAS and **(b)** PoPS across trait categories, with cis-only contribution (red) and added trans contribution (trans Δ, blue), at 20 PPCs using UKB-PPP pQTL models. Asterisks denote likelihood ratio test significance: *P < 0.05, **P < 0.01, ***P < $1\times10^{-4}$.

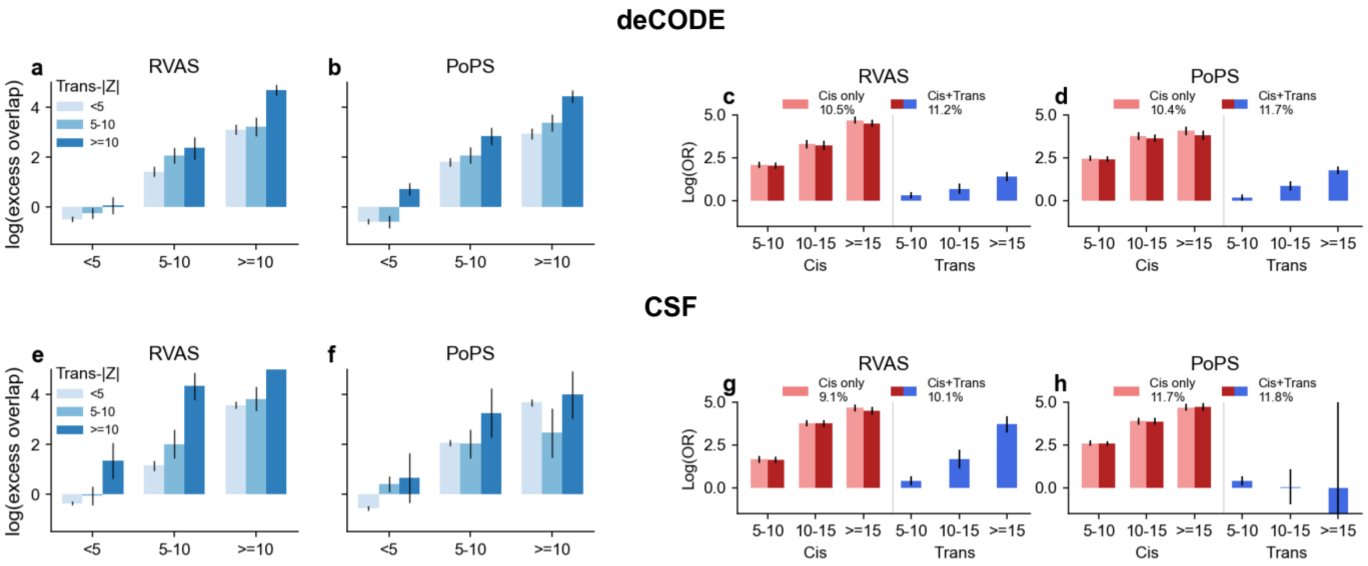


**Supplementary Figure 31. Cross-cohort validation using deCODE (a-d) and CSF (e-h) pQTL models. (a,b,e,f)** Joint cis/trans enrichment for RVAS and PoPS at 20 PPCs. **(c,d,g,h)** Logistic regression coefficients for binned cis-|Z| and trans-|Z|. deCODE results are broadly consistent with UKB-PPP; CSF shows wider confidence intervals due to smaller pQTL sample size.

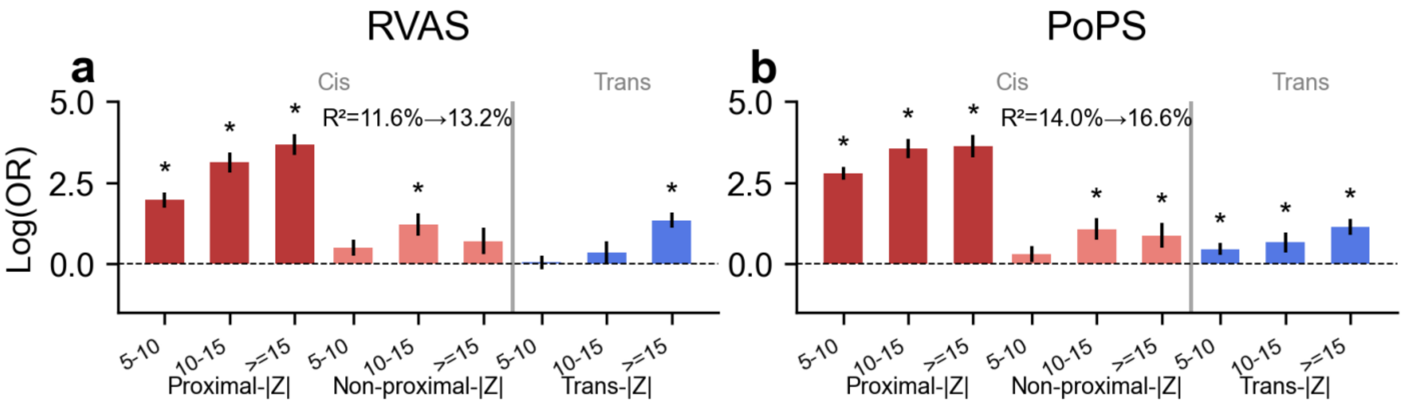


**Supplementary Figure 32. Proximal versus non-proximal cis-PWAS validation.** Logistic regression coefficients (log odds ratio) from models splitting cis features into proximal (within 2 kb of transcribed region) and non-proximal (remaining ±1 Mb) subcomponents, jointly fit with trans-|Z| bins at 20 PPCs, predicting **(a)** RVAS and **(b)** PoPS using UKB-PPP pQTL models. Inset shows pseudo-$R^2$ for unstratified (cis + trans) versus stratified (proximal cis + non-proximal cis + trans) models. Asterisks denote terms with |Z| > 1.96.

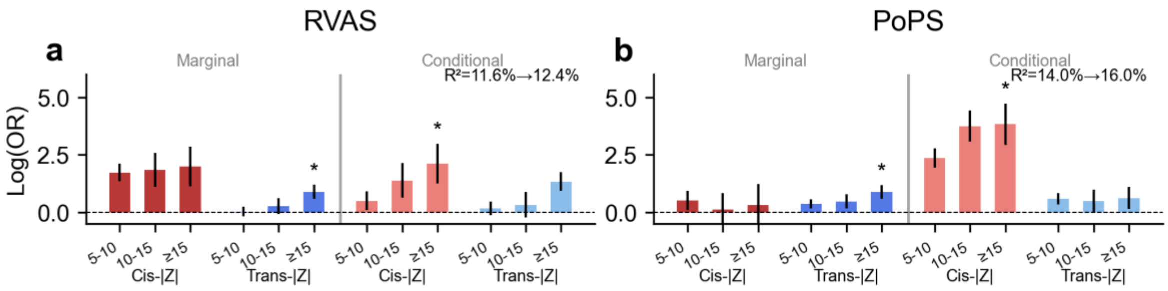


**Supplementary Figure 33. Marginal versus conditional PWAS features in validation models.** Logistic regression coefficients (log odds ratio) from joint models including marginal and conditional (stepwise) cis/trans-

|Z| bins at 20 PPCs using UKB-PPP pQTL models, predicting **(a)** RVAS and **(b)** PoPS. Labels show pseudo-$R^2$ for marginal-only versus marginal+conditional. Asterisks denote terms with |Z| > 1.96.

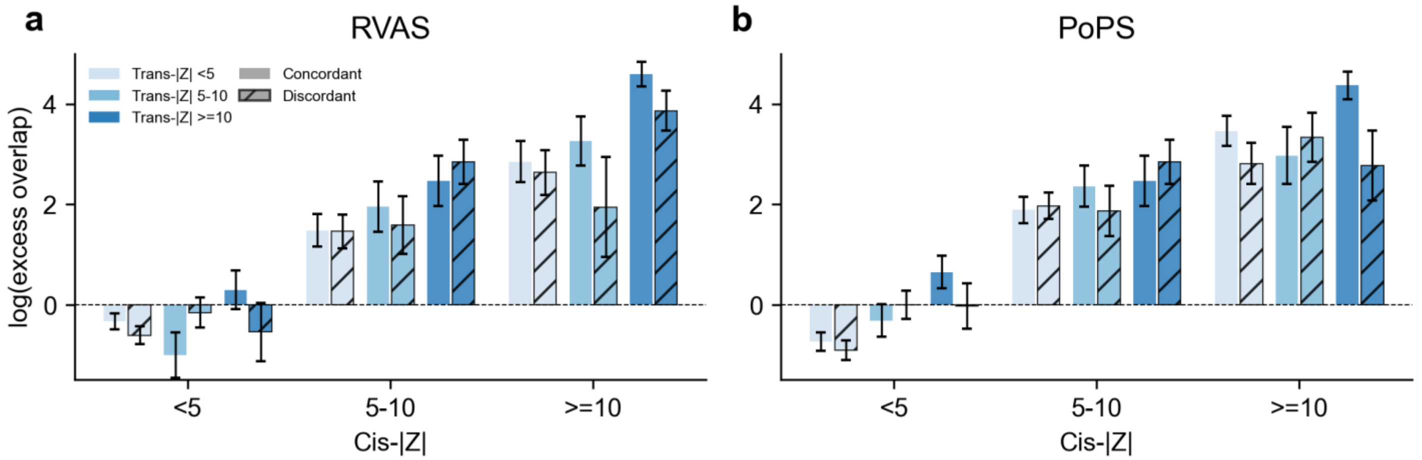


**Supplementary Figure 34. Validation enrichment with concordant versus discordant cis/trans sign.** Log(excess overlap) with **(a)** RVAS and **(b)** PoPS, stratified by cis-|Z| and trans-|Z| bins (colors) at 20 PPCs using UKB-PPP pQTL models. Concordant (solid) and discordant (hatched) bars are shown side by side. Error bars denote standard errors. Enrichment patterns are similar regardless of effect-direction agreement.

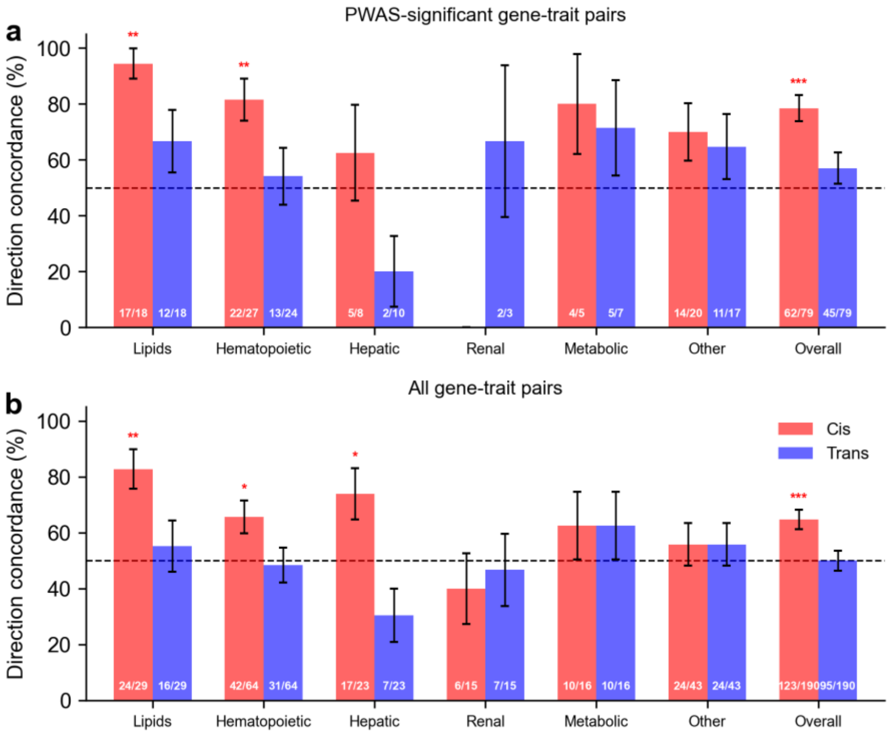


**Supplementary Figure 35. Effect-direction concordance between loss-of-function burden tests and cis/trans-PWAS.** Among Bonferroni-significant gene-trait pairs across 32 UK Biobank traits at 20 PPCs. **(a)** PWAS-significant pairs ($P < 10^{-5}$): cis-PWAS shows concordance (78.5%, $P=3.6 \times 10^{-7}$) whereas trans-PWAS does not (57.0%, P=0.26). **(b)** All burden-significant pairs, stratified by trait category. Dashed line: 50% null. Asterisks: *$P < 0.05$, **$P < 0.01$, ***$P < 1 \times 10^{-4}$.

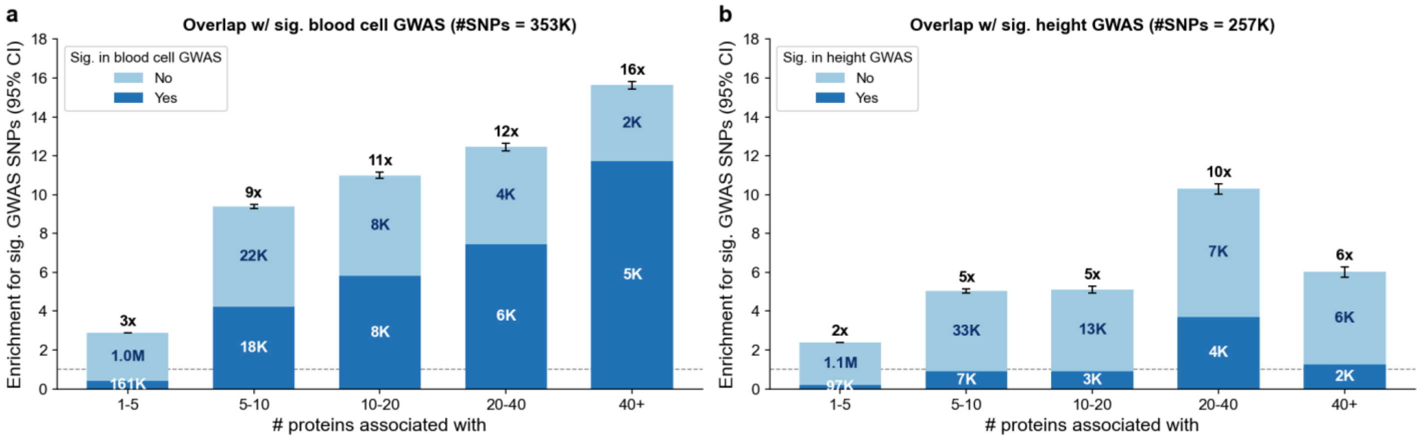


**Supplementary Figure 36. Enrichment of blood cell and height GWAS associations among pleiotropic pQTL loci.** Fold enrichment (with 95% CI) for overlap with genome-wide significant SNPs ($P < 5\times10^{-8}$) from **(a)** blood cell GWAS (353K SNPs across 7 independent blood cell traits) and **(b)** height GWAS (257K SNPs), stratified by the number of proteins each SNP is significantly associated with in UKB-PPP pQTL data ($P < 5\times10^{-8}$). Blood cell GWAS SNPs are strongly enriched among highly pleiotropic pQTL loci (16-fold at > 40 proteins), whereas height GWAS enrichment is more modest (6-fold), indicating that pQTL hotspots are preferentially driven by cell-type composition effects. We note that the different total numbers of genome-wide significant SNPs in blood cell GWAS vs. height GWAS affect the baseline, so identical ratios of overlapping to non-overlapping SNPs within a bin can produce different enrichment magnitudes.

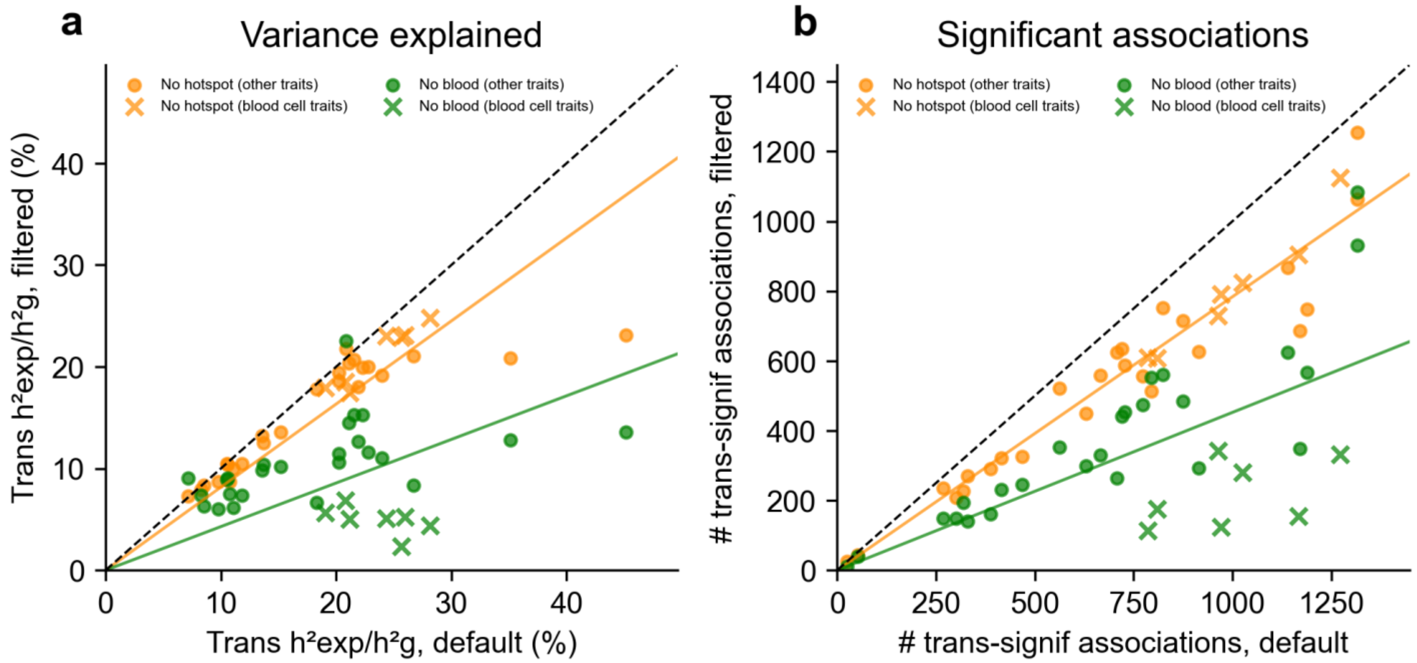


**Supplementary Figure 37. Impact of pleiotropic and cell-type-specific SNP filtering on proportion of variance explained. (a)** Proportion of SNP-heritability explained by trans-predicted protein levels per trait at 20 PPCs: default (x-axis) versus results filtered (y-axis) for pQTL hotspot SNPs ("nohotspot", orange, ~7K SNPs removed, top 0.1% by pQTL pleiotropy) and results filtered for blood cell GWAS SNPs ("noblood", green, 353K blood cell GWAS-significant SNPs removed; union across 7 blood cell traits: white blood cell count, red blood cell count, platelet count, eosinophil count, reticulocyte count, RBC distribution width, platelet distribution width). Crosses denote blood cell traits; circles denote other traits. Solid lines show OLS regression slopes through origin; dashed line indicates y=x. **(b)** Number of significant trans associations per trait: default versus filtered. Blood cell traits show the largest reductions under "noblood" filtering.

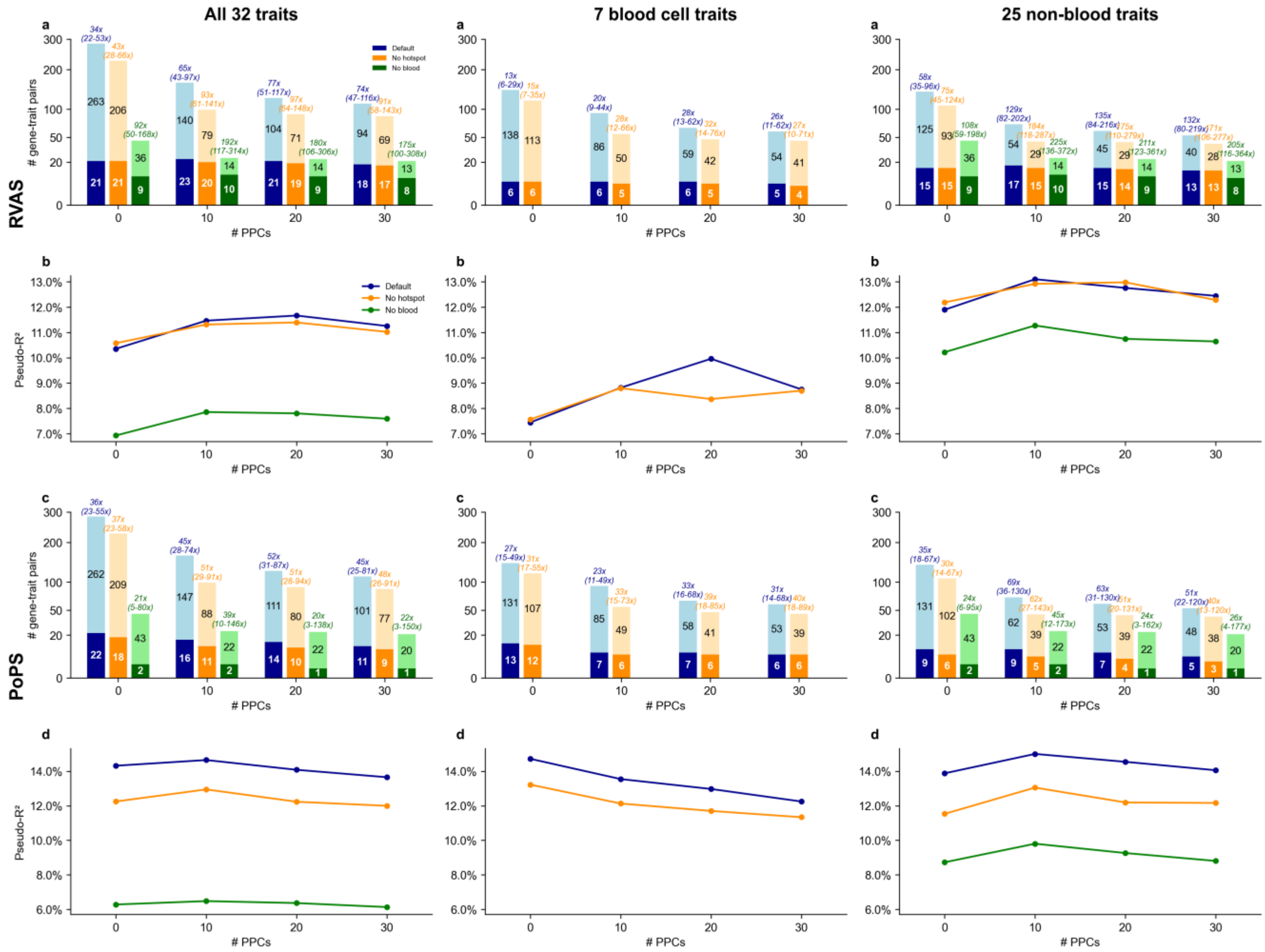


**Supplementary Figure 38. Impact of pleiotropic and cell-type-specific SNP filtering on validation gene set enrichment.** Each column shows results for all 32 traits (left), 7 blood cell traits (middle), and 25 non-blood traits (right). **(a)** Stacked bars showing gene-trait pairs with $|Z_{cis}| > 10$ and $|Z_{trans}| > 10$ that overlap (dark) or do not overlap (light) with RVAS, across PPCs (0, 10, 20, 30). We compared three filtering strategies: default (blue), “nohotspot” (~7K SNPs removed, top 0.1% by pQTL pleiotropy, orange), “noblood” (353K blood cell GWAS-significant SNPs removed, green; union across 7 blood cell traits: white blood cell count, red blood cell count, platelet count, eosinophil count, reticulocyte count, RBC distribution width, platelet distribution width) **(b)** Pseudo-*$R^2$* for RVAS by filtering strategy and PPCs. **(c,d)** Same as **(a,b)** for PoPS. Trans-PWAS prioritization is largely robust to removal of pleiotropic signals for both non-blood cell and blood cell traits, while blood cell trait results are substantially impacted by “noblood” filtering.

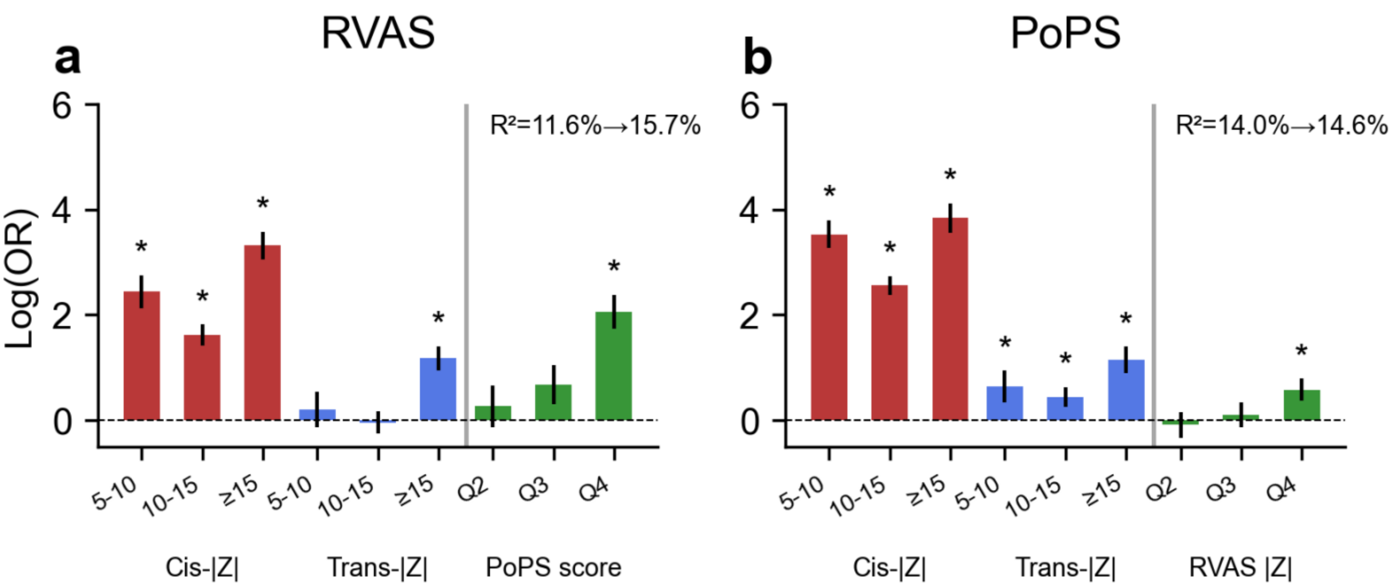


**Supplementary Figure 39. Reciprocal predictive power between RVAS and PoPS.** Logistic regression coefficients (log odds ratio) at 20 PPCs using UKB-PPP pQTL models. **(a)** Predicting RVAS with added PoPS quartiles (pseudo-$R^2$=15.7% vs. 11.6%, P=1.6x$10^{-25}$). **(b)** Predicting PoPS with added RVAS quartiles (pseudo-$R^2$=14.6% vs. 14.0%, P=1.3x$10^{-3}$). The asymmetric improvement indicates that PoPS captures substantial information beyond PWAS for predicting RVAS membership, whereas RVAS adds more modest information for PoPS prediction. Asterisks denote |z| > 1.96.

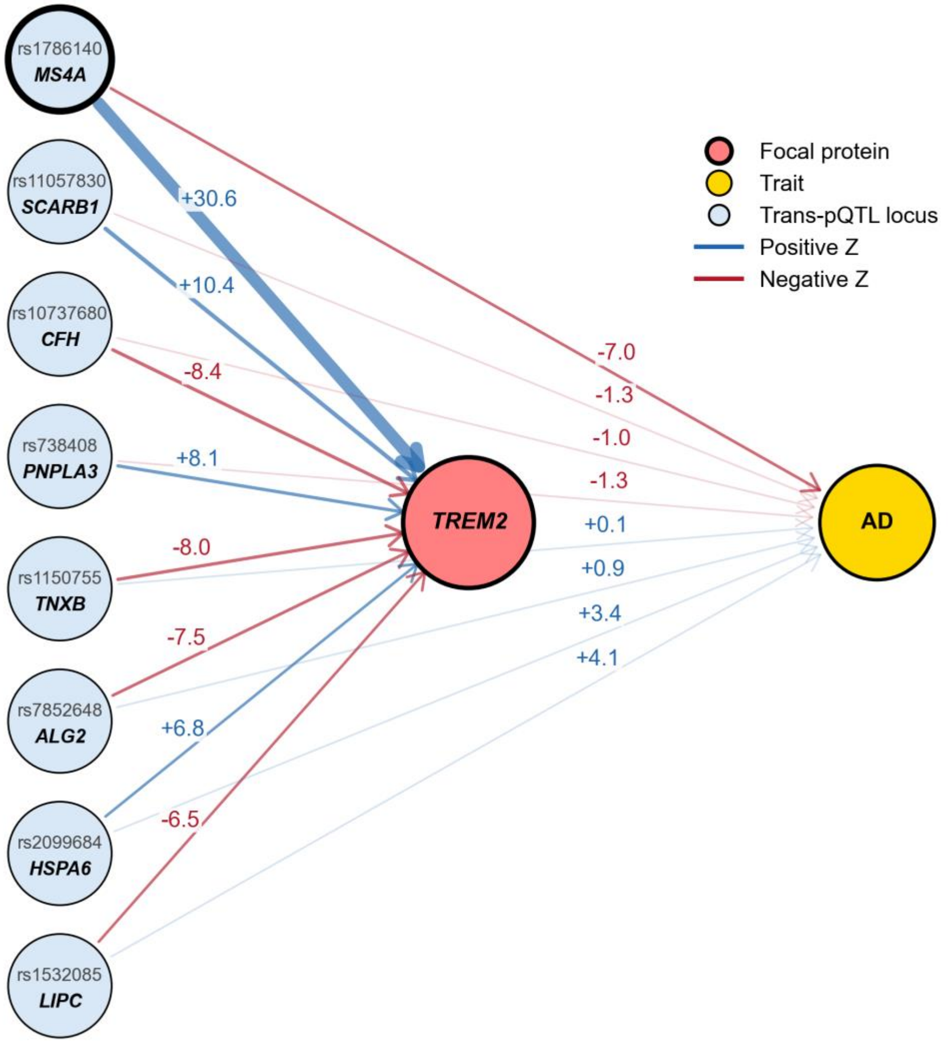


**Supplementary Figure 40. Additional results for association between *TREM2* and AD.** We show trans-pQTL for a focal gene and their corresponding GWAS associations with the trait. Numbers on edges indicate Z-scores: left values show trans-pQTL Z (effect of the SNP on the focal gene), right values show GWAS Z (effect of the SNP on the trait).

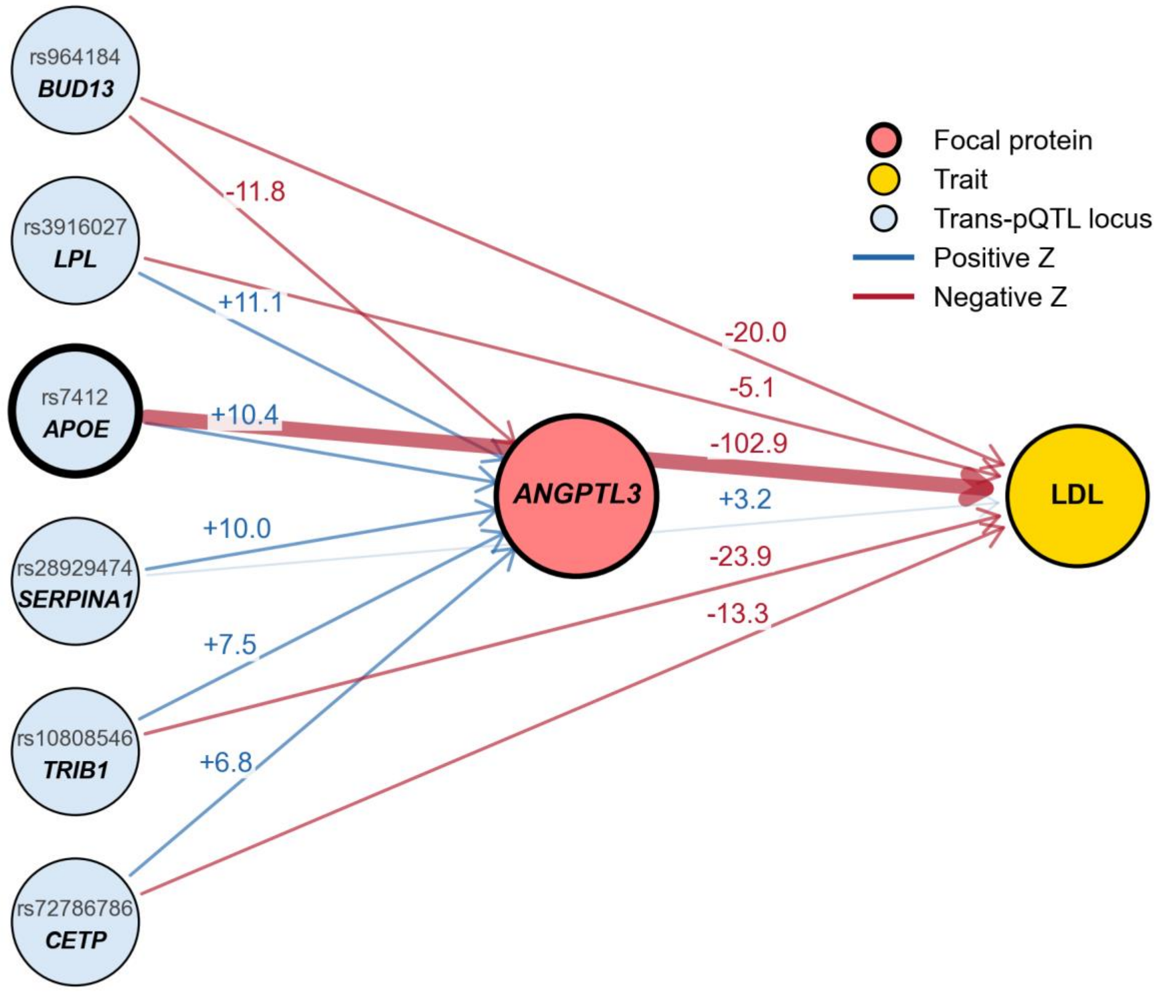


**Supplementary Figure 41. Additional results for association between *ANGPTL3* and LDL.** We show trans-pQTL for a focal gene and their corresponding GWAS associations with the trait. Numbers on edges indicate Z-scores: left values show trans-pQTL Z (effect of the SNP on the focal gene), right values show GWAS Z (effect of the SNP on the trait).

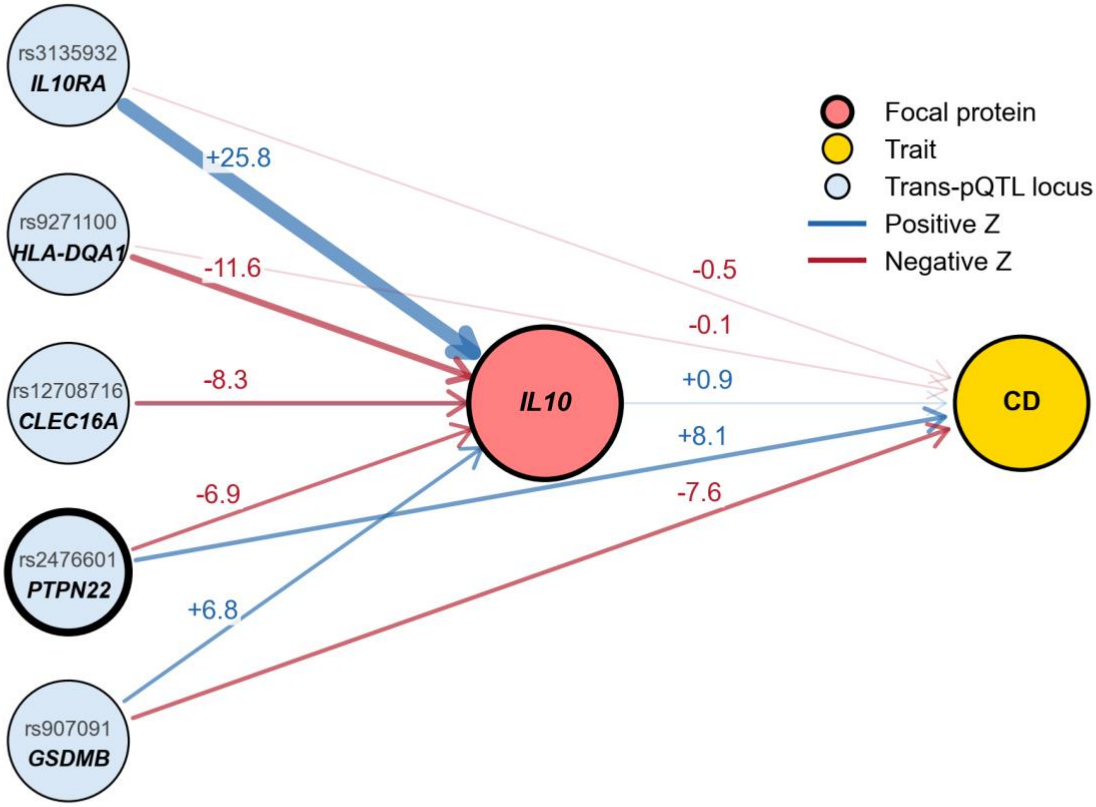


**Supplementary Figure 42. Additional results for association between *IL10* and CD.** We show trans-pQTL for a focal gene and their corresponding GWAS associations with the trait. Numbers on edges indicate Z-scores: left values show trans-pQTL Z (effect of the SNP on the focal gene), right values show GWAS Z (effect of the SNP on the trait).

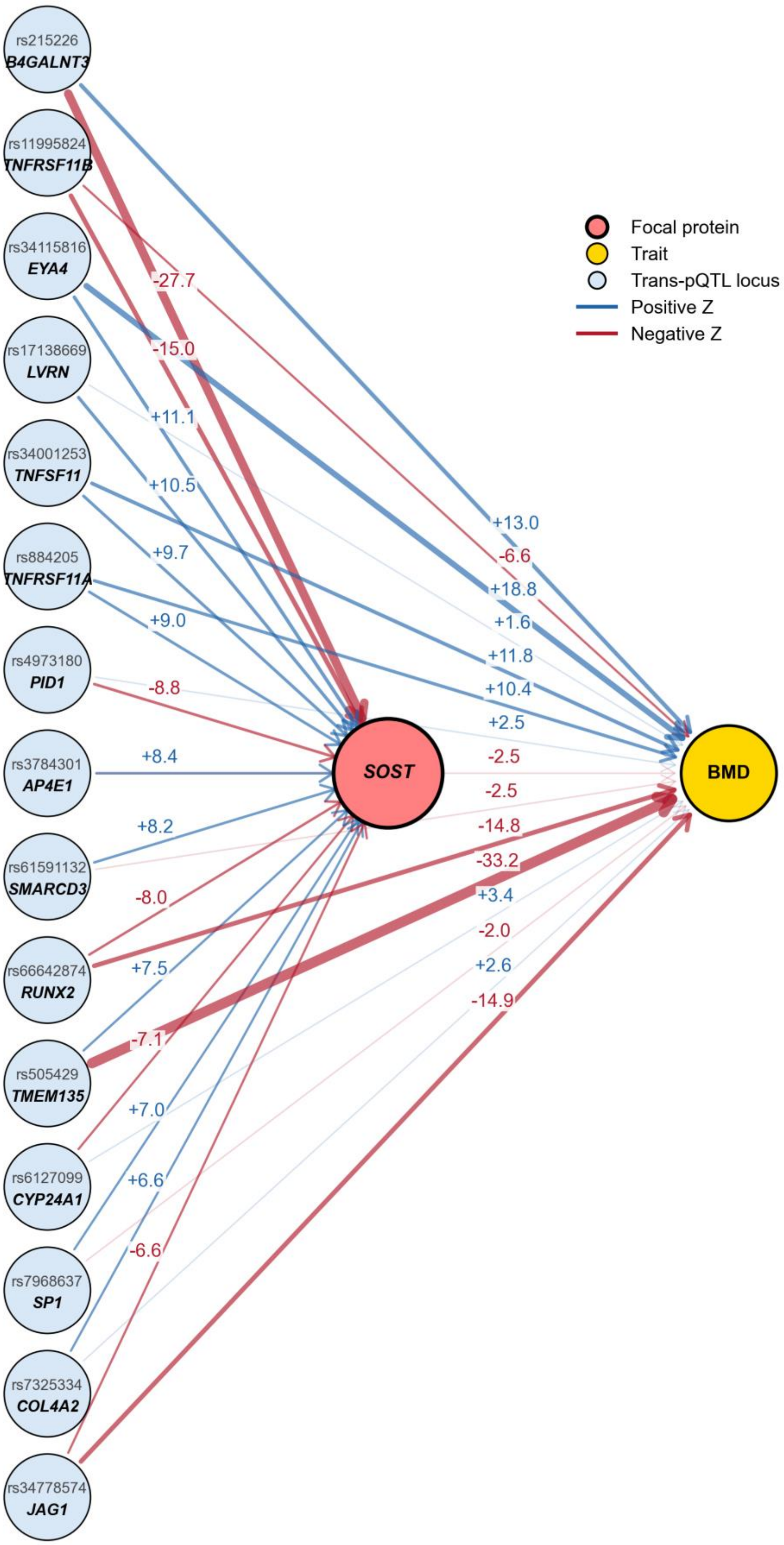

rs215226
B4GALNT3
rs11995824
TNFRSF11B
rs34115816
EYA4
rs17138669
LVRN
rs34001253
TNFSF11
rs884205
TNFRSF11A
rs4973180
PID1
rs3784301
AP4E1
rs61591132
SMARCD3
rs66642874
RUNX2
rs505429
TMEM135
rs6127099
CYP24A1
rs7968637
SP1
rs7325334
COL4A2
rs34778574
JAG1
SOST
BMD
Focal protein
Trait
Trans-pQTL locus
Positive Z
Negative Z
-27.7
-15.0
+11.1
+10.5
+9.7
+9.0
-8.8
+8.4
+8.2
-8.0
+7.5
-7.1
+7.0
+6.6
-6.6
+13.0
-6.6
+18.8
+1.6
+11.8
+10.4
+2.5
-2.5
-2.5
-14.8
-33.2
+3.4
-2.0
+2.6
-14.9


1254

**Supplementary Figure 43. Association between *SOST* and bone mineral density (BMD)**. We highlight an example with discordant cis-PWAS and trans-PWAS effects, suggesting a mixture of direct *SOST* regulation and negative feedback in bone remodeling. PolyPWAS associated *SOST* with BMD (cis Z = −20.4; trans Z = +22.3). *SOST* encodes sclerostin, an inhibitor of bone formation[117]. Loss-of-function mutations cause high-bone-mass phenotypes[118]; the negative cis-PWAS association is directionally consistent with this prior evidence. We identified 15 independent trans-pQTL loci contributing to the trans-PWAS association, 8 of which reach genome-wide significance for BMD. Notably, *TNFSF11* and *TNFRSF11A* encode core regulators of bone remodeling[119], suggesting that part of the trans association reflects upstream bone remodeling activity rather than direct effects of *SOST* itself. In this setting, increased bone remodeling could both increase BMD-related activity and increase *SOST* as a compensatory brake, producing a positive trans-PWAS contribution. By contrast, *B4GALNT3* is linked to sclerostin clearance, so lower sclerostin levels would be expected to increase BMD[120]. Together, these loci support a model in which the trans-PWAS association combines upstream feedback in bone remodeling and more direct regulation of circulating sclerostin.

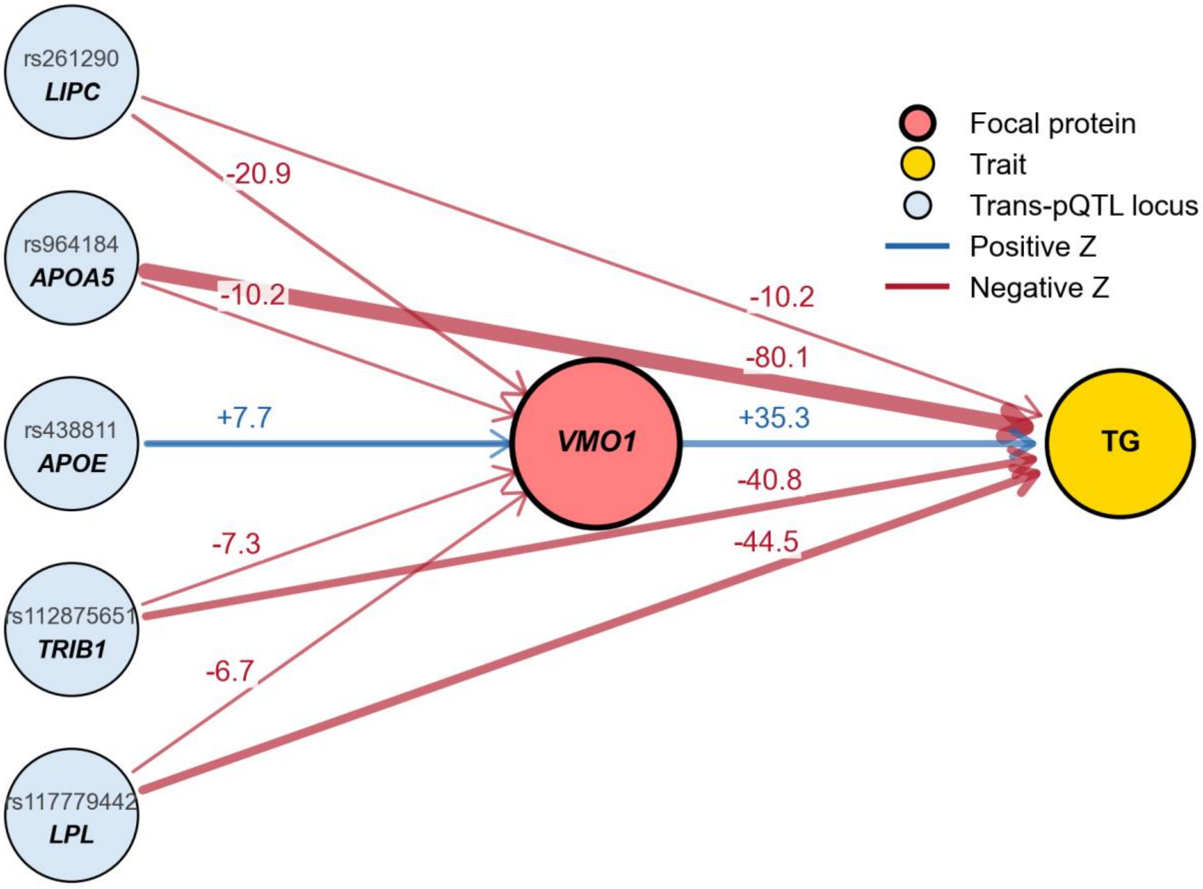


**Supplementary Figure 44. Association between *VMO1* and triglycerides (TG).** We highlight an example involving concordant cis-PWAS and trans-PWAS effects. PolyPWAS associated *VMO1* with triglycerides (cis Z = +5.1; trans Z = +59.3). *VMO1* has not been widely studied for its role in lipid regulation. The trans-PWAS association was driven by five independent trans-pQTL loci. These loci map to established lipid regulators, including *LIPC*, *APOA5, APOE, TRIB1, LPL*[121]. Together, the concordant cis and trans associations place *VMO1* within core triglyceride-regulatory networks, although the strong trans signal may partly reflect tagging through these lipid loci.

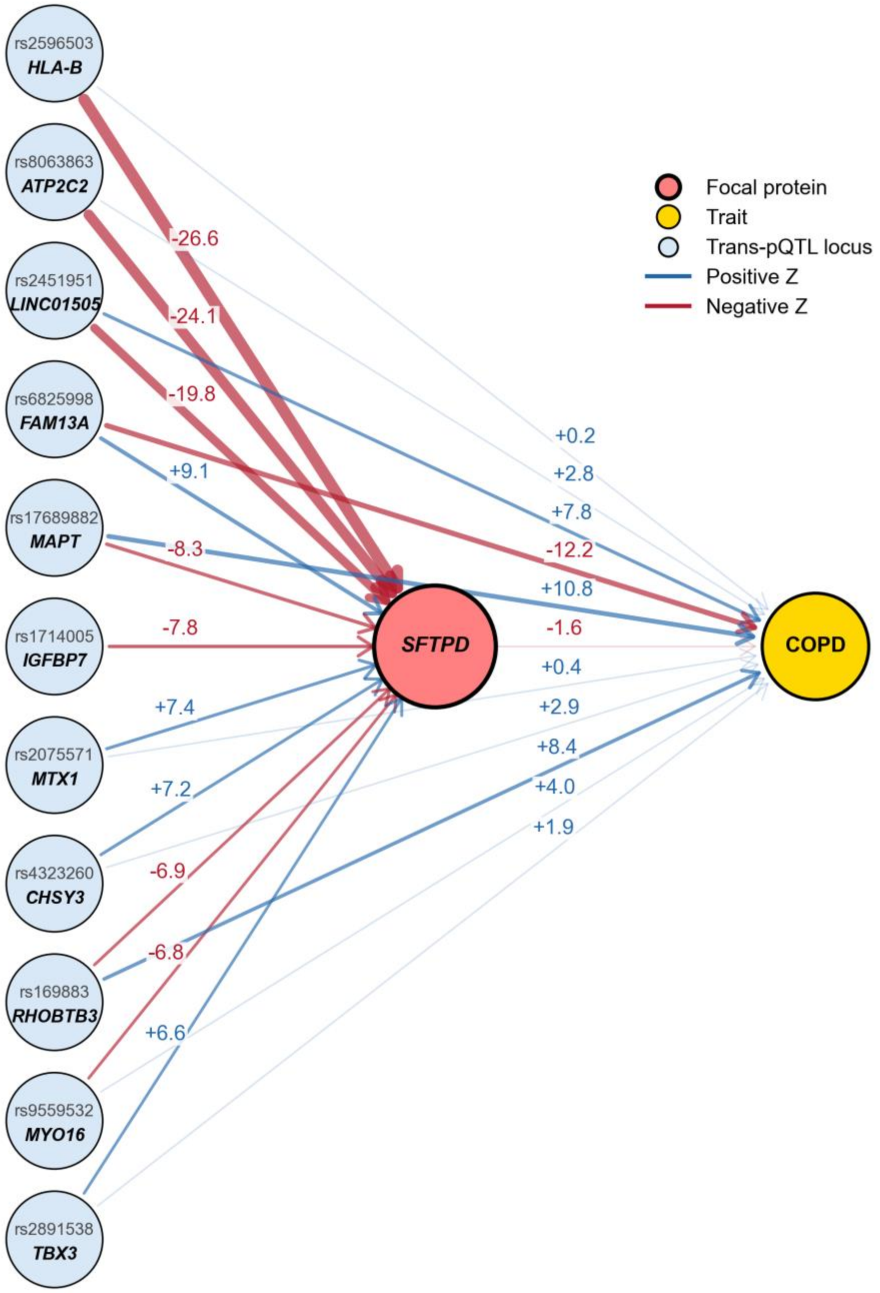


**Supplementary Figure 45. Association between *SFTPD* and chronic obstructive pulmonary disease (COPD).** We highlight an example with concordant cis-PWAS and trans-PWAS effects, where the trans-PWAS effect reflects a likely causal relationship. PolyPWAS associated *SFTPD* with COPD (cis Z = −7.2; trans Z = −8.5). *SFTPD* encodes surfactant protein D (SP-D), a soluble innate immune collectin enriched in alveolar epithelial cells. Prior Mendelian randomization analyses using serum SP-D-associated variants found that genetically higher serum SP-D was associated with lower COPD risk and slower FEV1 decline[122]. Our cis- and trans-PWAS results are consistent with that direction of effect. In our analysis, the negative trans-PWAS association was driven by 11 independent trans-pQTL, 4 at loci associated with COPD. For example, *FAM13A* is a well-established COPD susceptibility gene expressed in alveolar epithelial cells[123,124], suggesting that part of the trans association may reflect distal regulation in a cellular context relevant to SP-D biology. Together, the concordant cis and trans associations support that SP-D level is critical to COPD risk.

### Supplementary Tables

**Supplementary Table 1. pQTL studies**. pQTL datasets used in this study, including assay platform, tissue, cohort sample size, number of assayed proteins for UKB-PPP, deCODE, and CSF.

**Supplementary Table 2. GWAS studies.** GWAS datasets analyzed in this study, including trait sample size, LDSC heritability estimates, and study reference for 32 approximately independent UK Biobank diseases/traits (UKB-PPP excluded) and 56 additional GWAS. Mean observed-scale SNP-heritability is 0.216 for the 32 UK Biobank traits and 0.158 for the 56 additional traits (mean N = 364,098 and 320,592, respectively).

**Supplementary Table 3. Simulation statistics.** False positive rate or power, confidence intervals, estimated pQTL heritability, prediction accuracy varying pQTL heritability, genetic correlation, mediated versus non-mediated effects, and GWAS/pQTL polygenicity.

**Supplementary Table 4. Protein-level prediction.** UKB-PPP, deCODE, and CSF cis and trans protein prediction performance summaries across LD reference, prediction method, and PPC adjustments, reported as median and mean prediction $R^2$.

**Supplementary Table 5. Co-regulation analyses.** UKB-PPP, deCODE, and CSF cis and trans co-regulation score summaries across model settings and PPC adjustments.

**Supplementary Table 6. Cross-cohort genetic-correlations.** UKB-PPP vs. deCODE cross-cohort statistics for matched proteins, including cis and trans prediction concordance and cross-cohort genetic-correlation estimates.

**Supplementary Table 7. Trait heritability explained by predicted proteins.** Proportions of SNP-heritability explained by cis-predicted proteins, trans-predicted proteins, and their joint model for pQTL groups across PPC adjustments.

**Supplementary Table 8. PWAS discovery summary.** Summary for number of significant cis and trans PWAS associations across datasets and PPC adjustments.

**Supplementary Table 9. Cross-cohort PWAS concordance.** Cross-cohort PWAS concordance summary of UKB-PPP vs. deCODE and UKB-PPP vs. CSF at 20 PPCs.

**Supplementary Table 10. RVAS/PoPS validation overlap analyses.** Numerical results of validation analyses for RVAS and PoPS targets, including excess overlap effect size estimates for high-confidence PWAS bins across cis/trans strata.

**Supplementary Table 11. Logistic-regression gene-prioritization models.** Numerical results of logistic regression predicting RVAS and PoPS membership from PWAS association statistics.

**Supplementary Table 12. PolyPWAS runtime in UKB-PPP.** We report runtime for UKB-PPP including both protein prediction model training and association testing (Equation 1), for well-imputed versus HapMap3 SNPs.
